# X-Chromosome-wide association study for Alzheimer’s disease

**DOI:** 10.1101/2024.05.02.24306739

**Authors:** Julie Le Borgne, Lissette Gomez, Sami Heikkinen, Najaf Amin, Shahzad Ahmad, Seung Hoan Choi, Joshua Bis, Benjamin Grenier-Boley, Omar Garcia Rodriguez, Luca Kleineidam, Juan Young, Kumar Parijat Tripathi, Lily Wang, Achintya Varma, Sven van der Lee, Vincent Damotte, Itziar de Rojas, Sagnik Palmal, Vilmantas Giedraitis, Roberta Ghidoni, Victoria Fernandez, Patrick Gavin Kehoe, Ruth Frikke-Schmidt, Magda Tsolaki, Pascual Sánchez-Juan, Kristel Sleegers, Martin Ingelsson, Jonathan Haines, Lindsay Farrer, Richard Mayeux, Li-San Wang, Rebecca Sims, Anita DeStefano, Gerard D. Schellenberg, Sudha Seshadri, Philippe Amouyel, Julie Williams, Wiesje van der Flier, Alfredo Ramirez, Margaret Pericak-Vance, Ole Andreassen, Cornelia Van Duijn, Mikko Hiltunen, Agustín Ruiz, Josée Dupuis, Eden Martin, Jean-Charles Lambert, Brian Kunkle, Céline Bellenguez

## Abstract

Due to methodological reasons, the X-chromosome has not been featured in the major genome-wide association studies on Alzheimer’s Disease (AD). To finally address this and better characterize the genetic landscape of AD, we performed an in-depth X-Chromosome-Wide Association Study (XWAS) in 115,841 AD cases or AD proxy cases, including 52,214 clinically-diagnosed AD cases, and 613,671 controls. We considered three approaches to account for the different X-chromosome inactivation (XCI) states in females, i.e. random XCI, skewed XCI, and escape XCI. We did not detect any genome-wide significant signals (P ≤ 5 × 10^−8^) but identified four X-chromosome-wide significant loci (P ≤ 1.7 × 10^−6^). Two signals locate in the *FRMPD4* and *DMD* genes, while the two others are more than 300 kb away from the closest protein coding genes *NLGN4X* and *GRIA3*. Overall, this XWAS found no common genetic risk factors for AD on the non-pseudoautosomal region of the X-chromosome, but it identified suggestive signals warranting further investigations.

## Introduction

Alzheimer’s disease (AD) is a progressive neurodegenerative disease and the most common cause of dementia among the elderly. AD is caused by a combination of modifiable and non-modifiable risk factors, including genetics. Currently, more than 80 genetic loci are associated with AD risk, highlighting several underlying biological mechanisms for AD, including APP metabolism, Tau-mediated toxicity, lipid metabolism or immune-related processes^1–6^. Greater understanding of the genetics of AD is essential to improve the characterization of the pathophysiological processes involved in the disease. However, although the genetic landscape of AD has been extensively studied on the autosomes, little is known about the association of the X-chromosome gene variants with AD risk. To date, large-scale genome-wide association studies (GWAS) did not include the X-chromosome due to the need of specific analyses to account for its features.

While women carry two copies of the X-chromosome, men are hemizygous, meaning they have one X and one Y chromosome. To maintain balance around allelic dosage between the sexes, X-chromosome inactivation (XCI) occurs in females. This process is where one X chromosome is transcriptionally silenced during female development^7,8^. The choice of the silenced copy is most often random, but inactivation can also be skewed toward a specific copy. Such XCI ‘skewness’ can be subsequently acquired during life and has been described to increase with age in adults ^9–12^. Importantly, up to one‐third of X‐chromosome genes ‘escape’ inactivation and are expressed from both X‐chromosomes in female cells. However, these tend to be expressed less from the inactive X-chromosome. Notably, all the genes in the pseudoautosomal region (PAR) 1 of the X-chromosome have Y-chromosome homologues and escape inactivation. Additionally, some genes variably escape inactivation: their expression from the inactive X-chromosome differs between individuals or between cells and tissues within an individual^8,13^. The inactivation process and the distinction between the PAR and non-PAR regions are thus important considerations when performing an X-chromosome-wide association study (XWAS). For all these reasons, the X-chromosome needs to be treated separately from the autosomes in the quality control (QC), the imputation process and the analysis^14,15^, and has usually been excluded from GWAS, including for the large-scale AD ones. Yet, the X-chromosome represents about 5% of the genome in terms of size and number of genes (UCSC Genome Browser, https://genome.ucsc.edu/cgi-bin/hgTracks?db=hg38&chromInfoPage=), and thus the study of AD genetics remains incomplete.

Several X-chromosome genes have been associated with brain imaging phenotypes^16,17^. Furthermore, the X-chromosome carries, disproportionately for the whole genome, more than 15% of the known genes related to intellectual disabilities^18^. While genes related to intellectual disabilities are considered to modulate early neurodevelopmental stages well before neurodegenerative processes start, they might impact on the development of cognitive abilities and, potentially, on the establishment of cognitive reserve and brain resilience^19^. Additionally, XCI escape or skewness might contribute to observed sex differences reported in AD. Women have a higher risk of developing dementia than men: in the 65-69 and 85-89 age groups, the prevalence is 1.5% and 24.9% respectively for women, compared with 1.1% and 16.3% for men^20,21^. This difference can be partly explained by a greater longevity of women, but other factors may also be involved, such as a selective survival bias in men, socio-environmental factors, or different AD-related biological mechanisms between sexes^22^. Male and female differences have also been observed for AD and AD-related phenotypes, such as cognitive performance or in the impact of *APOE* variants on the disease risk or on Tau concentration, and such differences may be explained by XCI escape and skewness^8,23–25^. Consistent with this, in AD mouse models, having two X-chromosomes was associated with reduced mortality and cognitive impairment. This advantage conferred by a second X-chromosome could partly relate to the *KDM6A* gene, which escapes inactivation. A variant of the human version of this gene was associated with an increase in this gene’s expression in the brain, and with less cognitive decline in aging and preclinical AD^26^. Finally, in humans, expression/level of other X-linked genes or proteins are reportedly associated with cognitive change or tau pathology in a sex-specific manner^27,28^.

To investigate the impact of X-chromosome genetic variants on AD risk, we conducted an in-depth XWAS on 115,841 AD cases or AD proxy cases and 613,671 controls from the IGAP (International Genomics of Alzheimer’s Project), EADB (European Alzheimer & Dementia Biobank), UK Biobank and FinnGen studies (Supplementary Table S1). We considered three approaches to account for the different inactivation states in females, i.e., random XCI (r-XCI), skewed XCI (s-XCI), and escape XCI (e-XCI)^15^.

## Results

A total of 288,320, 276,902 and 263,169 common variants (minor allele frequency or MAF ≥ 1%) were analyzed in the r-XCI, e-XCI and s-XCI approaches, respectively. We observed a minor deviation from expected p-values in the r-XCI and e-XCI models (median genomic inflation factor λ = 1.074 and 1.087, respectively) and a deflation in the s-XCI model (median λ = 0.735), likely related to a lack of power (Supplementary material, Supplementary Figures S1-S3 and Supplementary Table S2). We did not identify any genome-wide significant signals (P ≤ 5 × 10^−8^) among X-chromosome common variants in any of the models (Figures 1, 2 and 3). However, three loci exhibited signals that were X-chromosome-wide significant (P ≤ 1.7 x 10^-6^) in the r-XCI approach: Xp22.32, *FRMPD4* and Xq25 (Figure 1, Table 1). No X-chromosome-wide significant signal was found in the e-XCI or s-XCI analyses (Figures 2 and 3). As expected, we observed correlated results between the r-XCI and e-XCI meta-analysis results (Supplementary Table S3).

**Table 1:** Summary of association analysis results with an X-chromosome-wide significant signal. P values are two-sided raw P values derived from a fixed-effect meta-analysis. CI, confidence interval; OR, odds ratio; MAF, minor allele frequency. ^a^Reference single-nucleotide polymorphism (SNP) (rs) number, according to dbSNP build 153, ^b^GRCh38 assembly, ^c^Nearest protein-coding gene according to GENCODE release 45, ^d^from Tukiainen et al., 2017^13^, ^e^Weighted average MAF across all discovery studies, ^f^Approximate OR calculated with respect to the minor allele.

| Variant<sup>a</sup> | Model | Meta-analysis | Position<sup>b</sup> | Gene<sup>c</sup> | Inactivation status<sup>d</sup> | Minor/<br>major<br>allele | MAF<sup>e</sup> | OR <sup>f</sup> | 95% CI | P-value |
| --- | --- | --- | --- | --- | --- | --- | --- | --- | --- | --- |
| rs4364769 | r-XCI | AD-proxy | 5,462,201 | *NLGN4X*<br>(Xp22.32) | variable | T/G | 0.12 | 1.079 | 1.048-1.110 | 2.55 x 10<sup>-7</sup> |
| rs5933929 |  |  | 11,916,372 | *FRMPD4* | inactive | C/A | 0.38 | 0.952 | 0.935-0.970 | 1.98 x 10<sup>-7</sup> |
| rs5972406 |  | excluding biobanks | 31,546,147 | *DMD* | inactive | A/G | 0.08 | 1.143 | 1.083-1.207 | 1.16 x 10<sup>-6</sup> |
| rs191195705 |  | AD-proxy | 122,643,733 | *GRIA3* (Xq25) | NA | A/C | 0.11 | 0.925 | 0.896-0.954 | 7.09 x 10<sup>-7</sup> |

**Figure 1:**
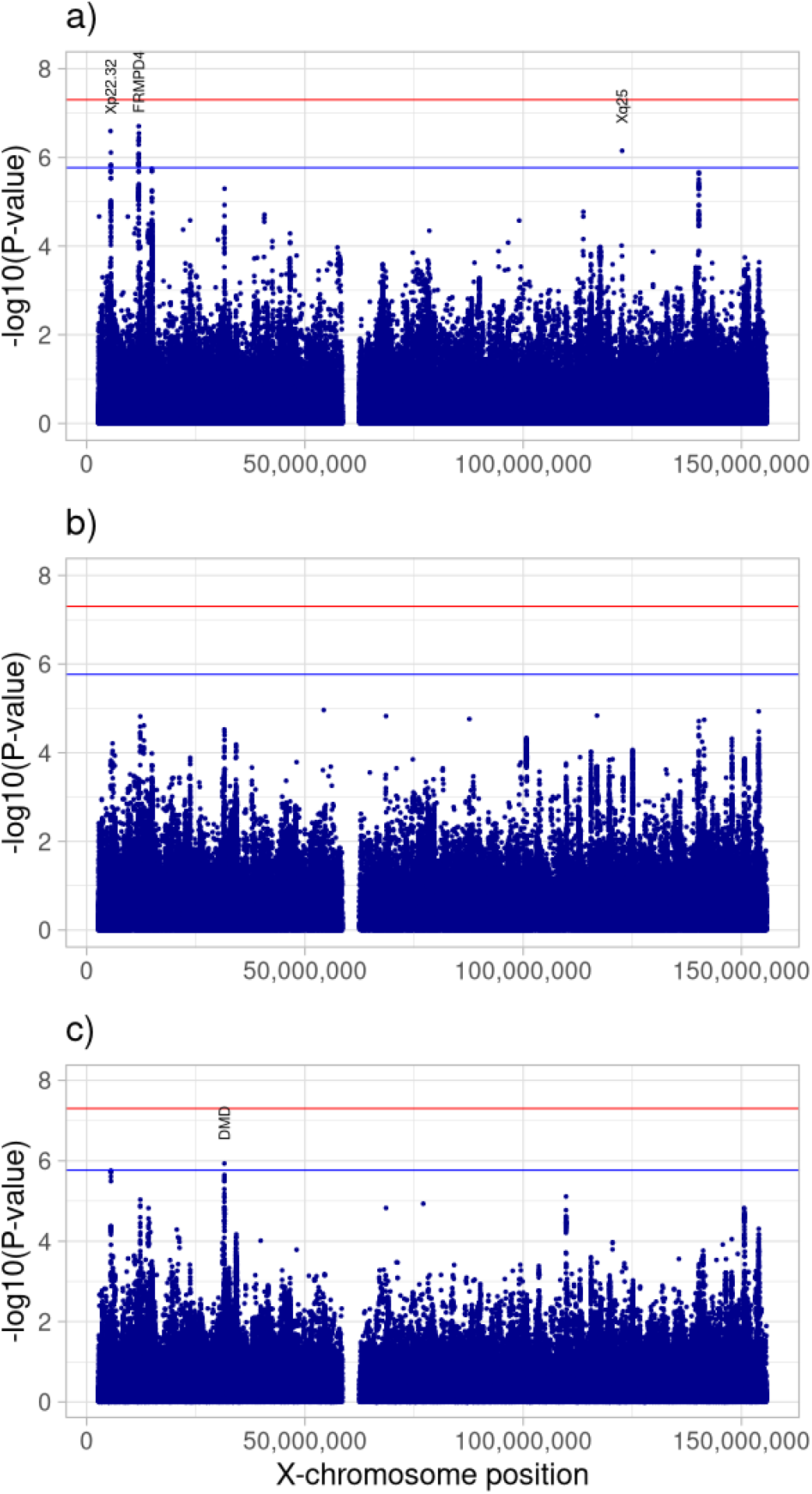
Manhattan plot of common variants (MAF ≥ 0.01) for the r-XCI approach in a) the meta-analysis including AD-proxy cases, b) the diagnosed AD cases meta-analysis and c) the meta-analysis excluding biobanks. The red and blue lines represent the genome-wide significant threshold (5 x 10^-8^) and the X-chromosome-wide significant threshold (1.7 x 10^-6^), respectively. The labels show the closest protein-coding gene (according to GENCODE release 45, https://www.gencodegenes.org/human/releases.html) to the index variant of each X-chromosome-wide significant locus.

**Figure 2:**
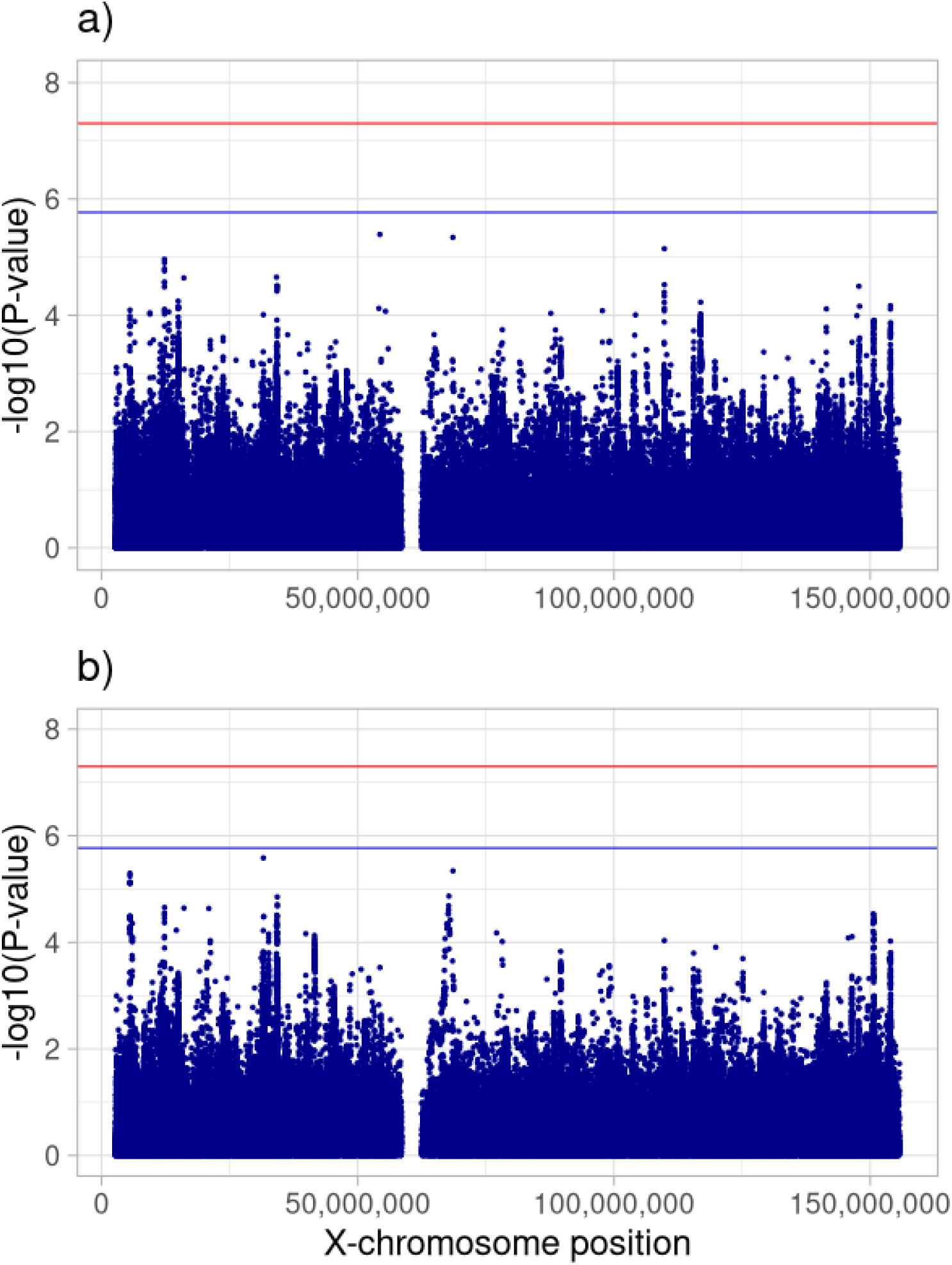
Manhattan plot of common variants (MAF ≥ 0.01) for the e-XCI approach in a) the diagnosed AD-cases meta-analysis and c) the meta-analysis excluding biobanks. The red and blue lines represent the genome-wide significant threshold (5 x 10^-8^) and the X-chromosome-wide significant threshold (1.7 x 10^-6^), respectively.

**Figure 3:**
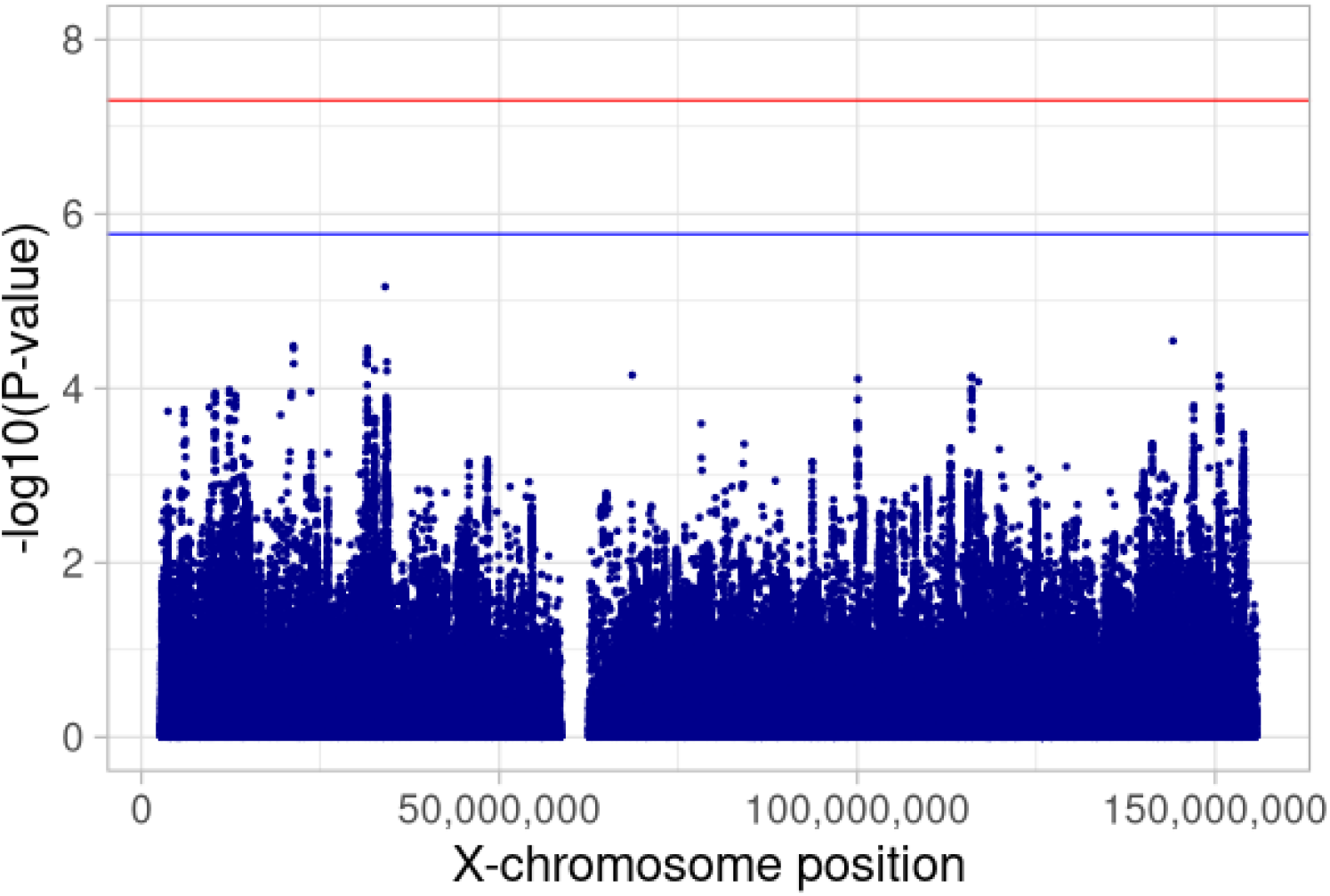
Manhattan plot of common variants (MAF ≥ 0.01) for the s-XCI approach meta-analysis, which excludes biobanks. The red and blue lines represent the genome-wide significant threshold (5 x 10^-8^) and the X-chromosome-wide significant threshold (1.7 x 10^-6^), respectively.

In more detail, rs4364769 (MAF = 0.12, OR = 1.079 [1.048-1.110], P = 2.55 x 10 ^-7^) was identified as the index variant of the Xp22.32 locus in the r-XCI meta-analysis (Table 1, Supplementary Figure S4). Several sensitivity analyses of this signal were performed, for example by excluding AD-proxy or biobank samples, or by further adjusting the analyses on age or *APOE* (Online Methods). The odds-ratio estimate of rs4364769 shows some variability across sensitivity analyses but confidence intervals overlap (Supplementary Table S4). The index variant of the Xp22.32 signal is located more than 300kb from the closest protein coding gene, *NLGN4X (*Neuroligin 4 X-Linked).

The index variant in the *FRMPD4* (FERM and PDZ Domain Containing 4) locus was rs5933929 (MAF = 0.38, OR = 0.952 [0.935-0.970], P = 1.98 x 10^-7^) in the r-XCI meta-analysis (Table 1, Supplementary Figure S5). This variant is located in an intron within some transcripts of *FRMPD4*. The odds-ratio of rs5933929 was consistent across sensitivity analyses (Supplementary Table S4).

rs191195705 was the index variant in the Xq25 signal in the r-XCI meta-analysis (MAF = 0.11, OR = 0.925 [0.896-0.954], P = 7.09 x 10^-7^, Table 1). Here the males and the UK Biobank (UKB)-proxy males carried a large part of the observed effect, leading to a lower signal in the sensitivity analyses excluding proxy or biobank cases, or in the female-only compared to the male-only meta-analyses (Supplementary Table S4, Supplementary Figure S6). However, the difference of effect between males and females was not significant (P = 0.51, Online Methods, Supplementary Table S4). rs191195705 is over 500 kb from the closest protein coding gene, *GRIA3* (Glutamate Ionotropic Receptor AMPA Type Subunit 3).

To account for potential results that we may have missed because of false negatives related to proxy samples or biobanks, we also performed the r-XCI and e-XCI analyses on the whole X-chromosome excluding these samples (note: samples from biobanks, including proxy, were not included in the s-XCI analysis in the first place, Online Methods). We did not identify any genome-wide significant signals among X-chromosome common variants, in any of the models, nor any X-chromosome-wide significant signal when considering only AD diagnosed cases (Figures 1 and 2, Supplementary Figures S1-S2). However, one X-chromosome-wide significant locus was identified in the r-XCI meta-analysis excluding biobanks (Table 1). The index variant was rs5972406 (MAF = 0.075, OR = 1.143 [1.083-1.207], P = 1.16 x 10^-6^), located in an intron of the *DMD* dystrophin gene (Table 1, Supplementary Figure S7).

As the XCI mechanism induces variability across females, one might expect stronger effects in males compared to females; we therefore performed an additional sex-stratified analysis, excluding proxy cases (Online Method), and compared the variant effect sizes in males and females. We did not identify any genome-wide nor X-chromosome-wide significant signals in either the male-only or female-only meta-analyses (Supplementary Figure S8). We also did not observe any genome-wide nor X-chromosome-wide significant difference of effect between males and females for any X-chromosome variants (Supplementary Figure S8).

## Discussion

We conducted the most comprehensive XWAS on AD to date, including 115,841 AD or AD-proxy cases and 613,671 controls and using three complementary models to account for the complexity related to this chromosome. Despite not detecting any genome-wide significant signals regardless of the approach used, we identified four X-chromosome-wide significant loci.

The signal in the *FRMPD4* locus was consistent across the sensitivity analyses, showing strong robustness, while the other signals in *NLGN4X, GRIA3* and *DMD* showed some variability.

*FRMPD4* (FERM and PDZ domain containing 4) is mostly expressed in brain tissues (GTex Portal, https://gtexportal.org/). Through its interaction with other proteins, the FRMPD4 protein is involved in the regulation of the morphogenesis and density of dendritic spines, and in the maintenance of excitatory synaptic transmission^*29*^. *FRMPD4* is an X-linked intellectual disability gene^30^ and is associated with low educational attainment^31^. The associated variant is in an intron within some transcripts of *FRMPD4* but is also close to the *MSL3* gene, which interacts with *KAT8*, a reported genetic risk factor for AD^2,32,33^. In addition, *FRMPD4* is an inactivated gene in females, while *MSL3* escapes inactivation^13^.

The signal at the intronic variant within the *DMD* dystrophin gene decreased when including proxy or biobank cases; further analyses are necessary to determine whether this is due to a falsely inflated signal in the clinically diagnosed samples, or to a less specific diagnosis in the proxy and biobank samples. *DMD* is inactivated in females^13^, and mutations in the gene can cause Duchenne muscular dystrophy. Some patients suffering from this disease can exhibit cognitive impairment, and a shift towards amyloidogenesis in memory-specific brain regions was found in mice mutated in the *DMD* gene (mdx mouse) compared to wild-type mice^34^. Additionally, the *DMD* rs5927116 variant was reportedly associated with the volume of entorhinal cortex in a small sample (N = 792); however, this signal is 1.4 Mb away from our AD signal and the variants are independent (LD measured by r^2^ < 0.2)^35^.

Identifying putative causal genes in the two other loci, Xp22.32 and Xq25, is more challenging, as the index variants are located more than 300 kb away from the closest protein coding gene, *NLGN4X* and *GRIA3*, respectively. Additionally, those variants are not eQTL/sQTL for any gene according to GTeX Portal. Expression of the *GRIA3* gene in the dorsolateral prefrontal cortex is reportedly associated with cognitive change in women during aging and AD^27^. However, the rs191195705 index variant of the Xq25 signal is associated with AD risk mainly in males in our analyses (Supplementary Table S4). Regarding the Xp22.32 locus, the rs5916169 variant, located at 127 kb from our index variant, is associated with functional connectivity^16^. However, this variant is not in LD (r^2^ = 0.005) with the AD index variant.

Although this study represents the largest XWAS for AD risk to date, we did not find any genome-wide-significant genetic association with AD risk among X-chromosome common variants. Technical or analytical reasons can partly explain this result, such as: 1) overall lower variant density, 2) lower coverage by genotyping platforms, 3) lower call rate of variants, 4) lower imputation quality, or 5) a lower effective sample size in males on the X-chromosome compared to the autosomes^36^. However, it is also possible that fewer genome-wide significant associations of X-chromosome loci with AD risk exist than on autosomes due to a lower density of functional variants on the X-chromosome. Indeed, Gorlov et al., 2023^36^ found a lower density of variants in both exonic and intronic regions on the X-chromosome compared to autosomes, which they link to a stronger selection against X-chromosome mutations.

In conclusion, this XWAS found no common genetic risk factor for AD on the non-pseudoautosomal region of the X-chromosome but identified suggestive signals with moderate impact on AD risk, which warrant further investigations. In particular, future analyses of sequencing data will help to address some of the technical issues described above, and will further allow to study the impact of X-chromosome rare variants or structural variants on AD risk.

## Online Methods

### 1) Samples

The XWAS is based on 115,841 AD or AD-proxy cases (58% females) and 613,671 controls (55% females) of European ancestry from 35 case-control studies, 2 family studies (LOAD and FHS), and 2 biobanks (UKB and FinnGen) (Supplementary material and Supplementary Table S1). 55,868 of the 115,841 cases were AD-proxy cases. Females were considered as AD-proxy cases if they indicated having at least one parent with dementia^37^. For males, only the mother’s status was used to define the proxy status (Supplementary material).

In a sensitivity analysis including only the diagnosed AD cases, a total of 63,838 AD-cases (59% females) and 806,335 controls (55% females) was considered (Supplementary Table S1).

In addition to the classical autosomal QC, an X-chromosome specific QC was performed prior to imputation for each study (Supplementary material and Supplementary Table S5). We did not analyze the PAR regions due to a lack of variants on most genotyping chips. Related individuals were excluded from UKB samples but were kept in FinnGen, where related individuals’ exclusion accounts for about 40% of the sample size^38^.

Thirty-four studies were imputed with the TOPMed panel (N = 112,690) and three studies were imputed with the 1000 Genomes panel (March 2012) (FHS, CHS and RS, N = 10,102, Supplementary Table S5). The FinnGen was imputed with a Finnish reference panel and the UKB with a combination of 1000 Genomes, HRC and UK10K panels.

### 2) Main analyses

#### a. Association tests

Since random X-chromosome inactivation is the most frequent case, we considered the r-XCI approach for our main analysis and the s-XCI and e-XCI approaches for secondary analyses. The approaches are described briefly below, while additional details are provided in the Supplementary material. For all the models, the analyses were adjusted on the principal components (PCs) and/or the genotyping center if necessary (Supplementary Table S5). Dosage or genotype probabilities were used for all studies but FinnGen, where best guessed genotypes were considered (Supplementary material).

##### r-XCI approach

The r-XCI approach is equivalent to an additive genetic model, where males are considered as homozygous females. Males’ and females’ genotypes were thus coded: genotype (G) = {0, 2} and G = {0, 1, 2} respectively. The association test was performed for each study in men and women jointly using an additive logistic regression model for case-control studies, a generalized estimating equation (GEE) model for family studies and a logistic mixed model for biobanks. To account for differences in genotypic variance between sexes, we considered a robust estimate of the variance for case-control studies^39,40^ and an adjustment on sex for family studies and biobanks (Supplementary Table S6). The association test on proxy status in UKB was performed separately for males and females, and a correction factor of 2 was applied to the association statistics (effect sizes and standard errors) of the female-only model (Supplementary material)^37,41^. The results were then combined across studies in a fixed effect meta-analysis with an inverse-variance weighted approach with METAL^42^.

##### e-XCI approach

Under the e-XCI hypothesis, males’ and females’ genotypes were coded G = {0, 1} and G = {0, 1, 2} respectively. Variant effects were estimated separately in females and in males, except in FinnGen, where the variant effects were estimated directly in both males and females combined with an adjustment on sex (Supplementary Table S6). Results were then combined across studies, males and females with a fixed effect meta-analysis, inverse variance weighted approach using METAL. We did not include AD-proxy in the e-XCI meta-analysis. As males and females are related in family studies, only female results from LOAD and FHS were included in the meta-analysis. The sex-stratified models were adjusted on PCs and/or the genotyping center only, except for two ADGC studies (PFIZER and TGEN2) and the CHARGE studies (FHS, RS and CHS), where models were additionally adjusted on age (Supplementary Table S7 and Supplementary material).

##### s-XCI approach

For the skewed XCI approach, males’ and females’ genotypes were coded G = {0, 2} and G = {0, 1, 2} respectively. A general genotypic model, including both an additive and a dominance variable, was estimated in females from case-control studies to account for non-random inactivation through the dominance variable, which equals 1 in female heterozygotes, and 0 otherwise. The χ^2^ test of the dominance effect was then added to the χ^2^ test of the additive effect estimated under r-XCI, which results in a two degree of freedom (df) test of the association of the variant with AD risk including its potential skewedness^40,43^ (Supplementary Table S6). We did not include family studies and biobanks in the s-XCI approach.

While analyses and QC of the results (see below) were performed with the coding scheme described above, odds-ratio and confidence intervals are provided on the real XCI scale, i.e G = {0, 1} for males and G = {0, 0.5, 1} for females under r-XCI and s-XCI, but G = {0, 1} for males and G = {0, 1, 2} for females under e-XCI (Supplementary Table S6).

##### Sex-stratified analyses

We additionally performed a sex-stratified analysis per study, and we combined the results across studies in males and females separately with a fixed effect meta-analysis and inverse-variance weighted approach using METAL^42^. Proxy cases were not included in this analysis. We then compared the variant effect sizes of males and females with a Wald test (Supplementary material).

#### b. Quality control of the results and definition of associated loci

A QC of the results was carried out for all the studies. We filtered out variants with at least one missing datum (on effect, standard error, or p-value), an absolute effect size greater than 5, or an imputation quality less than 0.3. We also filtered out the variants whose effective allele count (product of the imputation quality and the expected minimum minor allele count between the cases and the controls) was less than 5, and less than 10 for LOAD ^44^. For datasets imputed with 1000G and the UKB, we excluded variants for which the conversion of position or alleles from GRCh37 to GRCh38 was not possible or problematic, and variants with a difference in frequency > 0.5 compared with the reference panels TOPMed or 1000G.

After the meta-analysis, we filtered rare variants (MAF < 1%), the variants analyzed in less than 40% of AD cases (considering the effective sample size of females UKB-proxy, which is the raw sample size divided by four^37^), variants with heterogeneity p-value < 5 x 10^-8^ and variants where the difference between the maximum frequency and the minimum frequency across studies was higher than 0.4.

Inflation of the test statistics was checked in each study and in the meta-analysis by computing a genomic inflation factor lambda with the median approach implemented in the GenABEL 1.8-0 R package^45^, on common variants in low LD (r^2^ < 0.2) (Supplementary material). A signal was considered genome-wide or X-chromosome-wide significant in either approach if associated with AD risk with P ≤ 5 × 10^−8^ or P ≤ 1.7 x 10^-6^. This X-chromosome wide threshold is based on R = 2.93%, the relative number of tests performed on the X-chromosome (n = 257,766) versus on the autosomes (n = 8,525,514) in the EADB-core study, the largest dataset imputed with the TOPMed reference panel. As the genome-wide threshold of 5x10^-8^ corresponds to the Bonferroni correction for one million tests, we computed the corresponding threshold for the X-chromosome as 0.05 / (R*1,000,000) = 1.7 x 10^-6^.

Several sensitivity analyses of the signals were performed. Sensitivity analyses excluding AD-proxy or biobank samples were performed for the r-XCI and e-XCI meta-analyses (samples from biobanks, including proxy, were not included in the s-XCI analysis in the first place). Additionally, for the r-XCI signals, an analysis adjusted on sex, without robust variance, was performed. The results were obtained by meta-analyzing the sex-stratified models for all case-control studies and UKB, and a sex-combined model adjusted on sex for FinnGen, with males coded as homozygous females for all models (family studies were excluded) (Supplementary material, Supplementary Table S7). Sensitivity analyses including an adjustment on age and the number of APOE*ε*4 and APOE*ε*2 alleles were also performed for all signals. Results were obtained from the meta-analysis of adjusted sex-stratified models with the adequate coding of males and excluding family studies. Finally, a sensitivity analysis was performed using a stricter imputation quality filter (r^2^ > 0.8).

## Data availability

Summary statistics will be made available upon publication through the European Bioinformatics Institute GWAS Catalog (https://www.ebi.ac.uk/gwas/).

## Acknowledgments

**EADB:** This study was supported by grants from the Fondation pour la Recherche sur Alzheimer (convention 2022-A-01 and cluster grant), and the JPco-fuND-2 ‘Multinational research projects on Personalized Medicine for Neurodegenerative Diseases’ PREADAPT project (ANR-19-JPW2-0004). We thank the many study participants, researchers and staff for collecting and contributing to the data, the high-performance computing service at the University of Lille and the staff at CEA-CNRGH for their help with sample preparation and genotyping and excellent technical assistance. We thank Antonio Pardinas for his help. We thank the Netherlands Brain Bank. This research was conducted using the UKBB resource (application number 61054). This work was funded by a grant (EADB) from the EU Joint Programme – Neurodegenerative Disease Research. Inserm UMR1167 is also funded by the Inserm, Institut Pasteur de Lille, Lille Métropole Communauté Urbaine and French government’s LABEX DISTALZ program (Development of Innovative Strategies for a Transdisciplinary Approach to ALZheimer’s disease). This work was also supported by the Research Council of Finland grants 338182 and 334802, the Sigrid Jusélius Foundation, and the Strategic Neuroscience Funding of the University of Eastern Finland.

Full consortium acknowledgements and funding are in the Supplementary Note.

## Notes

### Competing Interest Statement

The authors have declared no competing interest.

### Author Declarations

Written informed consent was obtained from study participants or, for those with substantial cognitive impairment, from a caregiver, legal guardian or other proxy. Study protocols for all cohorts were reviewed and approved by the appropriate institutional review boards. All necessary patient/participant consent has been obtained and the appropriate institutional forms have been archived.

## References

1. Lambert, J. C., Ramirez, A., Grenier-Boley, B. & Bellenguez, C. Step by step: towards a better understanding of the genetic architecture of Alzheimer’s disease. Mol. Psychiatry 28, 2716–2727 (2023).

2. Bellenguez, C. et al. New insights into the genetic etiology of Alzheimer’s disease and related dementias. Nat. Genet. 54, 412–436 (2022).

3. Holstege, H. et al. Exome sequencing identifies rare damaging variants in ATP8B4 and ABCA1 as risk factors for Alzheimer’s disease. Nat. Genet. 54, 1786–1794 (2022).

4. Kunkle, B. W. et al. Genetic meta-analysis of diagnosed Alzheimer’s disease identifies new risk loci and implicates Aβ, tau, immunity and lipid processing. Nat. Genet. 51, 414–430 (2019).

5. Wightman, D. P. et al. A genome-wide association study with 1,126,563 individuals identifies new risk loci for Alzheimer’s disease. Nat. Genet. 53, 1276–1282 (2021).

6. Sims, R. et al. Rare coding variants in PLCG2, ABI3, and TREM2 implicate microglial-mediated innate immunity in Alzheimer’s disease. Nat. Genet. 49, 1373–1384 (2017).

7. Lyon, M. F. Gene Action in the X-chromosom (Mus musculus L.). Nature 190, 372–373 (1961).

8. Carrel, L. & Brown, C. J. When the lyon(Ized chromosome) roars: Ongoing expression from an inactive X chromosome. Philos. Trans. R. Soc. B Biol. Sci. 372, (2017).

9. Amos-Landgraf, J. M. et al. X chromosome-inactivation patterns of 1,005 phenotypically unaffected females. Am. J. Hum. Genet. 79, 493–499 (2006).

10. Shvetsova, E. et al. Skewed X-inactivation is common in the general female population. Eur. J. Hum. Genet. 27, 455–465 (2019).

11. Zito, A. et al. Heritability of skewed X-inactivation in female twins is tissue-specific and associated with age. Nat. Commun. 10, 1–11 (2019).

12. Busque, L. et al. Nonrandom X-inactivation patterns in normal females: Lyonization ratios vary with age. Blood 88, 59–65 (1996).

13. Tukiainen, T. et al. Landscape of X chromosome inactivation across human tissues. Nature 550, 244–248 (2017).

14. Sun, L., Wang, Z., Lu, T., Manolio, T. A. & Paterson, A. D. eXclusionarY: 10 years later, where are the sex chromosomes in GWASs? Am. J. Hum. Genet. 110, 903–912 (2023).

15. Chen, B., Craiu, R. V., Strug, L. J. & Sun, L. The X factor: A robust and powerful approach to X-chromosome-inclusive whole-genome association studies. Genet. Epidemiol. 45, 694–709 (2021).

16. Smith, S. M. et al. An expanded set of genome-wide association studies of brain imaging phenotypes in UK Biobank. Nat. Neurosci. 24, 737–745 (2021).

17. Mallard, T. T. et al. X-chromosome influences on neuroanatomical variation in humans. Nat. Neurosci. 24, 1216–1224 (2021).

18. Neri, G., Schwartz, C. E., Lubs, H. A. & Stevenson, R. E. XLID Update 2017. Am. J. Med. Genet. A 176, 1375 (2018).

19. Hickman, R. A., O’Shea, S. A., Mehler, M. F. & Chung, W. K. Neurogenetic disorders across the lifespan: from aberrant development to degeneration. Nat. Rev. Neurol. 18, 117–124 (2022).

20. Alzheimer Europe. Dementia in Europe yearbook 2019: Estimating the prevalence of dementia in Europe. Alzheimer Eur. 108 (2019).

21. Nichols, E. et al. Estimation of the global prevalence of dementia in 2019 and forecasted prevalence in 2050: an analysis for the Global Burden of Disease Study 2019. Lancet Public Heal. 7, e105–e125 (2022).

22. Shaw, C. et al. Evaluation of Selective Survival and Sex/Gender Differences in Dementia Incidence Using a Simulation Model. JAMA Netw. open 4, e211001 (2021).

23. Ferretti, M. T. & Santuccione Chadha, A. The missing X factor in Alzheimer disease. Nat. Rev. Neurol. 17, 727–728 (2021).

24. Babapour Mofrad, R. et al. Sex differences in CSF biomarkers vary by Alzheimer disease stage and APOE ϵ4 genotype. Neurology vol. 95 (2020).

25. Hohman, T. J. et al. Sex-specific association of apolipoprotein e with cerebrospinal fluid levels of tau. JAMA Neurol. 75, 989–998 (2018).

26. Davis, E. J. et al. A second X chromosome contributes to resilience in a mouse model of Alzheimer’s disease. Sci. Transl. Med. 12, (2020).

27. Davis, E. J. et al. Sex-Specific Association of the X Chromosome with Cognitive Change and Tau Pathology in Aging and Alzheimer Disease. JAMA Neurol. 78, 1249–1254 (2021).

28. Yan, Y. et al. Article X-linked ubiquitin-specific peptidase 11 increases tauopathy vulnerability in women increases tauopathy vulnerability in women. Cell 1–18 (2022) doi:10.1016/j.cell.2022.09.002.

29. Lee, H. W. et al. Preso, a novel PSD-95-interacting FERM and PDZ domain protein that regulates dendritic spine morphogenesis. J. Neurosci. 28, 14546–14556 (2008).

30. Piard, J. et al. FRMPD4 mutations cause X-linked intellectual disability and disrupt dendritic spine morphogenesis. Hum. Mol. Genet. 27, 589–600 (2018).

31. Okbay, A. et al. Polygenic prediction of educational attainment within and between families from genome-wide association analyses in 3 million individuals. Nat. Genet. 54, 437–449 (2022).

32. Jansen, I. E. et al. Genome-wide meta-analysis identifies new loci and functional pathways influencing Alzheimer’s disease risk. Nat. Genet. 51, 404–413 (2019).

33. Smith, E. R. et al. A Human Protein Complex Homologous to the Drosophila MSL Complex Is Responsible for the Majority of Histone H4 Acetylation at Lysine 16. Mol. Cell. Biol. 25, 9175–9188 (2005).

34. Hayward, G. C. et al. Characterization of Alzheimer’s disease-like neuropathology in Duchenne’s muscular dystrophy using the DBA/2J mdx mouse model. FEBS Open Bio 12, 154–162 (2022).

35. Wang, K.-W. et al. X chromosome-wide association study of quantitative biomarkers from the Alzheimer’s Disease Neuroimaging Initiative study. Front. Aging Neurosci. 15, 1–15 (2023).

36. Gorlov, I. P. & Amos, C. I. Why does the X chromosome lag behind autosomes in GWAS findings? PLoS Genet. 19, 1–19 (2023).

37. Liu, J. Z., Erlich, Y. & Pickrell, J. K. Case – control association mapping by proxy using family history of disease. Nat. Publ. Gr. 49, (2017).

38. Kurki, M. I., Karjalainen, J., Palta, P., Sipilä, T. P. & Kristiansson, K. FinnGen: Unique genetic insights from combining isolated population and national health register data. 1–56 (2022).

39. Clayton, D. G. Testing for association on the X chromosome. Biostatistics 9, 593–600 (2008).

40. Clayton, D. G. Sex chromosomes and genetic association studies. Genome Med. 1, 1–7 (2009).

41. Ghosh, A. et al. Leveraging Family History in Population-Based Case-Control Association Studies. Genet Epidemiol. 38, 114–122 (2014).

42. Willer, C. J., Li, Y. & Abecasis, G. R. METAL: Fast and efficient meta-analysis of genomewide association scans. Bioinformatics 26, 2190–2191 (2010).

43. Magi, R., Lindgren, C. M. & Morris, A. P. Meta-analysis of sex-specific genome-wide association studies. Genet. Epidemiol. 34, 846–853 (2010).

44. Satizabal, Claudia L., Adams, H. H., Hibar, D. P. & White, C. C. Genetic architecture of subcortical brain structures in 38,851 individuals. Physiol. Behav. 176, 139–148 (2019).

45. Aulchenko, Y. S., Ripke, S., Isaacs, A. & van Duijn, C. M. GenABEL: An R library for genome-wide association analysis. Bioinformatics 23, 1294–1296 (2007).

