## Supplementary Material for "X-Chromosome-wide association study for Alzheimer’s disease"

A. Samples

B. X-chromosome Quality control

C. Imputation

D. Covariate definition

E. Association Tests

F. Supplementary analyses

G. FinnGen Banner Authors

H. Supplementary acknowledgments

I. Supplementary Figures

J. Supplementary references

#### A. Samples

The EADB studies (EADI, Bonn, DemGene, GR@ACE/DEGESCO, EADB-core and GERAD) are described in more details in (Bellenguez et al., 2022).

***European Alzheimer's Disease Initiative (EADI) Consortium***

EADI is composed of several case-control studies and one population-based cohort, the 3C study (Lambert et al., 2009). Case-control studies are comprised of AD cases and cognitively normal controls across France. 3C Study is a population-based, prospective study of the relationship between vascular factors and dementia carried out in three French cities: Bordeaux, Montpellier, and Dijon. The AD status was then defined based on 12, 14-15 and 17-18 years follow-up for Dijon, Montpellier, and Bordeaux participants, respectively. The AD cases from 3C were included as cases in the EADI discovery dataset and the other individuals were retained as controls. All AD cases from EADI were clinically diagnosed of probable AD by neurologists according to the DSM-III-R and NINCDS-ADRDA criteria. Samples that passed DNA quality control were genotyped with Illumina Human 610-Quad BeadChips.

**Genetic and Environmental Risk in AD (GERAD) Consortium/Defining Genetic, Polygenic, and Environmental Risk for Alzheimer's Disease (PERADES) Consortium**

The GERAD/PERADES sample comprises 3,177 Alzheimer's disease cases and 7,277 controls with available age and sex data (Harold et al., 2009). Cases and elderly screened controls were recruited by several institutions in the United Kingdom and in the United States of America. 6129 population controls were drawn from large existing cohorts with available GWAS data, including the 1958 British Birth Cohort (1958BC) (http://www.b58cgene.sgul.ac.uk), the KORA F4 Study and the Heinz Nixdorf Recall Study. All Alzheimer's disease cases met criteria for either probable (NINCDS-ADRDA, DSM-IV) or definite (CERAD) Alzheimer's disease. All elderly controls were screened for dementia using the MMSE or ADAS-cog, and determined to be free from dementia at neuropathological examination or had a Braak score of 2.5 or lower. Genotypes from all cases and 4,617 controls were previously included in the AD GWAS by Harold and colleagues. Genotypes for the remaining 2,660 population controls were obtained from WTCCC2.

**The Norwegian DemGene Network**

This is a Norwegian network of clinical sites collecting cases from memory clinics based on a standardized examination of cognitive, functional, and behavioral measures and data on the progression of most patients. The Norwegian DemGene Network includes 2,224 cases and 3,089 healthy controls from different studies described elsewhere (Jansen et al., 2019). The cases were diagnosed according to recommendations from the NIA–AA, the NINCDS–ADRDA criteria, or the ICD-10 research criteria. The controls were screened with a standardized interview and cognitive tests. Additional controls from blood donors of the Oslo University Hospital, Ulleval Hospital, were included (n=4992, age between 18-65 years, 48% female). They were thoroughly screened for diseases and medication, and provided blood for DNA analysis, in line with approval from the Regional Committee for Medical and Health Research Ethics. Individuals from the DemGene study and blood donors were genotyped using either the Human Omni Express-24 v1.1 chip (Illumina Inc., San Diego, CA) or the DeCodeGenetics_V1_20012591_A1 chip at deCODE Genetics (Reykjavik, Iceland).

**<u>Bonn studies</u>**

<u>DietBB:</u> The DietBB sample included in this GWAS is a subsample extracted from the German study on aging, cognition and dementia (AgeCoDe) (Jessen et al., 2014; Luck et al., 2007) cohort, a general practice (GP) registry-based longitudinal study in elderly individuals. The DietBB samples has genome-wide genotype data which was included in this study. Participants were recruited in six German cities (Bonn, Dusseldorf, Hamburg, Leipzig, Mannheim, and Munich) with a total of 138 GPs connected to the study sites. The inclusion criteria for this study were an age of 75 years and older, absence of dementia according to GP judgment, and at least one contact with the GP within the past 12 months. Dementia was diagnosed according to the criteria set of DSM-IV in a consensus conference with the interviewer and an experienced geriatrician or geriatric psychiatrist. The etiological diagnosis of dementia in AD was established according to the NINCDS-ADRDA criteria for probable AD. Mixed dementia and dementia in AD were combined. If the information provided was sufficient to judge etiology, dementia diagnosis in subjects who were not interviewed personally was based on the Global Deterioration Scale 32 (score ≥4 points). Cohort participants were included if they were dementia-free at baseline. This criterion led to the selection of 320 participants. In 120 of these participants, dementia of the AD-type occurred at any follow up. The additional 200, free of dementia until last follow up of AgeCoDe, are included as controls.

<u>Bonn OMNI cohort:</u> the Bonn OMNI cohort consists of AD patients and controls derived from a larger German GWAS cohort which was recruited from the following sources: (i) the German Dementia Competence Network (DCN); (ii) AgeCoDe (described above); (iii) the interdisciplinary Memory Clinic at the University Hospital of Bonn; and (iv) Heinz Nixdorf Recall (HNR) study cohort, for the controls.

*The DCN:* The DCN cohort includes 1,095 patients with mild cognitive impairment (MCI) and 648 cases with mild Alzheimer's disease (AD) clinical dementia syndrome that were recruited from 14 university hospital memory clinics across Germany between 2003 and 2005 (Kornhuber et al., 2009). The diagnosis of mild dementia was set according to ICD-10 criteria. These changes must have persisted for at least 3 months. The etiological diagnosis of AD was assigned according to NINCDS-ADRDA criteria.

*Memory clinic Bonn:* The interdisciplinary Memory Clinic of the Department of Psychiatry and Department of Neurology at the University Hospital in Bonn provided further patients. Diagnoses were assigned according the NINCDS/ADRDA criteria and on the basis of clinical history, physical examination, neuropsychological testing (using the CERAD neuropsychological battery, including the MMSE), laboratory assessments, and brain imaging.

*Control samples:* The control samples were obtained from the population-based study, HNR study cohort (Schmermund et al., 2002; Stang et al., 2005). This sample was previously used for replication in Lambert et al. Briefly, 4814 participants aged 45 to 75 years were enrolled between 2000 and 2003. Cognitive performance of participants was evaluated at follow up 5 years and 10 years after baseline. Controls sample was selected if participant did not present cognitive impairment as reported at the last available evaluation.

**<u>GR@ACE/DEGESCO</u>**

The GR@ACE study (Moreno-Grau et al., 2019, de Rojas et al., 2021) recruited Alzheimer's disease (AD) patients from Fundació ACE, Institut Català de Neurociències Aplicades (Catalonia, Spain), and control individuals from three centers: Fundació ACE (Barcelona, Spain), Valme University Hospital (Seville, Spain), and the Spanish National DNA Bank–Carlos III (University of Salamanca, Spain) (http://www.bancoadn.org). Additional cases and controls were obtained from dementia cohorts included in the Dementia Genetics Spanish Consortium (DEGESCO) (Ruiz et al., 2014). At all sites, AD diagnosis was established by a multidisciplinary working group—including neurologists, neuropsychologists, and social workers—according to the DSM-IV criteria for dementia and the National Institute on Aging and Alzheimer's Association's (NIA–AA) 2011 guidelines for diagnosing AD. In our study, we considered as AD cases any individuals with dementia diagnosed with probable or possible AD at any point in their clinical course. Genotyping was conducted using the Axiom 815K Spanish biobank array (Thermo Fisher) at the Spanish National Centre for Genotyping (CeGEN, Santiago de Compostela, Spain). The genotyping array not only is an adaptation of the Axiom biobank genotyping array but also contains rare population-specific variations observed in the Spanish population.

**<u>The European Alzheimer's Disease DNA Biobank dataset (EADB)</u>**

This consortium groups together 20,464 Alzheimer's disease (AD) cases and 22,244 controls after quality controls from 16 European countries (Austria, Belgium, Bulgaria, Czech Republic, Denmark, Finland, France, Germany, Greece, Italy, Portugal, Spain, Sweden, Switzerland, The Netherlands and the UK). These samples were genotyped using the

ILLUMINA GSA array in three independent centers (France, Germany and the Netherlands) leading to define three nodes: EADB-France, EADB-Germany and EADB-Netherlands.

**EADB-France**

In the France node, samples were collected from nine countries (39 centers/studies), and after quality controls (QCs), we obtained 13,867 AD cases and 15,310 controls. All these samples were genotyped at the Centre National de Recherche en Génomique Humaine (CNRGH, Evry, France).

Belgium: The participants were part of a large prospective cohort (De Roeck et al., 2018) of Belgian AD patients and healthy elderly control individuals. The patients were ascertained at the memory clinic of Middelheim and Hoge Beuken (Hospital Network Antwerp, Belgium) and at the memory clinic of the University Hospitals of Leuven, Belgium. The control individuals were the partners of the patients or volunteers from the Belgian community. The study protocols were approved by the ethics committees of the Antwerp University Hospital and the participating neurological centers at the different hospitals of the BELNEU consortium and by the University of Antwerp.

Czech Republic: The Czech Brain Aging Study (CBAS) (Sheardova et al., 2019) is a longitudinal memory- clinic–based study recruiting subjects at risk of dementia (subjects referred for cognitive complaints-SCD, MCI). The CBAS+ study is a cross-sectional study of patients in the early stages of dementia. All subjects signed informed consent and both studies were approved by the local ethics committee.

Denmark: The Copenhagen General Population Study (CGPS) is a prospective study

of the Danish general population initiated in 2003 and still recruiting. Individuals were selected randomly based on the national Danish Civil Registration System to reflect the adult Danish population aged 20-100. Data were obtained from a self-administered questionnaire reviewed together with an investigator at the day of attendance, a physical examination, and from blood samples including DNA extraction.

Finland: The ADGEN cohort (Steinberg et al., 2015): a clinic-based collection of AD patients from Eastern and Northern Finland examined in the Department of Neurology in Kuopio University Hospital and the Department of Neurology in Oulu University Hospital. All the patients were diagnosed with probable AD according to the criteria of the National Institute of Neurological and Communicative Disorders and Stroke and the Alzheimer's disease and Related Disorders Association (NINCDS–ADRDA). The study was approved by the ethics committee of Kuopio University Hospital, Finland (420/2016). The FINGER study (Ngandu et al., 2015): a Finnish multidomain lifestyle RCT enrolling 1,260 older adults with an increased risk of dementia from the general population. The intensive lifestyle intervention lasted for two years, and follow-up extends currently up to seven years. The FINGER study was approved by the coordinating ethics committee of the Hospital District of Helsinki and Uusimaa (94/13/03/00/2009 and HUS/1204/2017), and all the participants gave written informed consent.

France: The BALTAZAR multicenter (23 memory centers) prospective study (Hanon et al., 2018): 1,040 participants from September 2010 to April 2015. They were classified as AD cases (n = 501) according to DSM IV-TR and NINCDS–ADRDA criteria as well as amnestic mild cognitive impairment (MCI) cases (a MCI, n = 417) and non-amnestic MCI cases (na MCI, n = 122) according to Petersen's criteria. A comprehensive battery of cognitive tests was performed, including MMSE, verbal fluency, and FCSRT. All the participants or their legal guardians gave written informed consent. The study was approved by the Paris ethics committee (CPP Ile de France IV Saint Louis Hospital). MEMENTO: a clinic-based study (Dufouil et al., 2017) aimed at better understanding the natural history of AD, dementia, and related diseases. Between 2011 and 2014, 2,323 individuals presenting either recently diagnosed MCI or isolated cognitive complaints were enrolled in 26 memory centers in France. This study was performed in accordance with the guidelines of the Declaration of Helsinki. The MEMENTO study protocol has been approved by the local ethics committee (Comité de Protection des Personnes Sud-Ouest et Outre Mer III; approval number 2010-A01394-35). All the participants provided written informed consent. The CNRMAJ-Rouen study (Nicolas et al., 2016): early onset AD patients (n = 870). The patients or their legal guardians provided written informed consent. This study was approved by the ethics committee of CPP Ile de France II.

Italy: The AD cases and controls were collated through Italy in different centers: Brescia, Cagliari, Florence, Milan, Rome, Pertugia, San Giovani Rotondo and Torino. AD cases were diagnosed according to DSM III-R,IV and NINCDS–ADRDA criteria. Controls were defined a minima as subjects without DMS-III-R dementia criteria and with integrity of their cognitive functions (MMS>25).

Spain: The Dementia Genetic Spanish Consortium (DEGESCO) is a national consortium comprising 23 research centers and hospitals across the country, that holds the institutional coverage of The Network Center for Biomedical Research in Neurodegenerative Diseases (CIBERNED). Created in 2013, DEGESCO's objective is the promotion and conduction of genetic studies aimed at understanding the genetic architecture of neurodegenerative dementias in the Spanish population and participates in coordinated actions in national and international frameworks. All DNA samples are in compliance with the Law of Biomedical Research (Law 14/2007) and the Royal Decree on Biobanks (RD 1716/2011). Patients included in the present study met clinical criteria for probable or possible disease established by the National Institute of Neurological and Communication Disorders and Stroke and the Alzheimer Disease and Related Disorders Association (NINCDS-

ADRDA). Cognitively healthy controls were unrelated individuals who had a documented MMSE in the normal range. Contributing centers in the France node genotyping were Centro de Biología Molecular Severo Ochoa (CSIC-UAM (Madrid), the Institute Biodonostia, University of Basque Contry (EHU-UPV, San Sebastián), Institut de Biomedicina de Valencia CSIC (València), and Sant Pau Biomedical Research Institute (Barcelona).

Sweden: Upsala. The Swedish AD patients were ascertained at the Memory Disorder Unit at Uppsala University Hospital. For all patients, the diagnosis was established according to the National Institute on Neurological Disorders and Stroke, and the Alzheimer's Disease and Related Disorders Association (NINDS-ADRDA) guidelines. (G. McKhann et al., 1984). Healthy control subjects were recruited from the same geographic region following advertisements in local newspapers and displayed no signs of dementia upon Mini Mental State Examination (MMSE). Swedish National Study on Aging and Care in Kungsholmen (SNAC-K) data was collected. The original SNAC-K population consisted of 4590 living and eligible persons who lived on the island of Kungsholmen in Central Stockholm, belonged to pre-specified age strata, and were randomly selected to take part in the study. Between 2001 and 2004, 3363 persons participated in the baseline assessment. They belonged to the age cohorts 60, 66,72, 78, 81, 84, 87, 90, 93, and 96 years and 99 years and older. The examination consists of three parts: a nurse interview, a medical examination, and a neuropsychological testing session. Altogether, the examination takes about six hours. The participants are re-examined each time they reach the next age cohort. All parts of the SNAC-K project have been approved by the ethical committee at Karolinska Institutet or the regional ethical review board. Informed consent was collected from all the participants or, if the person was severely cognitively impaired, from their next of kin.

The UK: MRC. The sample set comprises individuals with AD and healthy controls recruited across the MRC Centre for Neuropsychiatric Genetics and Genomics, Cardiff University, Cardiff, UK; Institute of Psychiatry, London, UK; University of Cambridge, Cambridge, UK. The collection of the samples was through multiple channels, including specialist NHS services and clinics, research registers and Join Dementia Research (JDR) platform. The participants were assessed at home or in research clinics along with an informant, usually a spouse, family member or close friend, who provided information about and on behalf of the individual with dementia. Established measures were used to ascertain the disease severity: Bristol activities of daily living (BADL), Clinical Dementia Rating scale (CDR), Neuropsychiatric Inventory (NPI) and Global Deterioration Scale (GlDS). Individuals with dementia completed the Addenbrooke's Cognitive Examination (ACE-r), Geriatric Depression Scale (GeDS) and National Adult Reading Test (NART) too. Control participants were recruited from GP surgeries and by means of self-referral (including existing studies and Joint Dementia Research platform). For all other recruitment, all AD cases met criteria for either probable (NINCDS-ADRDA, DSM-IV) or definite (CERAD) AD. All elderly controls were screened for dementia using the Mini Mental State Examination

(MMSE) or ADAS-cog, were determined to be free from dementia at neuropathological examination or had a Braak score of 2.5 or lower. Control samples were chosen to match case samples for age, gender,

ethnicity and country of origin. Informed consent was obtained for all study participants, and the relevant independent ethical committees approved study protocols. SOTON, University of Southampton, Southampton, UK. All AD cases met criteria for either probable (NINCDS-ADRDA, DSM-IV) or definite (CERAD) AD. All elderly controls were screened for dementia using the MMSE or ADAS-cog, were determined to be free from dementia at neuropathological examination or had a Braak score of 2.5 or lower. Nottingham and Manchester, University of Nottingham, Nottingham, UK and Manchester Brain Bank. All AD cases met criteria for either probable (NINCDS-ADRDA, DSM-IV) or definite (CERAD) AD. All elderly controls were screened for dementia using the MMSE or ADAS-cog, were determined to be free from dementia at neuropathological examination or had a Braak score of 2.5 or lower. KCL, London Neurodegenerative Diseases Brain Bank. All AD cases met criteria for either probable (NINCDS-ADRDA, DSM-IV) or definite (CERAD) AD. All elderly controls were screened for dementia using the MMSE or ADAS-cog, were determined to be free from dementia at neuropathological examination or had a Braak score of 2.5 or lower. PRION, All AD cases met criteria for either probable (NINCDS-ADRDA, DSM-IV) or definite (CERAD) AD. All elderly controls were screened for dementia using the MMSE or ADAS-cog, were determined to be free from dementia at neuropathological examination or had a Braak score of 2.5 or lower. CFAS Wales, The Cognitive Function and Ageing Study Wales (CFAS-Wales) is a longitudinal population-based study of people aged 65 years and over in rural and urban areas of Wales that aims to investigate physical and cognitive health in older age and examine the interactions between health, social networks, activity, and participation. Individuals aged 65 years and over were randomly sampled from general medical practice lists between 2011 and 2013, stratified by age to ensure equal numbers in two age groups, 65-74 years and 75 and over. The baseline sample included 3593 older people and included those living in care homes as well as those living at home. Those who provided written consent to join the study were interviewed in their own homes by trained interviewers and could choose to have the interview conducted through the medium of either English or Welsh. Participants were followed up 2 years later. All AD cases met criteria for either probable (NINCDS-ADRDA, DSM-IV) or definite (CERAD) AD. All elderly controls were screened for dementia using the MMSE or CAMCOG, and were determined to be free from dementia. UCL-DRC. the UCL Alzheimer’s disease cohort of the Dementia Research Centre (UCL - EOAD DRC) included patients seen at the Cognitive Disorders Clinics at The National Hospital for Neurology and Neurosurgery (Queen Square), or affiliated hospitals. Individuals were assessed clinically and diagnosed as having probable Alzheimer’s disease based on contemporary clinical criteria in use at the time, including imaging and neuropsychological testing where appropriate.

**EADB-Germany**

In the German node, samples were collected from seven countries (11 centers/studies) and after QCs, we obtained 4,159 AD cases and 4,545 controls. All these samples were genotyped at Life&brain (Bonn, Germany).

Germany: DELCODE (the multicenter DZNE-Longitudinal Cognitive Impairment and Dementia Study). This is an observational longitudinal memory clinic-based multicenter study in Germany comprising 400 subjects with Subjective cognitive decline (SCD), 200 mild cognitive impairment (MCI) patients, 100 AD dementia patients, 200 control subjects without subjective or objective cognitive decline, and 100 first-degree relatives of patients with a documented diagnosis of AD dementia. All patient groups (SCD, MCI, AD) are referrals, including self-referrals, to the participating memory centers. The control group and the relatives of AD dementia patients are recruited by standardized public advertisement. Ten university-based memory centers are participating, all being collaborators of local DZNE sites. All patient groups (SCD, MCI, AD) were assessed clinically at the respective memory centers before entering DELCODE. The assessments include medical history, psychiatric and neurological examination, neuropsychological testing, blood laboratory work-up, cerebrospinal fluid (CSF) biomarkers, and routine MRI, all according to the local standards. The Consortium to Establish a Registry for Alzheimer's Disease (CERAD) neuropsychological test battery was applied at all memory centers to measure cognitive function. German age, sex, and education-adjusted norms of the CERAD neuropsychological battery are available online (www.memoryclinic.ch). Detail description is of recruitment protocol is reported elsewhere. The VOGEL study: The VOGEL study is a prospective, observational, long-term follow-up study with three time points of investigation within 6–8 years. This cohort includes dementia and healthy subjects. Residents of the city of Würzburg born between 1936 and 1941 were recruited. Every participant underwent physical, psychiatric, and laboratory examinations and performed intense neuropsychological testing as well as VSEP and NIRS according to the published procedures. A total of 604 subjects were included. The Heidelberg/Mannheim memory clinic sample: This cohort includes 61 subjects from whom 40 MCI patients were recruited and assessed between 2012 and 2016. Some of those patients converted to dementia by AD or other dementias. The PAGES study: This study includes 301 subjects. AD patients were recruited at the memory clinic of the Department of Psychiatry, University of Munich, Germany. Participants in whom dementia associated with AD was diagnosed fulfilled the criteria for probable AD according to the NINCDS–ADRDA. The control group included participants who were randomly selected from the general population of Munich. Controls who had central nervous system diseases or psychotic disorders or who had first-degree relatives with psychotic disorders were excluded. The Technische Universität München study: This cohort includes 359 healthy, AD, and other dementias patients recruited from the Centre for Cognitive Disorders. All the participants provided written informed consent. A biobank was submitted to the ethics committee of the Technical University of Munich, School of Medicine (Munich, Germany), which raised no objections and approved the biobank (reference number 347-14). The Göttingen Universität study: This study includes 111 in- and outpatients with a healthy or AD dementia status from the Department of Psychiatry of the University of Göttingen. The study's ethical statement was provided locally at the Göttingen University Medical Centre. The German Dementia Competence Network (DCN) cohort: Individuals from the DCN cohort were recruited from university hospital memory clinics across Germany between 2003 and 2005 (Kornhuber et al., 2009). The study was approved by the respective ethics committees, and written informed consent was obtained from all the participants prior to inclusion. The German Study on Aging, Cognition, and Dementia (AgeCoDe): The AgeCoDe study is a general practice (GP) registry-based longitudinal study in elderly individuals that recruited patients aged 75 years and above in six German cities from 2003 to 2004 (Luck et al., 2007). The study was approved by the respective ethics committees, and written informed consent was obtained from all the participants prior to inclusion.

Greece: the HELIAD study, comprising 49 AD cases and 1,150 controls. HELIAD is a population-based, multidisciplinary, collaborative study designed to estimate, in the Greek population over the age of 64 years, the prevalence and incidence of MCI, AD, other forms of dementia, and other neuropsychiatric conditions of aging and to investigate associations between nutrition and cognitive dysfunction or age-related neuropsychiatric diseases. The participants were selected through random sampling from the records of two Greek municipalities, Larissa and Marousi. All the participants signed informed consent in Greek. Portugal: the Lisbon study from Portugal, totaling 78 AD cases and 74 controls. This cohort was recruited in 2008–2009 to investigate the connections between oxidative stress and lipid dyshomeostasis in AD. The project includes 190 subjects and was approved by the local ethics committee, and all the participants provided written informed consent. This study includes healthy and dementia-by-AD subjects.

Spain: Those samples are part of DEGESCO. DEGESCO Centers from whom DNA samples were genotyped in the German node (1,778 cases and 470 controls) were the Alzheimer Research Center and Memory Clinic, Fundació ACE, Institut Català de Neurociències Aplicades (Barcelona), the Neurology Service at University Hospital Marqués de Valdecilla (Santander), the Alzheimer's disease and other cognitive disorders, Neurology Department, at Hospital Clínic, IDIBAPS (Barcelona), the Molecular Genetics Laboratory, at the Hospital Universitario Central de Asturias (Oviedo), and Fundació Docència i Recerca Mútua de Terrassa and Movement Disorders Unit, Department of Neurology, University Hospital Mútua de Terrassa (Barcelona).

Switzerland: Two datasets from Switzerland and Austria were combined, totaling 182 AD cases and 388 controls. The Lausanne study: This study includes 137 community-dwelling participants aged 55+ years with cognitive impairment (memory clinic patients with

MCI, dementia) or normal cognition (recruited by advertisement, word of mouth). The study's ethical statement was provided locally at the Department of Psychiatry, Geneva University Centre, Switzerland. The VITA study: This is a longitudinal study of 606 individuals (Vienna, Austria) who were 75 years old in 2000, followed up every 30–90 months. This cohort includes dementia and healthy subjects. All the participants gave written informed consent. The study conformed to the latest version of the Declaration of Helsinki and was approved by the ethics committee of the City of Vienna, Austria.

**EADB-Netherlands**

In the Dutch node, samples were collected from six organizations in the Netherlands and after QCs, we obtained 2,438 AD cases and 2,389 controls. All these samples were genotyped at the Erasmus Medical University (Rotterdam, The Netherlands). The Medical Ethics Committee (METC) of the local institutes approved the studies. All the participants and/or their legal guardians gave written informed consent for participation in the clinical and genetic studies. Samples from the following institutes were included. 1) Erasmus Medical Center: most individuals were selected from population studies from the epidemiology department and accounted for most of the controls, while a smaller subset of samples originated from the neurology department, where AD was diagnosed according to the National Institute of Neurological and Communicative Disorders and Stroke-Alzheimer's Disease and Related Disorders Association (NINCDS-ADRDA) criteria for AD (G. M. McKhann et al., 2011). 2) The Amsterdam Dementia Cohort (ADC) (Van Der Flier & Scheltens, 2018): This cohort comprises patients who visit the memory clinic of the VU University Medical Centre, the Netherlands. The diagnosis of probable AD is based on the clinical criteria formulated by the NINCDS–ADRDA and based on the NIA–AA. Diagnosis of MCI was made according to Petersen and NIA-AA. Controls presented with subjective cognitive decline at the memory clinic, but performed within normal limits on all clinical investigations. 3) The 100-Plus study: This study includes Dutch-speaking individuals who (i) can provide official evidence for being aged 100 years or older, (ii) self-report to be cognitively healthy, which is confirmed by a proxy, (iii) consent to the donation of a blood sample, (iv) consent to (at least) two home visits from a researcher, and (v) consent to undergo an interview and neuropsychological test battery (Holstege et al., 2018). 4) Parelsnoer Institute: a collaboration between 8 Dutch University Medical Centers in which clinical data and biomaterials from patients suffering from chronic diseases (so called "Pearls") are collected according to harmonized protocols. The Pearl Neurodegenerative Diseases (Aalten et al., 2014) includes individuals diagnosed with dementia, mild cognitive impairment, and controls with subjective memory complaints. 5) The Netherlands Brain Bank: a non-profit organization that collects human brain tissue of donors with a variety of neurological and psychiatric disorders, but also of non-diseased donors. A clinical diagnosis of AD is based on the clinical criteria of probable AD (Dubois et al., 2007; G. M. McKhann et al., 2011). The selected AD patients for this study all received a definitive diagnosis which was based on autopsy. 6) Maastricht University Medical Center: a subset of individuals that were referred to the memory clinic for cognitive complaints were included if they participated in the BioBank-Alzheimer Centrum Limburg (BB-ACL). Diagnosis of MCI was made according to the criteria of Petersen, and diagnosis of AD-type dementia was made according to the criteria of the DSM-4, and the NINCDS-ADRDA (G. M. McKhann et al., 2011). The Alzheimer Center Amsterdam is supported by Stichting Alzheimer Nederland and Stichting VUmc fonds. The clinical database structure was developed with funding from Stichting Dioraphte. Genotyping of the Dutch case-control samples was performed in the context of EADB (European Alzheimer DNA biobank) funded by the JPco-fuND FP-829-029 (ZonMW projectnumber 733051061).

**Alzheimer's Disease Genetics Consortium (ADGC)**

The ADGC dataset comprises subjects from 35 datasets including two waves of the Adult Changes in Thought (ACT) cohort study [ACT1/ACT2]; ten waves of cases and cognitively normal controls from the National Institute on Aging (NIA) Alzheimer Disease Centers (ADCs); the Alzheimer Disease Neuroimaging Initiative (ADNI); the Biomarkers of Cognitive Decline Among Normal Individuals (BIOCARD) Cohort; two waves of the Religious Orders Study/Memory and Aging Project (ROSMAP1-2) and the Chicago Health and Aging Project (CHAP) cohort studies at Rush University; the Einstein Aging Study (EAS); the Multi-Site Collaborative Study for Genotype-Phenotype Associations in Alzheimer's Disease (GenADA) Study by GlaxoSmithKline; Mayo Clinic Jacksonville (MAYO) and Rochester (RMAYO) case-control datasets; the Multi-Institutional Research in Alzheimer's Genetic Epidemiology (MIRAGE) study; the NIA Late-Onset Alzheimer's Disease (LOAD) Family Study (NIA-LOAD); the Netherlands Brain Bank (NBB) case-control dataset; the Oregon Health and Science University (OHSU) case-control dataset; the Pfizer case-control dataset; the Texas Alzheimer's Research and Care Consortium (TARCC) dataset; the Translational Genomics Research Institute series 2 (TGEN2) dataset; the University of Miami (UM)/ Case Western Reserve University (CWRU)/ Mt. Sinai School of Medicine (MSSM) and UM/CWRU/TARCC wave 2 datasets [UM/CWRU/MSSM and UM/CWRU/TARCC2]; the Universitatsklinikum Saarlandes (UKS) case-control dataset; the University of Pittsburgh (UPITT) case-control dataset; Washington University (WASHU) wave 1 and 2 case-control datasets [WASHU1/WASHU2]; and the Washington Heights-Inwood Community Aging Project (WHICAP) study datasets.

Descriptions of the ACT1, ADC waves 1-7, ADNI, BIOCARD, CHAP, EAS, GenADA, MAYO, MIRAGE, NBB, NIA-LOAD, OHSU, PFIZER, RMAYO, ROSMAP1, ROSMAP2, TARCC, TGEN2, UKS, UM/CWRU/MSSM, UM/CWRU/TARCC2, UPITT, WASHU1, WASHU2, and WHICAP cohorts have been provided in previous ADGC and IGAP studies (G Jun et al., 2016; Gyungah Jun et al., 2010; Kunkle et al., 2019; Lambert et al., 2013; Naj et al., 2011; Sims et al., 2017). Here we update descriptions of these studies, where applicable, and provide descriptions for ACT2, ADC wave 8-10. All analyses were restricted to individuals of European ancestry. All subjects were recruited under protocols approved by the appropriate Institutional Review Boards (IRBs). BIOCARD, CHAP, EAS, NBB, RMAYO, ROSMAP2, WASHU2 and WHICAP were not included in the XWAS because they had less than 50 samples in at least one subgroup defined by sex and AD-status.

ACT1/ACT2: The ACT cohort is an urban and suburban elderly population from a stable HMO that includes 2,581 cognitively intact subjects age ≥ 65 who were enrolled between 1994 and 1998 (Kukull et al., 2002; Larson et al., 2006). An additional 811 subjects were enrolled in 2000-2002 using the same methods except oversampling clinics with more minorities. More recently, a Continuous Enrollment strategy was initiated in which new subjects are contacted, screened, and enrolled to keep 2,000 active at-risk person-years accruing in each calendar year. This resulted in an enrollment of 4,146 participants as of May 2009. All clinical data are reviewed at a consensus conference. Dementia onset is assigned half-way between the prior biennial and the exam that diagnosed dementia. A waiver of consent was obtained from the IRB to enroll deceased ACT participants. In total, ACT contributed data on 553 individuals with probable or possible Alzheimer's disease (70 with autopsy-confirmation) and on 1,579 cognitively normal elders (CNEs, 155 with autopsy-confirmation) who were included in the analyses, with 2,103 cases/1,571 CNEs in the first wave (ACT1) and 29 cases/8 CNEs in the second wave (ACT2).

NIA ADC Samples (ADC1-10): The NIA ADC cohort included subjects ascertained and evaluated by the clinical and neuropathology cores of the 32 NIA-funded ADCs. Data collection is coordinated by the National Alzheimer's Coordinating Center (NACC). NACC coordinates collection of phenotype data from the 32 ADCs, cleans all data, coordinates implementation of definitions of Alzheimer's disease cases and controls, and coordinates collection of samples. The complete ADC cohort consists of 3,311 autopsy-confirmed and 2,889 clinically-confirmed Alzheimer's disease cases, and 247 cognitively normal elders (CNEs) with complete neuropathology data who were older than 60 years at age of death, and 3,687 living CNEs evaluated using the Uniform dataset (UDS) protocol (Beekly et al., 2007; Morris et al., 2006) who were documented to not have mild cognitive impairment (MCI) and were between 60 and 100 years of age at assessment. Based on the data collected by NACC, the ADGC Neuropathology Core Leaders Subcommittee derived inclusion and exclusion criteria for Alzheimer's disease and control samples. All autopsied subjects were age ≥ 60 years at death. Based on the data collected by NACC, the ADGC Neuropathology Core Leaders Subcommittee derived inclusion and exclusion criteria for Alzheimer's disease and control samples. All autopsied subjects were age ≥ 60 years at death. Alzheimer's disease cases were demented according to NINCDS-ADRDA/DSMIV-V criteria or Clinical Dementia Rating (CDR) ≥ 137 (G. McKhann et al., 1984; G. M. McKhann et al., 2011). Neuropathologic stratification of cases followed NIA/Reagan criteria explicitly or used a similar approach when NIA/Reagan criteria were coded as not done, missing, or unknown. Cases were intermediate or high likelihood by NIA/Reagan criteria with moderate to frequent amyloid plaques (Mirra et al., 1993) and neurofibrillary tangle (NFT) Braak stage of III-VI (Braak & Braak, 1991; Nagy et al., 1998). Persons with Down's syndrome, non-Alzheimer's disease tauopathies and synucleinopathies were excluded. All autopsied controls had a clinical evaluation within two years of death. Controls did not meet NINCDS-ADRDA/DSMIV-V criteria for dementia, did not have a diagnosis of mild cognitive impairment (MCI), and had a CDR of 0, if performed. Controls did not meet or were low-likelihood Alzheimer's disease by NIA/Reagan criteria, had sparse or no amyloid plaques, and a Braak NFT stage of 0 – II. ADCs sent frozen tissue from autopsied subjects and DNA samples from some autopsied subjects and from living subjects to the ADCs to the National Cell Repository for Alzheimer's Disease (NCRAD). DNA was prepared by NCRAD for genotyping and sent to the genotyping site at Children's Hospital of Philadelphia. ADC samples were genotyped and analyzed in separate batches (waves 1-10). The ADC data used in the analyses (ADC1-10) consist of 6,292 cases and 4,980 CNEs in total.

ADNI: ADNI is a longitudinal, multi-site observational study including Alzheimer's disease, mild cognitive impairment (MCI), and elderly individuals with normal cognition assessing clinical and cognitive measures, MRI and PET scans (FDG and 11C PIB) and blood and CNS biomarkers. For this study, ADNI contributed data on 268 Alzheimer's disease cases with MRI confirmation of Alzheimer's disease diagnosis and 173 healthy controls with Alzheimer's disease-free status confirmed as of most recent follow-up. Alzheimer's disease subjects were between the ages of 55–90, had an MMSE score of 20–26 inclusive, met NINCDS-ADRDA criteria for probable Alzheimer's disease (G. McKhann et al., 1984; G. M. McKhann et al., 2011), and had an MRI consistent with the diagnosis of Alzheimer's disease. Control subjects had MMSE scores between 28 and 30 and a Clinical Dementia Rating of 0 without symptoms of depression, MCI or other dementia and no current use of psychoactive medications. According to the ADNI protocol, subjects were ascertained at regular intervals over 3 years, but for the purpose of our analysis we only used the final ascertainment status to classify case-control status. Additional details of the study design are available elsewhere (Gyungah Jun et al., 2010; Petersen et al., 2010).

GenADA: GenADA study data analyzed included 666 Alzheimer's disease cases and 712 CNEs ascertained from nine memory referral clinics in Canada between 2002 and 2005. Patients and CNEs were of non-Hispanic White (NHW) ancestry from Northern Europe. All patients with Alzheimer's disease satisfied NINCDS-ADRDA and DSM-IV criteria for probable Alzheimer's disease with Global Deterioration Scale scores of 3-7 (G. McKhann et al., 1984; G. M. McKhann et al., 2011). CNEs had MMSE test scores higher than 25 (mean 29.2 ± 1.1), a Mattis Dementia Rating Scale score of ≥ 136, a Clock Test without error, and no impairments on seven instrumental activities of daily living questions from the Duke Older American Resources and Services Procedures test. Data were collected under an academic-industrial grant from Glaxo-Smith-Kline, Canada by Principal Investigator P. St George-Hyslop. Detailed characteristics of this cohort have been described previously (Li et al., 2008).

MAYO/RMAYO: All 671 cases and 1,279 controls consisted of NHW subjects from the United States ascertained at the Mayo Clinic. All subjects were diagnosed by a neurologist at the Mayo Clinic in Jacksonville, Florida or Rochester, Minnesota. The neurologist confirmed a Clinical Dementia Rating score of 0 for all controls; cases had diagnoses of possible or probable Alzheimer's disease made according to NINCDS-ADRDA criteria (G. McKhann et al., 1984; G. M. McKhann et al., 2011). Autopsy-confirmed samples (221 cases, 216 CNEs) came from the brain bank at the Mayo Clinic in Jacksonville, FL and were evaluated by a single neuropathologist. In clinically-identified cases, the diagnosis of definite Alzheimer's disease was made according to NINCDS-ADRDA criteria. All Alzheimer's disease brains analyzed in the study had a Braak score of 4.0 or greater. Brains employed as controls had a Braak score of 2.5 or lower but often had brain pathology unrelated to Alzheimer's disease and pathological diagnoses that included vascular dementia, frontotemporal dementia, dementia with Lewy bodies, multi-system atrophy, amyotrophic lateral sclerosis, and progressive supranuclear palsy.

MIRAGE: The MIRAGE study is a family-based genetic epidemiology study of Alzheimer's disease that enrolled Alzheimer's disease cases and unaffected sibling controls at 17 clinical centers in the United States, Canada, Germany, and Greece (details elsewhere (Green et al., 2002)), and contributed 1,229 subjects (491 Alzheimer's disease cases and 738 CNEs), a subset of the cases and controls that were incorporated into our prior studies (Gyungah Jun et al., 2010; Naj et al., 2011) which met more stringent QC criteria for this study. Briefly, families were ascertained through a proband meeting the NINCDS-ADRDA criteria for definite or probable Alzheimer's disease (G. McKhann et al., 1984; G. M. McKhann et al., 2011). Unaffected sibling controls were verified as cognitively healthy based on a Modified Telephone Interview of Cognitive Status score ≥ 86 (Roccaforte et al., 1992).

UM/CWRU/TARCC2: The UM/CWRU/TARCC2 sample included 256 cases and 189 controls from the University of Miami, Case Western Reserve University, and the Texas Alzheimer's Research Care Consortium (wave 2). All Alzheimer's disease cases had onset of disease symptoms after age 65 years and met NINCDS-ADRDA criteria for probable or possible Alzheimer's disease (G. McKhann et al., 1984; G. M. McKhann et al., 2011). Controls were adjudicated to have MMSE scores greater than 28 and no clinically identified signs of cognitive impairment. Additional details of subject recruitment at these sites are described in the UM/CWRU/MSSM (formerly UM/VU/MSSM) and TARCC cohort descriptions in this supplement and elsewhere (G Jun et al., 2016; Naj et al., 2011; Sims et al., 2017).

NIA-LOAD: The NIA LOAD Family Study (Lee et al., 2008) recruited families with two or more affected siblings with LOAD and unrelated, CNEs similar in age and ethnic background. A total of 1,819 cases and 1,969 CNEs from 1,802 families were recruited through the NIA

LOAD study, NCRAD, and the University of Kentucky, with 1,798 cases and 1,568 CNEs included for analysis. One case per family was selected after determining the individual with the strictest diagnosis (definite > probable > possible LOAD). If there were multiple individuals with the strictest diagnosis, then the individual with the earliest age of onset was selected. The controls included only those samples that were neurologically evaluated to be normal and were not related to a study participant.

OHSU: The OHSU dataset includes 132 autopsy-confirmed Alzheimer's disease cases and 153 deceased controls that were evaluated for dementia within 12 months prior to death (age at death > 65 years), which are a subset of the 193 cases and 451 controls examined in our previous study (Gyungah Jun et al., 2010) meeting more stringent QC criteria in this study. Subjects were recruited from aging research cohorts at 10 NIA-funded ADC and did not overlap other samples assembled by the ADGC. A more extensive description of control samples can be found elsewhere (Kramer et al., 2011).

Pfizer: The Pfizer sample collection comprises Alzheimer's disease cases taken from the Lipitor's Effect in Alzheimer's Disease (LEADe) trial, including subjects who converted to Alzheimer's disease after ascertainment as MCI, as well as 216 probable Alzheimer's disease subjects enrolled by PrecisionMed for a case-control study and 149 subjects from a Phase II trial (#A3041005) of CP-457920 (a selective α5 GABAA receptor inverse agonist) in Alzheimer's disease. Samples were collected from multiple clinical sites, and with appropriate IRB/ethics committee approvals at each individual site, with written and informed consent given by subjects for use in follow-up studies. All subjects were diagnosed with probable or possible Alzheimer's disease if they met NINCDS-ADRDA and/or DSM-IV criteria, and had Mini-Mental Status Exam (MMSE) scores < 25 at baseline (G. McKhann et al., 1984; G. M. McKhann et al., 2011). The control group included subjects from two studies: 1) the PrecisionMed case-control study (#A9010012), which recruited elderly subjects free of neurological or psychiatric conditions, and 2) 999-GEN-0583-001, which obtained a reference population of cognitively, neurologically, and psychiatrically normal subjects. Controls have no neuropsychiatric conditions or diseases and had MMSE>27 at the time of enrollment. For Alzheimer's disease analysis, all cases with age-at-onset (AAO) less than 65 years were removed to exclude early-onset Alzheimer's disease subjects. All controls were re-matched with remaining cases according to gender, age (all controls are older than cases), and ethnicity (only individuals with NHW background were analyzed). The final Pfizer Alzheimer's disease case-control GWAS dataset included 696 cases and 762 controls. Cases from the PrecisionMed/ A3041005 and LEADe studies and age-matched controls were genotyped using the Illumina HumanHap550 array. APOE genotypes were determined from genotypes for rs429358 and rs7412 obtained using Taqman assays.

TARCC: The TARCC is a collaborative Alzheimer's research effort directed and funded by the Texas Council on Alzheimer's Disease and Related Disorders (the Council), as part of the

Darrell K Royal Texas Alzheimer's Initiative. Composed of Baylor College of Medicine (BCM), Texas Tech University Health Sciences Center (TTUHSC), University of North Texas Health Science Center (UNTHSC), the UT Southwestern Medical Center at Dallas (UTSW), University of Texas Health Science Center at San Antonio (UTHSCSA), Texas A&M Health Science Center (TAMHSC), and the University of Texas at Austin (UTA), this consortium was created to establish a comprehensive research cohort of well characterized subjects to address better diagnosis, treatment, and ultimately prevention of Alzheimer's disease (Hall et al., 2013). The resulting prospective cohort, the Texas Harris Alzheimer's Research Study, contains clinical, neuropsychiatric, genetic, and blood biomarker data on more than 3,000 participants diagnosed with Alzheimer's disease, mild cognitive impairment (MCI), and cognitively normal individuals. Longitudinal data/sample collection and follow-up on participants occurs on an annual basis. Two waves of case-control data from TARCC were examined as part of genetic analyses in the ADGC. Data from the TARCC included 323 cases and 181 controls in the first wave (included in the TARCC1 cohort), with 84 cases and 115 controls in the second wave (included in the UM/CWRU/TARCC2 cohort). All TARCC subjects were greater than 65 years of age at disease onset (cases) or at last disease-free exam (non-cases).

<u>TGEN2:</u> Among the TGEN2 data analyzed were 668 clinically- and neuropathologically-characterized brain donors, and 365 CNEs without dementia or significant Alzheimer's disease pathology. Of these cases and CNEs, 667 were genotyped as a part of the TGEN1 series (Reiman et al., 2007). Samples were obtained from twenty-one different National Institute on Aging-supported Alzheimer's disease Center brain banks and from the Miami Brain Bank as previously described (Caselli et al., 2007; Petyuk et al., 2018; Reiman et al., 2007; Webster et al., 2009). Additional individual samples from other brain banks in the United States, United Kingdom, and the Netherlands were also obtained in the same manner. The criteria for inclusion were as follows: self-defined ethnicity of European descent, neuropathologically confirmed Alzheimer's disease or neuropathology present at levels consistent with status as a control, and age of death greater than 65. Autopsy diagnosis was performed by board-certified neuropathologists and was based on the presence or absence of the characterization of probable or possible Alzheimer's disease. Where possible, Braak staging and/or CERAD classification were employed. Samples derived from subjects with a clinical history of stroke, cerebrovascular disease, comorbidity with any other known neurological disease, or with the neuropathological finding of Lewy bodies were excluded.

<u>UKS:</u> The UKS cohort is a thoroughly diagnosed case-control cohort from Universitätsklinikum des Saarlandes, consisting of individuals clinically diagnosed with sporadic Alzheimer's disease (N = 596; mean age onset, 72.2 ± 6.6 years) and cognitively healthy, age-, gender-, and ethnicity-matched population-based controls (N = 170; 64.1 ± 3.0 years).

UM/CWRU/MSSM: The UM/CWRU/MSSM dataset (formerly UM/VU/MSSM (Beecham et al., 2009; Edwards et al., 2010; Naj et al., 2010; Scott et al., 2001)) contains 1,177 cases and 1,126 CNEs ascertained at the University of Miami, Case Western Reserve University and Mt. Sinai School of Medicine, including 409 autopsy-confirmed cases and 136 controls, primarily from the Mt. Sinai School of Medicine (Haroutunian et al., 1998). An additional 16 cases were included and 34 controls excluded from the data analyzed in the Jun et al. 2010 study (Gyungah Jun et al., 2010). Each affected individual met NINCDS-ADRDA criteria for probably or definite Alzheimer's disease (G. McKhann et al., 1984; G. M. McKhann et al., 2011) with age at onset greater than 60 years as determined from specific probe questions within the clinical history provided by a reliable family informant or from documentation of significant cognitive impairment in the medical record. Cognitively healthy controls were unrelated individuals from the same catchment areas and frequency matched by age and gender, and had a documented MMSE or 3MS score in the normal range. Cases and controls had similar demographics: both had similar ages-at-onset/ages-at-exam of 71.1 (±17.4 SD) for cases and 73.5 (±10.6 SD) for controls, and cases and controls were 64.5% and 61.3% female, respectively.

UPITT: The University of Pittsburgh dataset contains 1,255 NHW Alzheimer's disease cases (of which 277 were autopsy-confirmed) recruited by the University of Pittsburgh Alzheimer's Disease Research Center, and 829 NHW, CNEs ages 60 and older (2 were autopsy-confirmed). All Alzheimer's disease cases met NINCDS-ADRDA criteria for probable or definite Alzheimer's disease (G. McKhann et al., 1984; G. M. McKhann et al., 2011). Additional details of the cohort used for GWAS have been previously published (Kamboh et al., 2012).

WASHU: An NHW LOAD case-control dataset consisting of 377 cases and 281 healthy elderly controls was used in analyses for this study. This dataset was split between two analysis datasets (WASHU1 and WASHU2). Participants were recruited as part of a longitudinal study of healthy aging and dementia. Diagnosis of dementia etiology was made in accordance with standard criteria and methods (Morris et al., 2006). Severity of dementia was assessed using the Clinical Dementia Rating scale (Hughes et al., 1982).

**CHARGE**

CHS

The Cardiovascular Health Study (CHS) is a population-based cohort study of risk factors for coronary heart disease and stroke in adults ≥65 years conducted across four field centers (Fried et al., 1991). The original predominantly European ancestry cohort of 5,201 persons was recruited in 1989-1990 from random samples of the Medicare eligibility lists; subsequently, an additional predominantly African-American cohort of 687 persons was enrolled for a total sample of 5,888. Blood samples were drawn from all participants at their baseline examination and DNA was subsequently extracted from available samples. Genotyping was performed at the General Clinical Research Center's Phenotyping/Genotyping Laboratory at Cedars-Sinai among CHS participants who consented to genetic testing and had DNA available using the Illumina 370CNV BeadChip system (for European ancestry participants, in 2007) or the Illumina HumanOmni1-Quad_v1 BeadChip system (for African-American participants, in 2010). CHS was approved by institutional review committees at each field center and individuals in the present analysis had available DNA and gave informed consent including consent to use of genetic information for the study of cardiovascular disease.

FHS

The Framingham Heart Study (FHS), started in 1948, is a three-generation community-based prospective cohort study. The FHS includes the Original cohort followed since 1948, the Offspring and their spouses followed since 1971, and the third generation enrolled in 2002. In this study, we included only original and offspring cohorts. The original cohort consisted of 5,209 adult men and women from Framingham, Massachusetts. Survivors undergo biennial examinations. The Offspring cohort is examined approximately once every 4 years. DNA extraction and genotyping were performed in the 1990s and we limited genetic analyses to high-quality samples. Prevalent study analyses included 1,787 participants aged 65 or older at DNA draw, excluding those with dementia other than AD. For incident analyses, 1,904 genotyped persons were included. The Institutional Review Board of the Boston Medical Campus approved the study. The Original cohort has been evaluated biennially since 1948, screened for dementia and AD in 1974-76, and under surveillance for incident cases since then. Offspring are examined every 4 years and screened for dementia using neuropsychological tests and brain MRI. Participants with baseline age <65 at DNA draw were excluded. Participants receive questionnaires, physical exams, and lab tests at clinic exams. Dementia screening and follow-up methods involve standardized neuropsychological tests, MMSE administration, and further testing for abnormalities. Neurological and neuropsychological examinations are conducted for suspected cognitive impairment, with a panel reviewing medical records for dementia determination based on DSM-IV and NINCDS-ADRDA criteria.

RS

This study included samples from the Rotterdam study (RS). RS is a prospective population-based study designed to investigate the etiology of age-related disorders. At the baseline examination in 1990-93, study recruited 7983 subjects ≥ 55 years of age from the Ommoord district of Rotterdam (RS-I). At the baseline entry and after every 3 to 4 years, all the study participants were extensively interviewed and physically examined at the dedicated research center. During 2000 to 2001, the baseline cohort (RS-I) was expanded by adding 3011 subjects ≥55 years of age, who were not yet part of RS-I (RS-II). Second expansion of RS was performed by recruiting 3932 persons having ≥45 years of age during 2006-2008 (RS-III). The study has been approved by the Medical Ethical Committee of Erasmus Medical Center and by the Ministry of Health, Welfare and Sport of the Netherlands. Written Informed consents were also obtained from each study participant to participate and to collect information from their treating physicians.

Blood was drawn for genotyping from participants of RS cohort during their first visit and DNA genotyping was performed at the internal genotyping facility of Erasmus Medical Center, Rotterdam. All samples were genotyped with the 550K, 550K duo, or 610K Illumina arrays.

**UK Biobank (UKB)**

We used the data August 2023 release of the UKB (application number 61054).

UKB-diagnosed: AD cases were extracted from UK Biobank self-report, ICD10 code G30 for diagnoses, primary care and cause of death. Our analysis included 3,865 diagnosed cases and 427,835 controls.

UKB-proxy: Participants were asked to report their parent dementia status and proxy AD/dementia cases included i) all female participants who reported at least one parent affected with dementia and ii) all male participants who reported an affected mother, in both cases either at baseline or follow up. Individuals who did not report dementia i) in both parents for females and ii) in mother only for males, were used as controls in the proxy AD/dementia analysis. Our analysis included 55,868 proxy cases of dementia and 235,171 proxy-controls.

**FinnGen**

Patients and control subjects in FinnGen provided informed consent for biobank research, based on the Finnish Biobank Act. Alternatively, separate research cohorts, collected prior the Finnish Biobank Act came into effect (in September 2013) and start of FinnGen (August 2017), were collected based on study-specific consents and later transferred to the Finnish biobanks after approval by Fimea (Finnish Medicines Agency), the National Supervisory Authority for Welfare and Health. Recruitment protocols followed the biobank protocols approved by Fimea. The Coordinating Ethics Committee of the Hospital District of Helsinki and Uusimaa (HUS) statement number for the FinnGen study is Nr HUS/990/2017.

The FinnGen study is approved by Finnish Institute for Health and Welfare (permit numbers: THL/2031/6.02.00/2017, THL/1101/5.05.00/2017, THL/341/6.02.00/2018, THL/2222/6.02.00/2018, THL/283/6.02.00/2019, THL/1721/5.05.00/2019 and

THL/1524/5.05.00/2020), Digital and population data service agency (permit numbers: VRK43431/2017-3, VRK/6909/2018-3, VRK/4415/2019-3), the Social Insurance Institution (permit numbers: KELA 58/522/2017, KELA 131/522/2018, KELA 70/522/2019, KELA 98/522/2019, KELA 134/522/2019, KELA 138/522/2019, KELA 2/522/2020, KELA 16/522/2020), Findata permit numbers THL/2364/14.02/2020, THL/4055/14.06.00/2020,,THL/3433/14.06.00/2020, THL/4432/14.06/2020, THL/5189/14.06/2020, THL/5894/14.06.00/2020, THL/6619/14.06.00/2020, THL/209/14.06.00/2021, THL/688/14.06.00/2021, THL/1284/14.06.00/2021, THL/1965/14.06.00/2021, THL/5546/14.02.00/2020, THL/2658/14.06.00/2021, THL/4235/14.06.00/2021 and Statistics Finland (permit numbers: TK-53-1041-17 and TK/143/07.03.00/2020 (earlier TK-53-90-20) TK/1735/07.03.00/2021).

The Biobank Access Decisions for FinnGen samples and data utilized in FinnGen Data Freeze 8 include: THL Biobank BB2017_55, BB2017_111, BB2018_19, BB_2018_34, BB_2018_67, BB2018_71, BB2019_7, BB2019_8, BB2019_26, BB2020_1, Finnish Red Cross Blood Service Biobank 7.12.2017, Helsinki Biobank HUS/359/2017, Auria Biobank AB17-5154 and amendment #1 (August 17 2020), AB20-5926 and amendment #1 (April 23 2020), Biobank Borealis of Northern Finland_2017_1013, Biobank of Eastern Finland 1186/2018 and amendment 22 § /2020, Finnish Clinical Biobank Tampere MH0004 and amendments (21.02.2020 & 06.10.2020), Central Finland Biobank 1-2017, and Terveystalo Biobank STB 2018001.

FinnGen research project is a public-private partnership combining genotype data from Finnish biobanks and digital health record data from Finnish health registries (Kurki et al., 2022). FinnGen utilizes biobank samples that consist of 1) prospective samples (‘new samples’) and 2) legacy samples.

'New samples' can be collected from voluntary individuals thought Hospital biobank, Terveystalo Biobank or Blood Service Biobank. Legacy samples are older sample cohorts that have been collected for a specific research project before the Finnish Biobank Act came into effect (September 2013) and have then been transferred to a biobank according to the Finnish Biobank Act 13 §. The ‘new samples’ were genotyped with FinnGen ThermoFisher Axiom custom array at the ThermoFisher genotyping service in San Diego, CA, US. The ‘legacy samples’ were genotyped over the years using various generations of Illumina and Affymetrix GWAS arrays.

We used the AD cases from the FinnGen Data Freeze 8 using G6_ALZHEIMER where cases are defined by having ICD-10 code G10 or ICD-9 code 3310 in either hospital discharge records or as the cause of death. In total, the G6_ALZHEIMER has 7,759 cases and 334,740 controls with high-quality genotypes and genotype-verified sex.

The following exclusions were applied: 1) all controls under the age of 30 at the common end date of follow-up for Data Freeze 8, Dec 31, 2019, and 2) all controls diagnosed with other dementias, *i.e.* whose inpatient or specialist outpatient HILMO registry data had any of the following ICD codes for hospital diagnosis or operation by the end of follow-up on the HILMO registry, Mar 24, 2021: ICD10 F01, F010-F013, F018, F019, F02, F020-F024, F028, F03, G310, G318; ICD9 290, 2901-4, 2908-9, 2941A, 3311A, 3312X, 3317, 3319X, 4378A, 4378X; ICD8 29009, 29011, 29018-9, 29209. After exclusions, there were 7,759 cases and 313,216 controls.

### B. Quality control

#### 1. EADB studies quality control

We applied the same X-chromosome quality control (QC) protocol to all EADB studies: EADB-core, EADI, GR@ACE/DEGESCO, GERAD, Bonn and DemGene.

All variants or samples failing the (Bellenguez et al., 2022) QC were excluded from the X-chromosome analysis. This QC consisted of assessment of chip's variants, variant intensity QC and autosomal sample QC (exclusion of individuals with high heterozygosity or missingness on the autosomes, of individuals with discordant genetic and clinical sex, of population outliers and of related individuals).

An additional sample and variant QC specific to the X-chromosome and adapted from the (Bellenguez et al., 2022) protocol was then performed. For the X-chromosome variant QC, only samples failing the heterozygosity, missingness or sex-check QC from the autosomal sample QC were removed. From this point, we replaced missing self-reported sex by genetic sex.

##### a. X-chromosome QC Protocol

The X-chromosome QC was applied to both PAR (pseudo autosomal region) and non-PAR regions (positions in assembly GRCh38: PAR1 = 10,001 – 2,781,478; PAR2 = 155,701,384 – 156,030,895; non-PAR = 2,781,479 – 155,701,383).

All the analyses of the X-chromosome QC were performed using PLINK (v1.9) [3].

**Sample Quality Control specific to the X-chromosome**

Pre-quality control. All variants failing those pre-QC criteria were excluded from all the sample QC steps:

- PAR variants showing departure from the Hardy-Weinberg equilibrium (HWE) in controls (P-value < $1x10^{-15}$);
- non-PAR variants showing departure from the HWE (P-value < $1x10^{-15}$) in female controls (or in female cases and controls if the number of controls was too low);
- variants showing a high missingness overall (> 0.025).

Sample QC. Were excluded:

- samples showing high missingness on X-chromosome (missingness > 0.02 in EADB-core and > 0.05 in all other studies) (including both PAR and non-PAR variants);
- male samples showing heterozygosity higher than 1% in non-PAR variants;
- samples for which genetic sex could not be determined.

**Variant Quality Control of the X-chromosome**

For the variant QC, the initial set of X-chromosome variants was used (re-integrating the variants failing the pre-QC of the sample QC). All samples failing the autosomal sample QC, or the sample QC specific to X-chromosome were removed for the variant QC of the X-chromosome.

1) Steps specific to the X-chromosome

non-PAR region. Were excluded variants:

- with missingness >0.05 in either males or females;
- with heterozygosity >0.01 in males;
- failing the HWE test (P-value < $5x10^{-8}$) in female controls (or in female cases and controls if the number of controls was too low).

2) Steps identical to autosomal variant QC

For the following steps, the same exclusion criteria as for the autosomes in (Bellenguez et al., 2022) protocol were applied to the X-chromosome variants.

PAR regions. Were excluded variants:

- showing a high missingness (> 0.05);
- failing the HWE tests (P-value < $5x10^{-8}$) in controls.

PAR and non-PAR regions. Were excluded variants:

- showing a differential missingness between cases and controls (Fisher's exact test P-value < $5x10^{-8}$) (if the samples are split in batches, the test was performed globally as well as for all the batches including both cases and controls and a variant was excluded if it failed in at least one test).
- failing the frequency checks. Population outliers were excluded for this step.
    - A frequency test comparing the allelic frequency in the study with the one in the reference panels (1) Genome Aggregation Database (Karczewski et al., 2020) (gnomAD) (Finnish and non-Finnish allele counts and frequencies were included) and (2) Haplotype Reference Consortium (McCarthy et al., 2016) (HRC) was performed. If the variant was not present in either panel, its allelic frequency was compared with the one in TopMED. The $\chi^2$ test threshold used was adapted to each study's sample size (see below).
    - If the study includes several sample batches, GWAS were performed between controls across batches using SNPTEST "newml" (Marchini et al., 2007) to assess the genotype frequency differences between the batches. The controls from a batch were compared to the ones from each of the other batches; we thus carried out as many GWAS as there are pairs of batches. For each GWAS,

the batches were converted into a binary variable and used as the analysis phenotype. Males' genotypes were coded 0/2 and females' genotypes were coded 0/1/2. We selected the batches with more than 400 controls. The related samples were also excluded for this step.

- failing ambiguous variants check. All ambiguous variants (A/T or C/G) with MAF > 0.4 were removed.
- failing duplicated variants check. For duplicated variants of the chip, only the copy with the minimum missingness was kept if both copies pass previous variant QC.

**Clinical data QC and definition of covariates**

For the association analyses, we additionally excluded controls with age below 30 and individuals with known pathogenic mutations.

The following covariates were included in some analyses:

- Principal components. The principal components used as adjustment in the analysis were computed using the flashPCA2 software, as reported previously (Abraham et al., 2017);
- Sex. Sex was defined as the self-reported sex, or, when missing, as the genetically determined sex. Samples with discordant sex between self-reported and genetically determined sex were excluded;
- Age. Age of AD cases was defined as the age at onset, if available. Otherwise, we used, by order of priority, the baseline age, the age at last exam and the age at death. For controls, age was defined as the age at last exam, and if not available the age at death and the baseline age, by order of priority;
- APOE ε4 and ε2. The number of APOE ε4 and ε2 alleles were coded 0, 1 or 2. APOE ε4 and ε2 were determined from the genotyped *APOE* status specified in the clinical file of the study. If unavailable in the clinical file of the study, *APOE ε4* and ε2 were defined using the imputed data; rs429358 and rs7412, the two *APOE* variants, had a good imputation quality ($r^2$ > 0.8) in all studies. For a given individual, genotypes of the two *APOE* variants were only considered if their probability was higher than 0.8. This means that *APOE* status could be missing even after imputation. For samples with both genotyped and imputed *APOE* status available, the *APOE s*tatus was set to missing if the genotyped and imputed statuses were different.
- Other study-specific variables, when necessary, such as the genotyping centre for EADB-core and the genotyping chip for Bonn (Supplementary Table S6).

b. X-chromosome QC specific thresholds per study

All the EADB studies X-chromosome sample and variant QC followed the described pipeline with the same metrics and thresholds, except when specified otherwise.

***European Alzheimer's Disease Initiative (EADI) Consortium***

For the frequency tests (1) only allele counts and frequencies from non-Finnish samples were extracted from the gnomAD reference panel and (2) the $\chi^2$ threshold used was set to 1,500.

After autosomal QC and exclusion of individuals with known pathogenic mutations, the EADI study was made up of 2,400 AD cases and 6,338 controls. After X-chromosome QC, the EADI study included 2,377 AD cases and 6,207 controls for 12,194 X-chromosome variants (including 20 from PAR1).

**Genetic and Environmental Risk in AD (GERAD)**

For the frequency tests, the $\chi^2$ threshold used was set to 500. All 10,641 variants passing QC were liftover from Assembly GrCh37 to GrCh38. 3,168 cases, 7,267 controls and 10,624 variants were used for imputation.

**The Norwegian DemGene Network**

The X-chromosome QC and imputation of the samples genotyped by DECODE and omni chips were performed separately.

*DECODE chip:* For all Hardy-Weinberg equilibrium tests, both female cases and controls were included, instead of only female controls as the number of controls was too low. For the frequency tests, the $\chi^2$ threshold used was set to 250. A total of 1 case and 1,892 variants (25 in PAR) were excluded in the X-chromosome QC. After autosomal QC, exclusion of individuals with known pathogenic mutations and X-chromosome QC, the DemGene DECODE chip batch included 299 cases, 11 controls and 15,685 variants (489 in PAR).

*Omni chip:* The DemGene omni chip batch is split in 14 sub-batches. The differential missingness between cases and controls test for variants was performed globally as well as for the 4 batches including both cases and controls with enough sample sizes and a variant was excluded if it failed in at least one test. For the frequency tests, the $\chi^2$ threshold used was set to 1,500. A GWAS across controls was also performed between the 4 batches with more than 400 controls (using the same pipeline and thresholds described above). 13 controls, 6 cases and 933 variants (20 in PAR) were excluded with the X-chromosome QC. After autosomal QC, exclusion of individuals with known pathogenic mutations and X-chromosome QC, DemGene omni chip included 1,392 cases, 7,301 controls and 16,530 variants (362 in PAR).

**Bonn studies**

The X-chromosome QC and imputation of the samples genotyped by dietBB and omni chips were performed separately.

*DietBB chip*: For the frequency tests (1) only allele counts and frequencies from non-Finnish samples were extracted from the gnomAD reference panel and (2) the $\chi^2$ threshold used was set to 250. 908 (29 in PAR) variants were excluded with the X-chromosome QC. No additional samples were excluded. After autosomal QC, exclusion of individuals with known pathogenic mutations and X-chromosome QC, Bonn dietBB chip batch included 139 cases, 177 controls and 21,627 variants (487 in PAR).

*Omni chip:* For the frequency tests (1) only allele counts and frequencies from non-Finnish samples were extracted from the gnomAD reference panel and (2) the $\chi^2$ threshold used was set to 500. 3 cases and 982 (59 in PAR) variants were excluded with the X-chromosome QC. After autosomal QC, exclusion of individuals with known pathogenic mutations and X-chromosome QC, Bonn omni chip batch included 496 cases, 1030 controls and 23,680 variants (791 in PAR).

**GR@ACE**

For the frequency tests, the $\chi^2$ threshold used was set to 1000. 146 cases, 20 controls and 304 (7 in PAR) variants were excluded with the X-chromosome QC. After autosomal QC, exclusion of individuals with known pathogenic mutations and X-chromosome QC, GR@ACE included 6,375 cases, 6,474 controls and 15,128 variants (54 in PAR).

**The European Alzheimer's Disease DNA Biobank dataset (EADB-core)**

The sample missingness threshold was set to 0.02 for EADB-core to remove less variants. For the frequency tests, the $\chi^2$ threshold used was set to 1000. A GWAS across controls was performed between the three genotyping centers of EADB-core. 172 cases, 201 controls and 1,095 (7 in PAR) variants were excluded with the X-chromosome QC. After autosomal QC, exclusion of individuals with known pathogenic mutations and X-chromosome QC, EADB-core included 19,977 cases, 21,525 controls and 16,943 variants (507 in PAR).

#### 2. ADGC quality control

For the X-chromosome QC, the same sample and variant QC used for the autosomes was applied, but additionally including the following steps:

- Samples showing high missingness on the X-chromosome, male samples showing high level of heterozygosity and samples for which genetic gender cannot be determined were excluded.

- X-chromosome non-PAR variants showing high missingness in either males or females or showing high heterozygosity in males were excluded.

#### 3. CHARGE quality control

**CHS**

Participant-level exclusions: European ancestry participants were excluded from the GWAS study sample due to the presence at study baseline of coronary heart disease, congestive heart failure, peripheral vascular disease, valvular heart disease, stroke or transient ischemic attack or lack of available DNA. Beyond laboratory genotyping failures, participants were excluded if they had a call rate<=95% or if their genotype was discordant with known sex or prior genotyping (to identify possible sample swaps). All non-European ancestry participants were excluded from the analysis. After quality control, genotyping was successful for 3,268 European ancestry participants.

SNP exclusions: In CHS, the following exclusions were applied to identify a final set of 306,655 autosomal SNPs: call rate < 97%, HWE P < $10^{-5}$, > 2 duplicate errors or Mendelian inconsistencies (for reference CEPH trios), heterozygote frequency = 0, SNP not found in HapMap. A similar X-chromosome QC than for the EADB studies was applied to CHS.

**FHS**

The same sample QC as autosomes were performed, with additional exclusions based on the following criteria:

- males with high level of heterozygosity;
- individuals for which genetic gender could not be determined;
- individuals with high missingness on the X-chromosome.

In the non-PAR region, were excluded variants:

- with high level of heterozygosity in males (> 1%);
- with high missingness in females or in males (>2% in females or males);
- with low MAF in females or in males (<1% in females or males):
- showing departure from HWE in female controls (p < 1e-6);
- showing differential missingness between males and females (p<1e-7).

In the PAR regions, the same exclusion criteria as for autosomes were used. Were excluded variants:

- with high missingness overall;
- with low MAF;
- showing departure from HWE.

**RS**

Genotyping quality control criteria include call rate < 95%, Hardy-Weinberg equilibrium *P* < $1.0 \times 10^{-6}$ and MAF < 1%. Moreover, study samples with excess autosomal heterozygosity, call rate < 97.5%, ethnic outliers and duplicate or family relationships were excluded during quality control analysis.

A similar X-chromosome QC than for the EADB studies was applied to RS.

#### 4. UKB Quality Control

The quality control of the UKB, including additional X-chromosome specific steps, is described in (Bycroft et al. 2018). The QC includes first a marker-based QC testing for batch, plates, and sex effect (genotype frequency differences), departure from HWE within each batch (only females included in the non-PAR region of the X chromosome) and discordance across control replicates. The genotype calls of the variants failing at least one test were set to missing. The p-value threshold used for the marker-based QC was set to $10^{-12}$. For the non-PAR region of the X chromosome all marker-based QC tests were performed separately using males-only (haploid), females-only (diploid), and both combined, but then used the smallest of the three p-. For the non-PAR region of the X chromosome all marker-based QC tests were performed separately using males-only (haploid), females-only (diploid), and both combined, but then used the smallest of the three p-. For the non-PAR region of the X chromosome all marker-based QC tests were performed separately using males-only (haploid), females-only (diploid), and both combined, but then used the smallest of the three p-values. For the non-PAR region of the X chromosome all marker-based QC tests were performed separately using males-only (haploid), females-only (diploid), and both combined, but then used the smallest of the three p-values.

Then, a sample QC was performed, removing samples with poor quality genotype calls, related individuals, population outliers, PC-adjusted heterozygosity above the mean (0.1903) and high missingness in the autosomes (0.05).

For our analysis, all related individuals up to third degree relatives were excluded, as well as all individuals of non-European ancestry. For UKB-diagnosed related individuals, controls were excluded over AD cases, while for UKB-proxy, controls were excluded over proxy-AD cases.

For UKB-proxy, participants were asked to report their parent dementia status and those who answered “Do not know” or “Prefer not to answer” were excluded from analyses. AD-diagnosed individuals among proxy-controls were excluded. All proxy controls whose parents age/age at death is missing or < 60 were removed.

An additional sex-chromosome QC step was applied: samples showing a putative sex chromosome aneuploidy were removed. After sample QC, markers that failed quality control in more than one batch, had a greater than 5% overall missing rate, and had a MAF of less than 0.0001 were removed.

#### 5. FinnGen Quality control

The genotype data processing from Data Freeze 7 onward was used (described in detail in: https://finngen.gitbook.io/finngen-handbook/finngen-data-specifics/red-library-data-individual-level-data/genotype-data/description-of-how-the-data-is-processed-in-refinery). Individuals with ambiguous sex, high genotype missingness (>5%), excess heterozygosity (+-3SD) and non-Finnish ancestry were excluded, and variants with high missingness (>2%), low HWE P-value (<1e-6) and low minor allele count (MAC<3) were excluded. No additional X-chromosome QC was performed in FinnGen, but a QC after imputation specific to the X-chromosome was performed (described below).

The covariates used in GWAS included the sex, age, defined as the age of first Alzheimer's diagnosis for cases and as the age at the common end date of follow-up for Data Freeze 8, Dec 31, 2019, for controls, population structure (the first 10 principal components), the main genotyping batches, and the *APOE* risk genotypes.

### C. Imputation

#### 1. TOPMed imputation for EADB studies

All samples and variants passing the X-chromosome QC were used as the input of the imputation process. Related samples and population outliers were not excluded for the imputation. All remaining heterozygous non-PAR variants in males were set as missing. Males were set as haploid in the non-PAR region (using +fixploidy bcftools (Danecek et al., 2021) plugin).

The imputation was performed on the Michigan Imputation Server (MIS) where the TOPMed Freeze5 reference panel was granted to the EADB consortium. The server version used was the 1.2.4 with Eagle v2.4 as the phasing software and Minimac4 v4-1.0.2 as the imputation software.

#### 2. TOPMed imputation for ADGC studies

The same pre-imputation protocol as for the EADB studies was followed. Samples were imputed with the TOPMed Freeze 8 reference panel.

#### 3. 1000 Genomes imputation for CHARGE

For CHS, after merging the genotypes from the two chips, a set of 10,377 X chromosome SNPs were used for imputation (updated to hg19 positions). MaCH was used to pre-phase the genotypes. The phased genotypes were imputed into a reference panel of 1,092 individuals of multiple ethnicities from the Phase1 version3 haplotypes of 1000 Genomes project using minimac (release stamp 2012-11-16). SNPs were excluded from analysis for variance of the allele dosage ≤0.01.

For FHS, heterozygous SNPs from the non-PAR region in males were set at missing and hemizygous males are treated as homozygous. Imputation was performed using 1000G data (Phase 1 v3, March 2012, MACGT1, ALL panel) as reference panel. MaCH was used to pre-phase the genotypes and IMPUTE2 and Minimac for the imputation.

A similar imputation protocol was followed for RS. The PAR regions were excluded for males. Imputation was performed using 1000G data (Phase 1 v3, March 2012, MACGT1, ALL panel) as reference panel. MaCH was used to pre-phase the genotypes and IMPUTE2 and Minimac for the imputation.

#### 4. Imputation for UKB

UKB dataset was phased with SHAPEIT3 and imputed with a new version of the IMPUTE2 program referred to as IMPUTE4 (Bycroft et al., 2018). The imputation panel used is a combination of HRC, UK10K and 1000 Genomes. All samples and variants passing the UKB autosomes and X-chromosome QC were used as the input of the imputation process: related individuals and population outliers were not excluded. As described in the IMPUTE2 X-chromosome imputation pipeline, males were set to haploid in the non-PAR region prior to imputation.

#### 5. FinnGen

The genotype data were imputed with a Finnish population specific reference panel, Sisu (V4), described in https://finngen.gitbook.io/finngen-handbook/finngen-data-specifics/red-library-data-individual-level-data/genotype-data/imputation-panel. Genotype imputation process is described in https://dx.doi.org/10.17504/protocols.io.xbgfijw. In the pipeline used by FinnGen R8 for the X-chromosome imputation, males were set as diploid for both the phasing and imputation. Thus, we included an additional QC after imputation for FinnGen.

Variant pre-QC. Were excluded in the pre-QC:

- variants showing departure from the HWE in female controls (p-value < $1x10^{-15}$);
- variants showing high missingness globally (> 0.025);
- variants in the X-transposed region (in Xq21.3, from position 89Mb to 93.5Mb in assembly 38).

All variants failing pre-QC were excluded to all the following sample QC steps.

Sample QC. Were excluded:

- samples showing missingness rate > 0.05 on the X-chromosome;
- male samples showing high level of heterozygosity (more than 1%).

Variant QC. For the variant QC, the initial set of X-chromosome variants was used (re-integrating the variants failing the pre-QC of the sample QC). All samples failing the general sample QC, or the X-chromosome specific sample QC were removed for the X-chromosome variant QC. Were excluded:

- variants showing high missingness in either males or females (>0.05);
- variants showing high heterozygosity in males (> 0.01);
- variants failing the HWE test (P-value < $5x10^{-8}$) in female controls;
- variants in the X-transposed region (in Xq21.3, from position 89Mb to 93.5Mb in assembly 38).

### E. Association tests

#### 1. EADB studies

For each EADB study (all case-control), we performed a logistic regression of AD status in males and females combined with an additive genetic model and a robust variance estimation with the snpStats (v 3.4) package in R (snpStats, 2023). We also performed sex stratified logistic regressions of the AD status on the genetic variants with an additive genetic model using SNPTEST (v 2.5.6) “newml” method (Marchini et al., 2007). Each stratified model included only samples from one of the subsets defined by sex (female-only or male-only). Additionally, a logistic regression of the AD status on the genetic variants was performed using SNPTEST “newml” method using a general genetic model and including only females. GP (genotype probabilities) were used for all models in snpStats and SNPTEST and males were coded as female homozygous (equivalent to genotype (G) = {0, 2} for males and G = {0, 1, 2} for females).

All analyses were adjusted on principal components and other study-specific variables, when necessary (Supplementary Table S6).

As sensitivity analyses, we also performed the sex-stratified additive and general genetic models adjusted on i) age and ii) age, *APOEε4* and *APOEε2* statuses.

#### 2. ADGC studies

The same protocol was followed for ADGC studies association tests.

#### 3. CHARGE studies

The same protocol was followed for CHARGE studies association tests.

#### 4. UKB

*UKB with diagnosed cases:*

We performed a sex-combined regression of the AD status on the genetic variants with an additive genetic model and adjusted on sex using a logistic mixed model as implemented in SAIGE (v1.0.9) with G = {0, 2} for males and G = {0, 1, 2} for females. We also ran sex-stratified regression with an additive genetic model. Dosages were used for all models. Analyses were adjusted on principal components and genotyping center. We also performed the sex-stratified models adjusted on i) age and ii) age, APOEε4 and APOEε2 statuses.

The genetic relatedness, used in the first step of the SAIGE analysis, was constructed from autosomal variants:

- that were genotyped;
- with MAF ≥1%;
- with HWE P >= $1 \times 10^{-15}$;
- with missingness < 0.01;
- not involved in inter-chromosomal LD (the list of those variants is available in the Supplementary Table 19 of REGENIE paper (Mbatchou et al., 2021));
- not in the *APOE* region (40 to 50 Mb on chr 19 in GrCh37 and GrCh38);
- not in regions of high LD;
- remaining after LD pruning using a $r^2$ threshold of 0.9 with a window size of 1,000 markers and a step size of 100 markers.

We set the option « impute_method » to « best guessed » in step 2.

<u>*UKB with proxy cases:*</u>

The association test on proxy status in UKB was performed separately for males and females using the SAIGE protocol described above, and a correction factor was applied to the association statistics.

Let us consider a variant with two alleles. We note $f_x$ and $f_{xx}$ the allelic frequency in males and females, respectively. Males X-chromosome is only transmitted by the mother. Thus, at the $n^{th}$ generation, we have the following frequency in males: $f_x(n) = f_{xx}(n-1)$. Females receive their X-chromosome from both parents. The allelic frequency in females at the $n^{th}$ generation is: $f_{xx}(n) = (f_{xx}(n-1) + f_x(n-1)) / 2$. If we compare allelic frequencies in males and females at the $n^{th}$ generation, we have:

$f_x(n) - f_{xx}(n) = - ½ (f_x(n-1) - f_{xx}(n-1)) = (- ½)^n (f_x(0) - f_{xx}(0))$, and thus $\lim(f_x(n) - f_{xx}(n))_{n->+inf} = 0$, which means that, at equilibrium (e): $f_x(e) = f_{xx}(e)$.

Thus, the frequency of the X-chromosome variants remains constant across generations and is the same in males and females. Then, the proxy-GWAS approach developed for the autosomes can also be applied to the XWAS. For females, both mother and father dementia statuses are considered, and a female is a proxy case if either the father or the mother is affected. Thus, the frequency in female AD-proxy is the same as in autosomes:

$f_p = (f_A + f_C)/2$, where $f_p$, $f_A$ and $f_C$ are the allele frequency in proxy cases, AD-cases and controls, respectively. For the males, only the status of the mother is considered, which means that the frequency of the males AD-proxy is: $f_p = f_A$

We thus performed a sex stratified regression of the AD-proxy status and included a correction factor of two on the $\beta$ and its corresponding standard error only for the female model.

However, as we did not consider the sex of the parent with this method, we did not use the sex-stratified models in either the sex-stratified analysis, or the e-XCI approach.

#### 5. FinnGen

The association analyses were performed in the FinnGen sandbox using a standard FinnGen-implemented WDL pipeline for REGENIE (v2.2.4) with a minor modification to enable the use of a plink file set as input (https://finngen.gitbook.io/documentation/v/r8/methods/phewas/logistic-regression).

We performed two sex-combined mixed logistic regression of the AD status on the genetic variants with an additive genetic model and adjusted on sex with REGENIE (v2.2.4) (default settings), one with r-XCI genotype coding (genotype (G) = {0, 2} for males and G = {0, 1, 2} for females) and one with e-XCI coding (G = {0, 1} for males and G = {0, 1, 2} for females). We also ran sex-stratified regression with an additive genetic model. Best guessed genotypes were used for all models. Analyses were adjusted on principal components and genotyping center. We also performed the sex-stratified models adjusted on i) age and ii) age, *APOEε4* and *APOEε2* statuses.

The genetic relatedness, used in the first step of the REGENIE analysis, was constructed only with autosomal markers. We thus used the same variants selected for the autosome analysis in the original FinnGen Data Freeze 8 genetic relationship matrix (GRM) file. In the GRM file were included variants 1) imputed with an INFO score > 0.95 in all batches and 2) with > 97 % non-missing genotypes and 3) MAF > 1 %. The remaining variants were LD pruned with a 1Mb window and a $r^2$ threshold of 0.1. The original FinnGen Data Freeze 8 GRM file was additionally modified to remove all variants present in the original GRM within ±1MB (43 variants) of the variant chr19_44870482_A_G (rs4081918) (the closest variant to the Alzheimer's risk variants in the *APOE* locus).

#### 6. Meta-analysis

The models used in the three approaches can be written as in Supplementary Table S6.

A r-XCI meta-analysis adjusted on sex (sensitivity analysis) was obtained from the meta-analysis of i) the sex-stratified models of case-control studies and the UKB and ii) the sex-combined model adjusted on sex for Finngen. For this meta-analysis, we included the sex-stratified models only adjusted on PCs (Supplementary Table S7).

FinnGen was excluded from the e-XCI meta-analyses adjusted on i) age and ii) age and *APOE*, and from the r-XCI meta-analyses adjusted on i) sex and age and ii) sex, age and *APOE*. Indeed, only the sex-stratified results were available for these models, which cannot be meta-analysed together because FinnGen samples are related.

Inflation of the test statistics was computed using only independent common variants, defined as variants 1) with MAF > 0.01 and 2) selected with the PLINK pruning procedure among EADB-core variants, by keeping only one variant from each pair of variants with $r^2$>0.8 and within 500 kb from each other. LD was computed in female samples only.

For results display, all r-XCI and s-XCI approaches OR and confidence intervals were rescaled to the real XCI coding (equivalent to G = {0, 1} for males and G = {0, 0.5, 1}).

##### 7. Sex-stratified analysis

The differences of effect between males and females were obtained using the sex-stratified meta-analyses; we computed the interaction p-value with a Wald test using the effect size ($\beta_i$) and corresponding standard error ($se_i$) of the interaction between two groups:

$\beta_i = \beta_F - \beta_M$ ; $se_i$ = square root($se_M^2 + se_F^2$), where $\beta_F$ and $\beta_M$ are the effect sizes of the female-only and male-only (reference) models, respectively, and $se_M$ and $se_F$ are their standard errors.

#### F. Supplementary analyses

Gene–based analyses were performed using MAGMA v1.08 (de Leeuw et al., 2015). The analysis was corrected for the number of variants in each gene, LD between variants and LD between genes. LD was computed from the EADB-core TOPMed imputed dataset using only genotypes with high imputation quality (at least one GP ≥ 0.9 in EADB-core). Each variant with a high imputation quality was assigned to its closest gene, using a window of 35 kb upstream and 10 kb downstream. We used q-values to account for multiple testing (804 genes were considered in the analysis). However, we did not identify any X chromosome gene significantly associated with AD risk at the X chromosome level, whatever the approach, with the gene-based analysis.

#### G. FinnGen Banner Authors

Aarno Palotie[1], Mark Daly[1], Bridget Riley-Gills[2], Howard Jacob[2], Coralie Viollet[3], Slavé Petrovski[3], Chia-Yen Chen[4], Sally John[4], George Okafo[5], Robert Plenge[6], Joseph Maranville[6], Mark McCarthy[7], Rion Pendergrass[7], Margaret G. Ehm[8], Kirsi Auro[9], Simonne Longerich[10], Anders Mälarstig[11], Anna Vlahiotis[11], Katherine Klinger[12], Clement Chatelain[12], Matthias Gossel[12], Karol Estrada[13], Robert Graham[13], Dawn Waterworth[14], Chris O´Donnell[15], Nicole Renaud[15], Tomi P. Mäkelä[16], Jaakko Kaprio[17], Minna Ruddock[18], Petri Virolainen[19], Antti Hakanen[19], Terhi Kilpi[20], Markus Perola[20], Jukka Partanen[21], Taneli Raivio[22], Jani Tikkanen[23], Raisa Serpi[23], Kati Kristiansson[24], Veli-Matti Kosma[25], Jari Laukkanen[26], Marco Hautalahti[27], Outi Tuovila[28], Jeffrey Waring[2], Bridget Riley-Gillis[2], Fedik Rahimov[2], Ioanna Tachmazidou[3], Chia-Yen Chen[4], Zhihao Ding[5], Marc Jung[5], Hanati Tuoken[5], Shameek Biswas[6], Rion Pendergrass[7], Margaret G. Ehm[8], David Pulford[29], Neha Raghavan[10], Adriana Huertas-Vazquez[10], Jae-Hoon Sul[10], Anders Mälarstig[11], Xinli Hu[11], Åsa Hedman[11], Katherine Klinger[12], Robert Graham[13], Dawn Waterworth[14], Nicole Renaud[15], Ma´en Obeidat[15], Jonathan Chung[15], Jonas Zierer[15], Mari Niemi[15], Samuli Ripatti[17], Johanna Schleutker[30], Markus Perola[20], Mikko Arvas[21], Olli Carpén[22], Reetta Hinttala[23], Johannes Kettunen[23], Arto Mannermaa[25], Katriina Aalto-Setälä[31], Mika Kähönen[24], Jari Laukkanen[26], Johanna Mäkelä[27], Reetta Kälviäinen[32], Valtteri Julkunen[32], Hilkka Soininen[32], Anne Remes[33], Mikko Hiltunen[34], Jukka Peltola[35], Minna Raivio[36], Pentti Tienari[36], Juha Rinne[37], Roosa Kallionpää[37], Juulia Partanen[38], Adam Ziemann[2], Nizar Smaoui[2], Anne Lehtonen[2], Susan Eaton[4], Heiko Runz[4], Sanni Lahdenperä[4], Shameek Biswas[6], Natalie Bowers[7], Edmond Teng[7], Rion Pendergrass[7], Fanli Xu[39], David Pulford[29], Kirsi Auro[9], Laura Addis[39], John Eicher[39], Qingqin S Li[40], Karen He[14], Ekaterina Khramtsova[14], Neha Raghavan[10], Martti Färkkilä[36], Jukka Koskela[36], Sampsa Pikkarainen[36], Airi Jussila[35], Katri Kaukinen[35], Timo Blomster[33], Mikko Kiviniemi[32], Markku Voutilainen[37], Mark Daly[41], Jeffrey Waring[2], Nizar Smaoui[2], Fedik Rahimov[2], Anne Lehtonen[2], Tim Lu[7], Natalie Bowers[7], Rion Pendergrass[7], Linda McCarthy[39], Amy Hart[14], Meijian Guan[14], Jason Miller[10], Kirsi Kalpala[11], Melissa Miller[11], Xinli Hu[11], Kari Eklund[36], Antti Palomäki[37], Pia Isomäki[35], Laura Pirilä[37], Oili Kaipiainen-Seppänen[32], Johanna Huhtakangas[33], Nina Mars[17], Jeffrey Waring[2], Fedik Rahimov[2], Apinya Lertratanakul[2], Nizar Smaoui[2], Anne Lehtonen[2], Coralie Viollet[42], Marla Hochfeld[6], Natalie Bowers[7], Rion Pendergrass[7], Jorge Esparza Gordillo[39], Kirsi Auro[9], Dawn Waterworth[14], Fabiana Farias[10], Kirsi Kalpala[11], Nan Bing[11], Xinli Hu[11], Tarja Laitinen[35], Margit Pelkonen[32], Paula Kauppi[36], Hannu Kankaanranta[43], Terttu Harju[33], Riitta Lahesmaa[37], Nizar Smaoui[2], Coralie Viollet[42], Susan Eaton[4], Hubert Chen[7], Rion Pendergrass[7], Natalie Bowers[7], Joanna Betts[39], Kirsi Auro[9], Rajashree Mishra[39], Majd Mouded[44], Debby Ngo[44], Teemu Niiranen[45], Felix Vaura[45], Veikko Salomaa[45], Kaj Metsärinne[37], Jenni Aittokallio[37], Mika Kähönen[35], Jussi Hernesniemi[35], Daniel Gordin[36], Juha Sinisalo[36], Marja-Riitta Taskinen[36], Tiinamaija Tuomi[36], Timo Hiltunen[36], Jari Laukkanen[46], Amanda Elliott[47], Mary Pat Reeve[17], Sanni Ruotsalainen[17], Dirk Paul[3], Natalie Bowers[7], Rion Pendergrass[7], Audrey Chu[39], Kirsi Auro[9], Dermot Reilly[48], Mike Mendelson[49], Jaakko

Parkkinen[11], Melissa Miller[11], Tuomo Meretoja[50], Heikki Joensuu[51], Olli Carpén[36], Johanna Mattson[36], Eveliina Salminen[36], Annika Auranen[52], Peeter Karihtala[51], Päivi Auvinen[32], Klaus Elenius[37], Johanna Schleutker[37], Esa Pitkänen[17], Nina Mars[17], Mark Daly[1], Relja Popovic[2], Jeffrey Waring[2], Bridget Riley-Gillis[2], Anne Lehtonen[2], Margarete Fabre[42], Jennifer Schutzman[7], Natalie Bowers[7], Rion Pendergrass[7], Diptee Kulkarni[39], Kirsi Auro[9], Alessandro Porello[14], Andrey Loboda[10], Heli Lehtonen[11], Stefan McDonough[11], Sauli Vuoti[53], Kai Kaarniranta[54], Joni A Turunen[55], Terhi Ollila[36], Hannu Uusitalo[35], Juha Karjalainen[17], Esa Pitkänen[17], Mengzhen Liu[2], Heiko Runz[4], Stephanie Loomis[4], Erich Strauss[7], Natalie Bowers[7], Hao Chen[7], Rion Pendergrass[7], Kaisa Tasanen[33], Laura Huilaja[33], Katariina Hannula-Jouppi[36], Teea Salmi[35], Sirkku Peltonen[37], Leena Koulu[37], Nizar Smaoui[2], Fedik Rahimov[2], Anne Lehtonen[2], David Choy[7], Rion Pendergrass[7], Dawn Waterworth[14], Kirsi Kalpala[11], Ying Wu[11], Pirkko Pussinen[36], Aino Salminen[36], Tuula Salo[36], David Rice[36], Pekka Nieminen[36], Ulla Palotie[36], Maria Siponen[32], Liisa Suominen[32], Päivi Mäntylä[32], Ulvi Gursoy[37], Vuokko Anttonen[33], Kirsi Sipilä[56], Rion Pendergrass[7], Hannele Laivuori[17], Venla Kurra[35], Laura Kotaniemi-Talonen[35], Oskari Heikinheimo[36], Ilkka Kalliala[36], Lauri Aaltonen[36], Varpu Jokimaa[37], Johannes Kettunen[33], Marja Vääräsmäki[33], Outi Uimari[33], Laure Morin-Papunen[33], Maarit Niinimäki[33], Terhi Piltonen[33], Katja Kivinen[17], Elisabeth Widen[17], Taru Tukiainen[17], Mary Pat Reeve[17], Mark Daly[1], Niko Välimäki[57], Eija Laakkonen[58], Jaakko Tyrmi[59], Heidi Silven[60], Eeva Sliz[60], Riikka Arffman[60], Susanna Savukoski[60], Triin Laisk[61], Natalia Pujol[61], Mengzhen Liu[2], Bridget Riley-Gillis[2], Rion Pendergrass[7], Janet Kumar[8], Kirsi Auro[9], Iiris Hovatta[62], Chia-Yen Chen[4], Erkki Isometsä[36], Hanna Ollila[17], Jaana Suvisaari[45], Antti Mäkitie[63], Argyro Bizaki-Vallaskangas[35], Sanna Toppila-Salmi[64], Tytti Willberg[37], Elmo Saarentaus[17], Antti Aarnisalo[36], Eveliina Salminen[36], Elisa Rahikkala[33], Johannes Kettunen[33], Kristiina Aittomäki[65], Fredrik Åberg[66], Mitja Kurki[67], Samuli Ripatti[17], Mark Daly[41], Juha Karjalainen[17], Aki Havulinna[68], Juha Mehtonen[17], Priit Palta[17], Shabbeer Hassan[17], Pietro Della Briotta Parolo[17], Wei Zhou[69], Mutaamba Maasha[69], Shabbeer Hassan[17], Susanna Lemmelä[17], Manuel Rivas[70], Aarno Palotie[17], Aoxing Liu[17], Arto Lehisto[17], Andrea Ganna[17], Vincent Llorens[17], Hannele Laivuori[17], Taru Tukiainen[17], Mary Pat Reeve[17], Henrike Heyne[17], Nina Mars[17], Joel Rämö[17], Elmo Saarentaus[17], Hanna Ollila[17], Rodos Rodosthenous[17], Satu Strausz[17], Tuula Palotie[71], Kimmo Palin[57], Javier Garcia-Tabuenca[72], Harri Siirtola[72], Tuomo Kiiskinen[17], Jiwoo Lee[67], Kristin Tsuo[67], Amanda Elliott[47], Kati Kristiansson[20], Mikko Arvas[21], Kati Hyvärinen[73], Jarmo Ritari[73], Olli Carpén[22], Johannes Kettunen[23], Katri Pylkäs[60], Eeva Sliz[60], Minna Karjalainen[60], Tuomo Mantere[23], Eeva Kangasniemi[24], Sami Heikkinen[34], Arto Mannermaa[25], Eija Laakkonen[58], Nina Pitkänen[19], Samuel Lessard[12], Clément Chatelain[12], Lila Kallio[19], Tiina Wahlfors[20], Jukka Partanen[21], Eero Punkka[22], Raisa Serpi[23], Sanna Siltanen[24], Veli-Matti Kosma[25], Teijo Kuopio[26], Anu Jalanko[17], Huei-Yi Shen[17], Risto Kajanne[17], Mervi Aavikko[17], Helen Cooper[17], Denise Öller[17], Rasko Leinonen[74], Henna Palin[24], Malla-Maria Linna[22], Mitja Kurki[67], Juha Karjalainen[17], Pietro Della Briotta Parolo[17], Arto Lehisto[17], Juha Mehtonen[17], Wei Zhou[69], Masahiro Kanai[69], Mutaamba Maasha[69], Zhili Zheng[69], Hannele Laivuori[17], Aki Havulinna[68], Susanna Lemmelä[17], Tuomo Kiiskinen[17], L. Elisa Lahtela[17], Mari Kaunisto[17], Elina Kilpeläinen[17], Timo P. Sipilä[17], Oluwaseun Alexander Dada[17], Awaisa Ghazal[17], Anastasia

Kytölä[17], Rigbe Weldatsadik[17], Sanni Ruotsalainen[17], Kati Donner[17], Timo P. Sipilä[17], Anu Loukola[22], Päivi Laiho[20], Tuuli Sistonen[20], Essi Kaiharju[20], Markku Laukkanen[20], Elina Järvensivu[20], Sini Lähteenmäki[20], Lotta Männikkö[20], Regis Wong[20], Auli Toivola[20], Minna Brunfeldt[20], Hannele Mattsson[20], Kati Kristiansson[20], Susanna Lemmelä[17], Sami Koskelainen[20], Tero Hiekkalinna[20], Teemu Paajanen[20], Priit Palta[17], Shuang Luo[17], Tarja Laitinen[35], Mary Pat Reeve[17], Shanmukha Sampath Padmanabhuni[17], Marianna Niemi[72], Harri Siirtola[72], Javier Gracia-Tabuenca[72], Mika Helminen[72], Tiina Luukkaala[72], Iida Vähätalo[72], Jyrki Tammerluoto[17], Marco Hautalahti[75], Johanna Mäkelä[75], Sarah Smith[75], Tom Southerington[75], Petri Lehto[75]

[1]Institute for Molecular Medicine Finland (FIMM), HiLIFE, University of Helsinki, Helsinki, Finland; Broad Institute of MIT and Harvard; Massachusetts General Hospital
[2]Abbvie, Chicago, IL, United States
[3]Astra Zeneca, Cambridge, United Kingdom
[4]Biogen, Cambridge, MA, United States
[5]Boehringer Ingelheim, Ingelheim am Rhein, Germany
[6]Bristol Myers Squibb, New York, NY, United States
[7]Genentech, San Francisco, CA, United States
[8]GlaxoSmithKline, Collegeville, PA, United States
[9]GlaxoSmithKline, Espoo, Finland
[10]Merck, Kenilworth, NJ, United States
[11]Pfizer, New York, NY, United States
[12]Translational Sciences, Sanofi R&D, Framingham, MA, USA
[13]Maze Therapeutics, San Francisco, CA, United States
[14]Janssen Research & Development, LLC, Spring House, PA, United States
[15]Novartis Institutes for BioMedical Research, Cambridge, MA, United States
[16]HiLIFE, University of Helsinki, Finland, Finland
[17]Institute for Molecular Medicine Finland (FIMM), HiLIFE, University of Helsinki, Helsinki, Finland
[18]Arctic biobank / University of Oulu
[19]Auria Biobank / University of Turku / Hospital District of Southwest Finland, Turku, Finland
[20]THL Biobank / Finnish Institute for Health and Welfare (THL), Helsinki, Finland
[21]Finnish Red Cross Blood Service / Finnish Hematology Registry and Clinical Biobank, Helsinki, Finland
[22]Helsinki Biobank / Helsinki University and Hospital District of Helsinki and Uusimaa, Helsinki
[23]Northern Finland Biobank Borealis / University of Oulu / Northern Ostrobothnia Hospital District, Oulu, Finland
[24]Finnish Clinical Biobank Tampere / University of Tampere / Pirkanmaa Hospital District, Tampere, Finland

[25]Biobank of Eastern Finland / University of Eastern Finland / Northern Savo Hospital District, Kuopio, Finland
[26]Central Finland Biobank / University of Jyväskylä / Central Finland Health Care District, Jyväskylä, Finland
[27]FINBB - Finnish biobank cooperative
[28]Business Finland, Helsinki, Finland
[29]GlaxoSmithKline, Stevenage, United Kingdom
[30]Auria Biobank / Univ. of Turku / Hospital District of Southwest Finland, Turku, Finland
[31]Faculty of Medicine and Health Technology, Tampere University, Tampere, Finland
[32]Northern Savo Hospital District, Kuopio, Finland
[33]Northern Ostrobothnia Hospital District, Oulu, Finland
[34]University of Eastern Finland, Kuopio, Finland
[35]Pirkanmaa Hospital District, Tampere, Finland
[36]Hospital District of Helsinki and Uusimaa, Helsinki, Finland
[37]Hospital District of Southwest Finland, Turku, Finland
[38]Institute for Molecular Medicine Finland, HiLIFE, University of Helsinki, Finland
[39]GlaxoSmithKline, Brentford, United Kingdom
[40]Janssen Research & Development, LLC, Titusville, NJ 08560, United States
[41]Institute for Molecular Medicine, Finland (FIMM), HiLIFE, University of Helsinki, Helsinki, Finland; Broad Institute of MIT and Harvard; Massachusetts General Hospital
[42]AstraZeneca, Cambridge, United Kingdom
[43]University of Gothenburg, Gothenburg, Sweden/ Seinäjoki Central Hospital, Seinäjoki, Finland/ Tampere University, Tampere, Finland
[44]Novartis, Basel, Switzerland
[45]Finnish Institute for Health and Welfare (THL), Helsinki, Finland
[46]Central Finland Health Care District, Jyväskylä, Finland
[47]Institute for Molecular Medicine Finland (FIMM), HiLIFE, University of Helsinki, Helsinki, Finland; Broad Institute, Cambridge, MA, USA and Massachusetts General Hospital, Boston, MA, USA
[48]Janssen Research & Development, LLC, Boston, MA, United States
[49]Novartis, Boston, MA, United States
[50]Department of Breast Surgery, Helsinki University Hospital Comprehensive Cancer Center and University of Helsinki, Helsinki, Finland
[51]Department of Oncology, Helsinki University Hospital Comprehensive Cancer Center and University of Helsinki, Helsinki, Finland
[52]Pirkanmaa Hospital District , Tampere, Finland
[53]Janssen-Cilag Oy, Espoo, Finland
[54]Northern Savo Hospital District, Kuopio, Finland; Department of Molecular Genetics, University of Lodz, Lodz, Poland
[55]Helsinki University Hospital and University of Helsinki, Helsinki, Finland; Eye Genetics Group, Folkhälsan Research Center, Helsinki, Finland

[56]Research Unit of Oral Health Sciences Faculty of Medicine, University of Oulu, Oulu, Finland; Medical Research Center, Oulu, Oulu University Hospital and University of Oulu, Oulu, Finland
[57]University of Helsinki, Helsinki, Finland
[58]University of Jyväskylä, Jyväskylä, Finland
[59]University of Oulu, Oulu, Finland / University of Tampere, Tampere, Finland
[60]University of Oulu, Oulu, Finland
[61]Estonian biobank, Tartu, Estonia
[62]University of Helsinki, Finland
[63]Department of Otorhinolaryngology - Head and Neck Surgery, University of Helsinki and Helsinki University Hospital, Helsinki, Finland
[64]University of Eastern Finland and Kuopio University Hospital, Department of Otorhinolaryngology, Kuopio, Finland and Department of Allergy, Helsinki University Hospital and University of Helsinki, Finland
[65]Department of Medical Genetics, Helsinki University Central Hospital, Helsinki, Finland
[66]Transplantation and Liver Surgery Clinic, Helsinki University Hospital, Helsinki University, Helsinki, Finland
[67]Institute for Molecular Medicine Finland (FIMM), HiLIFE, University of Helsinki, Helsinki, Finland; Broad Institute, Cambridge, MA, United States
[68]Institute for Molecular Medicine Finland (FIMM), HiLIFE, University of Helsinki, Helsinki, Finland; Finnish Institute for Health and Welfare (THL), Helsinki, Finland
[69]Broad Institute, Cambridge, MA, United States
[70]University of Stanford, Stanford, CA, United States
[71]University of Helsinki and Hospital District of Helsinki and Uusimaa, Helsinki, Finland
[72]University of Tampere, Tampere, Finland
[73]Finnish Red Cross Blood Service, Helsinki, Finland
[74]Institute for Molecular Medicine Finland (FIMM), HiLIFE, University of Helsinki, Helsinki, Finland; European Molecular Biology Laboratory, European Bioinformatics Institute, Cambridge, UK
[75]Finnish Biobank Cooperative - FINBB

#### H. Supplementary acknowledgments

**Additional support for EADB cohorts:** The work for this manuscript was further supported by the CoSTREAM project (www.costream.eu) and funding from the European Union's Horizon 2020 research and innovation programme under grant agreement No 667375. This work is also funded by la Fondation pour la Recherche Médicale (FRM) (EQU202003010147); this work was also supported by Italian Ministry of Health (Ricerca Corrente) (R.G., IRCCS Istituto Centro San Giovanni di Dio Fatebenefratelli, Brescia); Ministero dell'Istruzione, dell'Università e della Ricerca–MIUR project "Dipartimenti di Eccellenza 2018–2022" to Department of Neuroscience "Rita Levi Montalcini", University of Torino (IR), and AIRAlzh Onlus-ANCC-COOP (SB); Partly supported by "Ministero della Salute", I.R.C.C.S. Research Program, Ricerca Corrente 2018-2020, Linea n. 2 "Meccanismi genetici, predizione e terapie innovative delle malattie complesse" and by the "5 x 1000" voluntary contribution to the Fondazione I.R.C.C.S. Ospedale "Casa Sollievo della Sofferenza"; and RF-2018-12366665, Fondi per la ricerca 2019 (Sandro Sorbi). Copenhagen General Population Study (CGPS): We thank staff and participants of the CGPS for their important contributions. Karolinska Institutet AD cohort: Dr. C.G. and co-authors of the Karolinska Institutet AD cohort report grants from Swedish Research Council (VR) 2015-02926, 2018-02754, 2015-06799, Swedish Alzheimer Foundation, Stockholm County Council ALF and resarch school, Karolinska Institutet StratNeuro, Swedish Demensfonden, and Swedish brain foundation, during the conduct of the study. ADGEN: This work was supported by Academy of Finland (grant numbers 307866); Sigrid Jusélius Foundation; the Strategic Neuroscience Funding of the University of Eastern Finland; EADB project in the JPNDCO-FUND program (grant number 301220). CBAS: Supported by the project no. LQ1605 from the National Program of Sustainability II (MEYS CR), Supported by Ministry of Health of the Czech Republic, grant nr. NV19-04-00270 (All rights reserved), Grant Agency of Charles University Grants No. 693018 and 654217; the Ministry of Health, Czech Republic—conceptual development of research organization, University Hospital Motol, Prague, Czech Republic Grant No. 00064203; the Czech Ministry of Health Project AZV Grant No. 16—27611A; and Institutional Support of Excellence 2. LF UK Grant No. 699012. CNRMAJ-Rouen: This study received fundings from the Centre National de Référence Malades Alzheimer Jeunes (CNRMAJ). The Finnish Geriatric Intervention Study for the Prevention of Cognitive Impairment and Disability (FINGER) data collection was supported by grants from the Academy of Finland, La Carita Foundation, Juho Vainio Foundation, Novo Nordisk Foundation, Finnish Social Insurance Institution, Ministry of Education and Culture Research Grants, Yrjö Jahnsson Foundation, Finnish Cultural Foundation South Osthrobothnia Regional Fund, and EVO/State Research Funding grants of University Hospitals of Kuopio, Oulu and Turku, Seinäjoki Central Hospital and Oulu City Hospital, Alzheimer's Research & Prevention Foundation USA, AXA Research Fund, Knut and Alice Wallenberg Foundation Sweden, Center for Innovative Medicine (CIMED) at Karolinska Institutet Sweden, and Stiftelsen Stockholms sjukhem Sweden.

FINGER cohort genotyping was funded by EADB project in the JPND CO-FUND (grant number 301220). Research at the Belgian EADB site is funded in part by the Alzheimer Research Foundation (SAO-FRA), The Research Foundation Flanders (FWO), and the University of Antwerp Research Fund. FK is supported by a BOF DOCPRO fellowship of the University of Antwerp Research Fund. SNAC-K is financially supported by the Swedish Ministry of Health and Social Affairs, the participating County Councils and Municipalities, and the Swedish Research Council. BDR Bristol: We would like to thank the South West Dementia Brain Bank (SWDBB) for providing brain tissue for this study. The SWDBB is part of the Brains for Dementia Research programme, jointly funded by Alzheimer’s Research UK and Alzheimer’s Society and is supported by BRACE (Bristol Research into Alzheimer’s and Care of the Elderly) and the Medical Research Council. BDR Manchester: We would like to thank the Manchester Brain Bankfor providing brain tissue for this study. The Manchester Brain Bank is part of the Brains for Dementia Research programme, jointly funded by Alzheimer’s Research UK and Alzheimer’s Society. BDR KCL: Human post-mortem tissue was provided by the London Neurodegenerative Diseases Brain Bank which receives funding from the UK Medical Research Council and as part of the Brains for Dementia Research programme, jointly funded by Alzheimer’s Research UK and the Alzheimer’s Society. The CFAS Wales study was funded by the ESRC (RES-060-25-0060) and HEFCW as ‘Maintaining function and well-being in later life: a longitudinal cohort study’. We are grateful to the NISCHR Clinical Research Centre for their assistance in tracing participants and in interviewing and in collecting blood samples, and to general practices in the study areas for their cooperation. MRC: We thank all individuals who participated in this study. Cardiff University was supported by the Alzheimer's Society (AS; grant RF014/164) and the Medical Research Council (MRC; grants G0801418/1, MR/K013041/1, MR/L023784/1) (R.S. is an AS Research Fellow). Cardiff University was also supported by the European Joint Programme for Neurodegenerative Disease (JPND; grant MR/L501517/1), Alzheimer's Research UK (ARUK; grant ARUK-PG2014-1), the Welsh Assembly Government (grant SGR544:CADR), Brain’s for dementia Research and a donation from the Moondance Charitable Foundation. Cardiff University acknowledges the support of the UK Dementia Research Institute, of which J.W. is an associate director. Cambridge University acknowledges support from the MRC. Patient recruitment for the MRC Prion Unit/UCL Department of Neurodegenerative Disease collection was supported by the UCLH/UCL Biomedical Centre and NIHR Queen Square Dementia Biomedical Research Unit. The University of Southampton acknowledges support from the AS. King's College London was supported by the NIHR Biomedical Research Centre for Mental Health and the Biomedical Research Unit for Dementia at the South London and Maudsley NHS Foundation Trust and by King's College London and the MRC. ARUK and the Big Lottery Fund provided support to Nottingham University. A.Ram. : Part of the work was funded by the JPND EADB grant (German Federal Ministry of Education and Research (BMBF) grant: 01ED1619A). A. Ram. is also supported by the German Research Foundation (DFG) grants Nr: RA 1971/6-1, RA1971/7-1, and RA 1971/8-1. German Study on Ageing, Cognition and Dementia in Primary Care Patients (AgeCoDe): This study/publication is part of the German Research Network on Dementia (KND), the German Research Network on Degenerative Dementia (KNDD; German Study on Ageing, Cognition and Dementia in Primary Care Patients; AgeCoDe), and the Health Service Research Initiative (Study on Needs, health service use, costs and health-related quality of life in a large sample of oldestold primary care patients (85+; AgeQualiDe)) and was funded by the German Federal Ministry of Education and Research (grants KND: 01GI0102, 01GI0420, 01GI0422, 01GI0423, 01GI0429, 01GI0431, 01GI0433, 01GI0434; grants KNDD: 01GI0710, 01GI0711, 01GI0712, 01GI0713, 01GI0714, 01GI0715, 01GI0716; grants Health Service Research Initiative: 01GY1322A, 01GY1322B, 01GY1322C, 01GY1322D, 01GY1322E, 01GY1322F, 01GY1322G). VITA study: The support of the Ludwig Boltzmann Society and the AFI Germany have supported the VITA study. The former VITA study group should be acknowledged: W. Danielczyk, G. Gatterer, K Jellinger, S Jugwirth, KH Tragl, S Zehetmayer. Vogel Study: This work was financed by a research grant of the ‘‘Vogelstiftung Dr. Eckernkamp’’. HELIAD study: This study was supported by the grants: IIRG-09-133014 from the Alzheimer’s Association, 189 10276/8/9/2011 from the ESPA-EU program Excellence Grant (ARISTEIA) and the ΔΥ2β/οικ.51657/14.4.2009 of the Ministry for Health and Social Solidarity (Greece). Biobank Department of Psychiatry, UMG: Prof. Jens Wiltfang is supported by an Ilídio Pinho professorship and iBiMED (UID/BIM/04501/2013), and FCT project PTDC/DTP_PIC/5587/2014 at the University of Aveiro, Portugal. Lausanne study: This work was supported by grants from the Swiss National Research Foundation (SNF 320030_141179). PAGES study: Harald Hampel is an employee of Eisai Inc. During part of this work he was supported by the AXA Research Fund, the “Fondation partenariale Sorbonne Université” and the “Fondation pour la Recherche sur Alzheimer”, Paris, France. Mannheim, Germany Biobank: Department of geriatric Psychiatry, Central Institute for Mental Health, Mannheim, University of Heidelberg, Germany. Genotyping for the Swedish Twin Studies of Aging was supported by NIH/NIA grant R01 AG037985. Genotyping in TwinGene was supported by NIH/NIDDK U01 DK066134. WvdF is recipient of Joint Programming for Neurodegenerative Diseases (JPND) grants PERADES (ANR-13-JPRF-0001) and EADB (733051061). Gothenburg Birth Cohort (GBC) Studies: We would like to thank UCL Genomics for performing the genotyping analyses. The studies were supported by The Stena Foundation, The Swedish Research Council (2015-02830, 2013-8717), The Swedish Research Council for Health, Working Life and Wellfare (2013-1202, 2005-0762, 2008-1210, 2013-2300, 2013- 2496, 2013-0475), The Brain Foundation, Sahlgrenska University Hospital (ALF), The Alzheimer’s Association (IIRG-03-6168), The Alzheimer’s Association Zenith Award (ZEN-01-3151), Eivind och Elsa K:son Sylvans Stiftelse, The Swedish Alzheimer Foundation. Clinical AD, Sweden: We would like to thank UCL Genomics for performing the genotyping analyses. Barcelona Brain Biobank: Brain Donors of the Neurological Tissue Bank of the Biobanc-Hospital Clinic-IDIBAPS and their families for their generosity. Hospital Clínic de Barcelona Spanish Ministry of Economy and Competitiveness-Instituto de Salud Carlos III and Fondo Europeo de Desarrollo Regional (FEDER), Unión Europea, “Una manera de hacer Europa” grants (PI16/0235 to Dr. R. Sánchez-Valle and PI17/00670 to Dr. A.Antonelli). AA is funded by Departament de Salut de la Generalitat de Catalunya, PERIS 2016-2020 (SLT002/16/00329). Work at JP-T laboratory was possible thanks to funding from Ciberned and generous gifts from Consuelo Cervera Yuste and Juan Manuel Moreno Cervera. Sydney Memory and Ageing Study (Sydney MAS): We gratefully acknowledge and thank the following for their contributions to Sydney MAS: participants, their supporters and the Sydney MAS Research Team (current and former staff and students). Funding was awarded from the Australian National Health and Medical Research Council (NHMRC) Program Grants (350833, 568969, 109308). This work was supported by InnoMed (Innovative Medicines in Europe), an integrated project funded by the European Union of the Sixth Framework program priority (FP6-2004- LIFESCIHEALTH-5). Oviedo: This work was partly supported by Grant from Fondo de Investigaciones Sanitarias-Fondos FEDER EuropeanUnion to V.A. PI15/00878. Project MinE: The ProjectMinE study was supported by the ALS Foundation Netherlands and the MND association (UK) (Project MinE, www.projectmine.com). The SPIN cohort: We are indebted to patients and their families for their participation in the “Sant Pau Initiative on Neurodegeneration cohort”, at the Sant Pau Hospital (Barcelona). This is a multimodal research cohort for biomarker discovery and validation that is partially funded by Generalitat de Catalunya (2017 SGR 547 to JC), as well as from the Institute of Health Carlos III-Subdirección General de Evaluación and the Fondo Europeo de Desarrollo Regional (FEDER- “Una manera de Hacer Europa”) (grants PI11/02526, PI14/01126, and PI17/01019 to JF; PI17/01895 to AL), and the Centro de Investigación Biomédica en Red Enfermedades Neurodegenerativas programme (Program 1, Alzheimer Disease to AL). We would also like to thank the Fundació Bancària Obra Social La Caixa (DABNI project) to JF and AL; and Fundación BBVA (to AL), for their support in funding this follow-up study. Adolfo López de Munain is supported by Fundación Salud 2000 (PI2013156), CIBERNED and Diputación Foral de Gipuzkoa (Exp.114/17). P.S.J. is supported by CIBERNED and Carlos III Institute of Health, Spain (PI08/0139, PI12/02288, and PI16/01652, PI20/01011), jointly funded by Fondo Europeo de Desarrollo Regional (FEDER), Unión Europea, “Una manera de hacer Europa”. We thank Biobanco Valdecilla for their support. Amsterdam dementia Cohort (ADC): Research of the Alzheimer center Amsterdam is part of the neurodegeneration research program of Amsterdam Neuroscience. The AlzheimerCenter Amsterdam is supported by Stichting Alzheimer Nederland and Stichting VUmc fonds. The clinical database structure was developed with funding from Stichting Dioraphte. Genotyping of the Dutch case-control samples was performed in the context of EADB (European Alzheimer&Dementia biobank) funded by the JPco-fuND FP-829-029 (ZonMW project number #733051061). This research is performed by using data from the Parelsnoer Institute an initiative of the Dutch Federation of University Medical Centres (www.parelsnoer.org). 100-Plus study: We are grateful for the collaborative efforts of all participating centenarians and their family members and/or relations. We thank the Netherlands Brain Bank for supplying DNA for genotyping. This work was supported by Stichting AlzheimerNederland (WE09.2014-03), Stichting Diorapthe, Horstingstuit foundation, Memorabel (ZonMW project number #733050814, #733050512) and Stichting VUmcFonds. Additional support for EADB cohorts: WF, SL, HH are recipients of

ABOARD, a public-private partnership receiving funding from ZonMW (#73305095007) and Health~Holland, Topsector Life Sciences & Health (PPP-allowance; #LSHM20106). The DELCODE study was funded by the German Center for Neurodegenerative Diseases (Deutsches Zentrum für Neurodegenerative Erkrankungen (DZNE)), reference number BN012.

**Gra@ce.** The Genome Research @ Fundació ACE project (GR@ACE) is supported by Grifols SA, Fundación bancaria 'La Caixa', Fundació ACE, and CIBERNED (Centro de Investigación Biomédica en Red Enfermedades Neurodegenerativas (Program 1, Alzheimer Disease to MB and AR)). A.R. and M.B. receive support from the European Union/EFPIA Innovative Medicines Initiative Joint undertaking ADAPTED and MOPEAD projects (grant numbers 115975 and 115985, respectively). M.B. and A.R. are also supported by national grants PI13/02434, PI16/01861, PI17/01474 and PI19/01240. Acción Estratégica en Salud is integrated into the Spanish National R + D + I Plan and funded by ISCIII (Instituto de Salud Carlos III)–Subdirección General de Evaluación and the Fondo Europeo de Desarrollo Regional (FEDER–'Una manera de hacer Europa'). Some control samples and data from patients included in this study were provided in part by the National DNA Bank Carlos III (www.bancoadn.org, University of Salamanca, Spain) and Hospital Universitario Virgen de Valme (Sevilla, Spain); they were processed following standard operating procedures with the appropriate approval of the Ethical and Scientific Committee. The present work has been performed as part of the doctoral program of I. de Rojas at the Universitat de Barcelona (Barcelona, Spain).

**EADI**. This work has been developed and supported by the LABEX (laboratory of excellence program investment for the future) DISTALZ grant (Development of Innovative Strategies for a Transdisciplinary approach to ALZheimer's disease) including funding from MEL (Metropole européenne de Lille), ERDF (European Regional Development Fund) and Conseil Régional Nord Pas de Calais. This work was supported by Inserm, the National Foundation for Alzheimer's disease and related disorders, the Institut Pasteur de Lille and the Centre National de Recherche en Génomique Humaine, CEA, the JPND PERADES, the Laboratory of Excellence GENMED (Medical Genomics) grant no. ANR-10-LABX-0013 managed by the National Research Agency (ANR) part of the Investment for the Future program, and the FP7 AgedBrainSysBio. The Three-City Study was performed as part of collaboration between the Institut National de la Santé et de la Recherche Médicale (Inserm), the Victor Segalen Bordeaux II University and Sanofi-Synthélabo. The Fondation pour la Recherche Médicale funded the preparation and initiation of the study. The 3C Study was also funded by the Caisse Nationale Maladie des Travailleurs Salariés, Direction Générale de la Santé, MGEN, Institut de la Longévité, Agence Française de Sécurité Sanitaire des Produits de Santé, the Aquitaine and Bourgogne Regional Councils, Agence Nationale de la Recherche, ANR supported the COGINUT and COVADIS projects. Fondation de France and the joint French Ministry of Research/Inserm "Cohortes et collections de données biologiques" programme.

Lille Génopôle received an unconditional grant from Eisai. The Three-city biological bank was developed and maintained by the laboratory for genomic analysis LAG-BRC - Institut Pasteur de Lille.

**GERAD/PERADES**. We thank all individuals who participated in this study. Cardiff University was supported by the Wellcome Trust, Alzheimer's Society (AS; grant RF014/164), the Medical Research Council (MRC; grants G0801418/1, MR/K013041/1, MR/L023784/1), the European Joint Programme for Neurodegenerative Disease (JPND, grant MR/L501517/1), Alzheimer's Research UK (ARUK, grant ARUK-PG2014-1), Welsh Assembly Government (grant SGR544:CADR), a donation from the Moondance Charitable Foundation, UK Dementia's Platform (DPUK, reference MR/L023784/1), and the UK Dementia Research Institute at Cardiff. Cambridge University acknowledges support from the MRC. ARUK supported sample collections at the Kings College London, the South West Dementia Bank, Universities of Cambridge, Nottingham, Manchester and Belfast. King's College London was supported by the NIHR Biomedical Research Centre for Mental Health and Biomedical Research Unit for Dementia at the South London and Maudsley NHS Foundation Trust and Kings College London and the MRC. Alzheimer's Research UK (ARUK) and the Big Lottery Fund provided support to Nottingham University. Ulster Garden Villages, AS, ARUK, American Federation for Aging Research, NI R&D Office and the Royal College of Physicians/Dunhill Medical Trust provided support for Queen's University, Belfast. The University of Southampton acknowledges support from the AS. The MRC and Mercer's Institute for Research on Ageing supported the Trinity College group. DCR is a Wellcome Trust Principal Research fellow. The South West Dementia Brain Bank acknowledges support from Bristol Research into Alzheimer's and Care of the Elderly. The Charles Wolfson Charitable Trust supported the OPTIMA group. Washington University was funded by NIH grants, Barnes Jewish Foundation and the Charles and Joanne Knight Alzheimer's Research Initiative. Patient recruitment for the MRC Prion Unit/UCL Department of Neurodegenerative Disease collection was supported by the UCLH/UCL Biomedical Research Centre and their work was supported by the NIHR Queen Square Dementia BRU, the Alzheimer's Research UK and the Alzheimer's Society. LASER-AD was funded by Lundbeck SA. The AgeCoDe study group was supported by the German Federal Ministry for Education and Research grants 01 GI 0710, 01 GI 0712, 01 GI 0713, 01 GI 0714, 01 GI 0715, 01 GI 0716, 01 GI 0717. Genotyping of the Bonn case-control sample was funded by the German centre for Neurodegenerative Diseases (DZNE), Germany. The GERAD Consortium also used samples ascertained by the NIMH AD Genetics Initiative. HH was supported by a grant of the Katharina-Hardt-Foundation, Bad Homburg vor der Höhe, Germany. The KORA F4 studies were financed by Helmholtz Zentrum München; German Research Center for Environmental Health; BMBF; German National Genome Research Network and the Munich Center of Health Sciences. The Heinz Nixdorf Recall cohort was funded by the Heinz Nixdorf Foundation and BMBF. We acknowledge use of genotype data from the 1958 Birth Cohort collection and National Blood Service, funded by the MRC and the Wellcome Trust which was genotyped by the Wellcome Trust Case Control Consortium and the Type-1 Diabetes Genetics Consortium, sponsored by the National Institute of Diabetes and Digestive and Kidney Diseases, National Institute of Allergy and Infectious Diseases, National Human Genome Research Institute, National Institute of Child Health and Human Development and Juvenile Diabetes Research Foundation International. The project is also supported through the following funding organisations under the aegis of JPND - www.jpnd.eu (United Kingdom, Medical Research Council (MR/L501529/1; MR/R024804/1) and Economic and Social Research Council (ES/L008238/1)) and through the Motor Neurone Disease Association. This study represents independent research part funded by the National Institute for Health Research (NIHR) Biomedical Research Centre at South London and Maudsley NHS Foundation Trust and King's College London. Prof Jens Wiltfang is supported by an Ilídio Pinho professorship and iBiMED (UID/BIM/04501/2013), at the University of Aveiro, Portugal.

**Rotterdam study. Rotterdam (RS)**. This study was funded by the Netherlands Organisation for Health Research and Development (ZonMW) as part of the Joint Programming for Neurological Disease (JPND) as part of the PERADES Program (Defining Genetic Polygenic, and Environmental Risk for Alzheimer's disease using multiple powerful cohorts, focused Epigenetics and Stem cell metabolomics), Project number 733051021. This work was funded also by the European Union Innovative Medicine Initiative (IMI) programme under grant agreement No. 115975 as part of the Alzheimer's Disease Apolipoprotein Pathology for Treatment Elucidation and Development (ADAPTED, https://www.imi-adapted.eu) and the European Union's Horizon 2020 research and innovation programme as part of the Common mechanisms and pathways in Stroke and Alzheimer's disease CoSTREAM project (www.costream.eu, grant agreement No. 667375). The current study is supported by the Deltaplan Dementie and Memorabel supported by ZonMW (Project number 733050814) and Alzheimer Nederland. The Rotterdam Study is funded by Erasmus Medical Center and Erasmus University, Rotterdam, Netherlands Organization for the Health Research and Development (ZonMw), the Research Institute for Diseases in the Elderly (RIDE), the Ministry of Education, Culture and Science, the Ministry for Health, Welfare and Sports, the European Commission (DG XII), and the Municipality of Rotterdam. The authors are grateful to the study participants, the staff from the Rotterdam Study and the participating general practitioners and pharmacists. The generation and management of GWAS genotype data for the Rotterdam Study (RS-I, RS-II, RS-III) was executed by the Human Genotyping Facility of the Genetic Laboratory of the Department of Internal Medicine, Erasmus MC, Rotterdam, The Netherlands. The GWAS datasets are supported by the Netherlands Organization of Scientific Research NWO Investments (Project number 175.010.2005.011, 911-03-012), the Genetic Laboratory of the Department of Internal Medicine, Erasmus MC, the Research Institute for Diseases in the Elderly (014-93-015; RIDE2), the Netherlands Genomics Initiative (NGI)/Netherlands Organization for Scientific Research (NWO) Netherlands Consortium for Healthy Aging (NCHA), project number 050-060-810. We thank Pascal Arp, Mila Jhamai,

Marijn Verkerk, Lizbeth Herrera and Marjolein Peters, MSc, and Carolina Medina-Gomez, MSc, for their help in creating the GWAS database, and Karol Estrada, PhD, Yurii Aulchenko, PhD, and Carolina Medina- Gomez, MSc, for the creation and analysis of imputed data.

**DemGene.** The project has received funding from The Research Council of Norway (RCN) Grant Nos. 213837, 223273, 225989, 248778, and 251134 and EU JPND Program RCN Grant Nos. 237250, 311993, the South-East Norway Health Authority Grant No. 2013-123, the Norwegian Health Association, and KG Jebsen Foundation. The RCN FRIPRO Mobility grant scheme (FRICON) is co-funded by the European Union's Seventh Framework Programme for research, technological development and demonstration under Marie Curie grant agreement No 608695. European Community's grant PIAPP-GA-2011-286213 PsychDPC.

**Bonn study.** This group would like to thank Dr. Heike Koelsch for her scientific support. The Bonn group was funded by the German Federal Ministry of Education and Research (BMBF): Competence Network Dementia (CND) grant number 01GI0102, 01GI0711, 01GI042

**ADGC.** The National Institutes of Health, National Institute on Aging (NIH-NIA) supported this work through the following grants: ADGC, U01 AG032984, RC2 AG036528; Samples from the National Cell Repository for Alzheimer's Disease (NCRAD), which receives government support under a cooperative agreement grant (U24 AG21886) awarded by the National Institute on Aging (NIA), were used in this study. We thank contributors who collected samples used in this study, as well as patients and their families, whose help and participation made this work possible; Data for this study were prepared, archived, and distributed by the National Institute on Aging Alzheimer's Disease Data Storage Site (NIAGADS) at the University of Pennsylvania (U24-AG041689-01); NACC, U01 AG016976; NIA LOAD (Columbia University), U24 AG026395, U24 AG026390, R01AG041797; Banner Sun Health Research Institute P30 AG019610; Boston University, P30 AG013846, U01 AG10483, R01 CA129769, R01 MH080295, R01 AG017173, R01 AG025259, R01 AG048927, R01AG33193, R01 AG009029; Columbia University, P50 AG008702, R37 AG015473, R01 AG037212, R01 AG028786; Duke University, P30 AG028377, AG05128; Einstein Aging Study NIA grant at Albert Einstein College of Medicine, P01 AG03949. Emory University, AG025688; Group Health Research Institute, UO1 AG006781, UO1 HG004610, UO1 HG006375, U01 HG008657; Indiana University, P30 AG10133, R01 AG009956, RC2 AG036650; Johns Hopkins University, P50 AG005146, R01 AG020688; Massachusetts General Hospital, P50 AG005134; Mayo Clinic, P50 AG016574, R01 AG032990, KL2 RR024151; Mount Sinai School of Medicine, P50 AG005138, P01 AG002219; New York University, P30 AG08051, UL1 RR029893, 5R01AG012101, 5R01AG022374, 5R01AG013616, 1RC2AG036502, 1R01AG035137; North Carolina A&T University, P20 MD000546, R01 AG28786-01A1; Northwestern University, P30 AG013854; Oregon Health & Science University, P30 AG008017, R01 AG026916; Rush University, P30 AG010161, R01 AG019085, R01 AG15819, R01 AG17917, R01 AG030146, R01 AG01101, RC2 AG036650, R01 AG22018;

TGen, R01 NS059873; University of Alabama at Birmingham, P50 AG016582; University of Arizona, R01 AG031581; University of California, Davis, P30 AG010129; University of California, Irvine, P50 AG016573; University of California, Los Angeles, P50 AG016570; University of California, San Diego, P50 AG005131; University of California, San Francisco, P50 AG023501, P01 AG019724; University of Kentucky, P30 AG028383, AG05144; University of Michigan, P30 AG053760 and AG063760; University of Pennsylvania, P30 AG010124; University of Pittsburgh, P50 AG005133, AG030653, AG041718, AG07562, AG02365; University of Southern California, P50 AG005142; University of Texas Southwestern, P30 AG012300; University of Miami, R01 AG027944, AG010491, AG027944, AG021547, AG019757; University of Washington, P50 AG005136, R01 AG042437; University of Wisconsin, P50 AG033514; Vanderbilt University, R01 AG019085; and Washington University, P50 AG005681, P01 AG03991, P01 AG026276. HP was supported by AG025711. ER was supported by CCNA. The Kathleen Price Bryan Brain Bank at Duke University Medical Center is funded by NINDS grant # NS39764, NIMH MH60451 and by Glaxo Smith Kline. Support was also from the Alzheimer's Association (LAF, IIRG-08-89720; MP-V, IIRG-05-14147), the US Department of Veterans Affairs Administration, Office of Research and Development, Biomedical Laboratory Research Program, and BrightFocus Foundation (MP-V, A2111048). P.S.G.-H. is supported by Wellcome Trust, Howard Hughes Medical Institute, and the Canadian Institute of Health Research. Genotyping of the TGEN2 cohort was supported by Kronos Science. The TGen series was also funded by NIA grant AG041232 to AJM and MJH, The Banner Alzheimer's Foundation, The Johnnie B. Byrd Sr. Alzheimer's Institute, the Medical Research Council, and the state of Arizona and also includes samples from the following sites: Newcastle Brain Tissue Resource (funding via the Medical Research Council, local NHS trusts and Newcastle University), MRC London Brain Bank for Neurodegenerative Diseases (funding via the Medical Research Council), South West Dementia Brain Bank (funding via numerous sources including the Higher Education Funding Council for England (HEFCE), Alzheimer's Research Trust (ART), BRACE as well as North Bristol NHS Trust Research and Innovation department and DeNDRoN), The Netherlands Brain Bank (funding via numerous sources including Stichting MS Research, Brain Net Europe, Hersenstichting Nederland Breinbrekend Werk, International Parkinson Fonds, Internationale Stiching Alzheimer Onderzoek), Institut de Neuropatologia, Servei Anatomia Patologica, Universitat de Barcelona. ADNI data collection and sharing was funded by the National Institutes of Health Grant U01 AG024904 and Department of Defense award number W81XWH-12-2-0012. ADNI is funded by the National Institute on Aging, the National Institute of Biomedical Imaging and Bioengineering, and through generous contributions from the following: AbbVie, Alzheimer's Association; Alzheimer's Drug Discovery Foundation; Araclon Biotech; BioClinica, Inc.; Biogen; Bristol-Myers Squibb Company; CereSpir, Inc.; Eisai Inc.; Elan Pharmaceuticals, Inc.; Eli Lilly and Company; EuroImmun; F. Hoffmann-La Roche Ltd and its affiliated company Genentech, Inc.; Fujirebio; GE Healthcare; IXICO Ltd.; Janssen Alzheimer Immunotherapy Research & Development, LLC.; Johnson & Johnson Pharmaceutical Research & Development LLC.; Lumosity;

Lundbeck; Merck & Co., Inc.; Meso Scale Diagnostics, LLC.; NeuroRx Research; Neurotrack Technologies; Novartis Pharmaceuticals Corporation; Pfizer Inc.; Piramal Imaging; Servier; Takeda Pharmaceutical Company; and Transition Therapeutics. The Canadian Institutes of Health Research is providing funds to support ADNI clinical sites in Canada. Private sector contributions are facilitated by the Foundation for the National Institutes of Health (www.fnih.org). The grantee organization is the Northern California Institute for Research and Education, and the study is coordinated by the Alzheimer's Disease Cooperative Study at the University of California, San Diego. ADNI data are disseminated by the Laboratory for Neuro Imaging at the University of Southern California. We thank Drs. D. Stephen Snyder and Marilyn Miller from NIA who are ex-officio ADGC members. FTLD-TDP GWAS: National Institute on Aging (AG101024, AG066597 and AG017586)

**CHARGE**. **Cardiovascular Health Study (CHS)**. This CHS research was supported by NHLBI contracts HHSN268201200036C, HHSN268200800007C, HHSN268201800001C, N01HC55222, N01HC85079, N01HC85080, N01HC85081, N01HC85082, N01HC85083, N01HC85086, 75N92021D00006; and NHLBI grants U01HL080295, U01HL130114, R01HL087652, R01HL105756, R01HL103612, R01HL120393 and 75N92021D00006 with additional contribution from the National Institute of Neurological Disorders and Stroke (NINDS). Additional support was provided through R01AG023629, R01AG033193, R01AG15928, R01AG20098, and U01AG049505 from the National Institute on Aging (NIA). A full list of principal CHS investigators and institutions can be found at CHS-NHLBI.org. The provision of genotyping data was supported in part by the National Center for Advancing Translational Sciences, CTSI grant UL1TR001881, and the National Institute of Diabetes and Digestive and Kidney Disease Diabetes Research Center (DRC) grant DK063491 to the Southern California Diabetes Endocrinology Research Center. **Framingham Heart Study**. This work was supported by the National Heart, Lung, and Blood Institute's Framingham Heart Study (contracts N01-HC-25195 and HHSN268201500001I). This study was also supported by grants from the National Institute on Aging: R01AG033193, U01AG049505, U01AG52409, R01AG054076, RF1AG0059421 (S. Seshadri). S. Seshadri and A.L.D. were also supported by additional grants from the National Institute on Aging (R01AG049607, R01AG033040, RF1AG0061872, U01AG058589) and the National Institute of Neurological Disorders and Stroke (R01-NS017950, NS100605). The content is solely the responsibility of the authors and does not necessarily represent the official views of the US National Institutes of Health.

**FinnGen.** We want to acknowledge the participants and investigators of FinnGen study. The FinnGen project is funded by two grants from Business Finland (HUS 4685/31/2016 and UH 4386/31/2016) and the following industry partners: AbbVie Inc., AstraZeneca UK Ltd, Biogen MA Inc., Bristol Myers Squibb (and Celgene Corporation & Celgene International II Sàrl), Genentech Inc., Merck Sharp & Dohme LCC, Pfizer Inc., GlaxoSmithKline Intellectual Property Development Ltd., Sanofi US Services Inc., Maze Therapeutics Inc., Janssen Biotech Inc, Novartis Pharma AG, and Boehringer Ingelheim International GmbH. Following biobanks are acknowledged for delivering biobank samples to FinnGen: Auria Biobank (www.auria.fi/biopankki), THL Biobank (www.thl.fi/biobank), Helsinki Biobank (www.helsinginbiopankki.fi), Biobank Borealis of Northern Finland (https://www.ppshp.fi/Tutkimus-ja-opetus/Biopankki/Pages/Biobank-Borealis-briefly-in-English.aspx), Finnish Clinical Biobank Tampere (www.tays.fi/en-US/Research_and_development/Finnish_Clinical_Biobank_Tampere), Biobank of Eastern Finland (www.ita-suomenbiopankki.fi/en), Central Finland Biobank (www.ksshp.fi/fi-FI/Potilaalle/Biopankki), Finnish Red Cross Blood Service Biobank (www.veripalvelu.fi/verenluovutus/biopankkitoiminta), Terveystalo Biobank (www.terveystalo.com/fi/Yritystietoa/Terveystalo-Biopankki/Biopankki/) and Arctic Biobank (https://www.oulu.fi/en/university/faculties-and-units/faculty-medicine/northern-finland-birth-cohorts-and-arctic-biobank). All Finnish Biobanks are members of BBMRI.fi infrastructure (www.bbmri.fi). Finnish Biobank Cooperative -FINBB (https://finbb.fi/) is the coordinator of BBMRI-ERIC operations in Finland. The Finnish biobank data can be accessed through the Fingenious® services (https://site.fingenious.fi/en/) managed by FINBB.

### I. Supplementary Figures

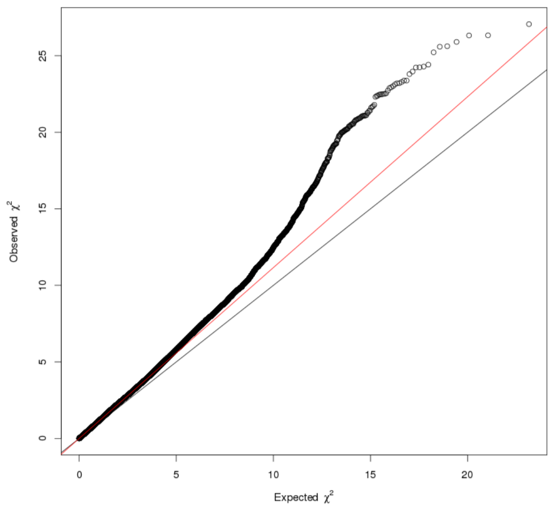


a)

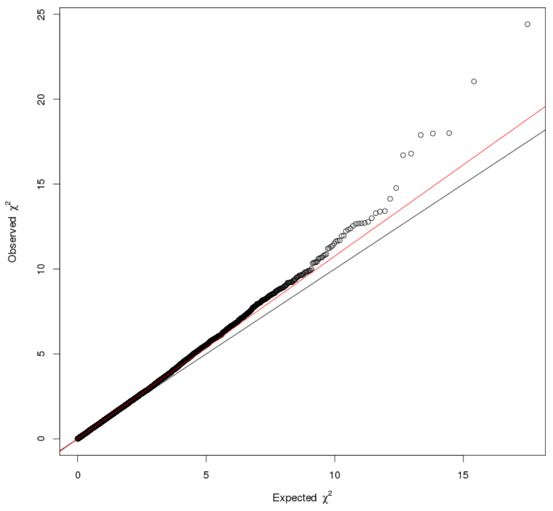


b)

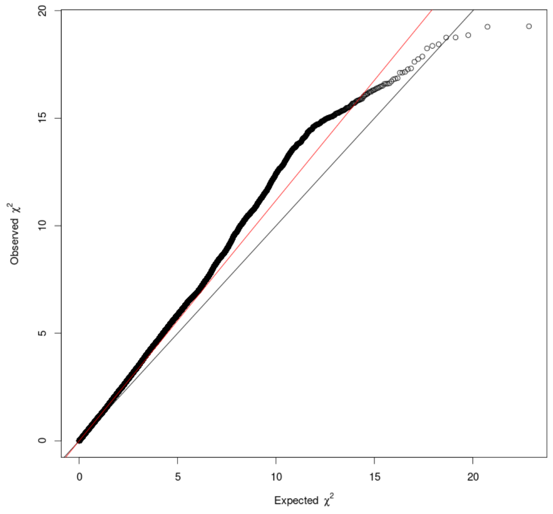


c)

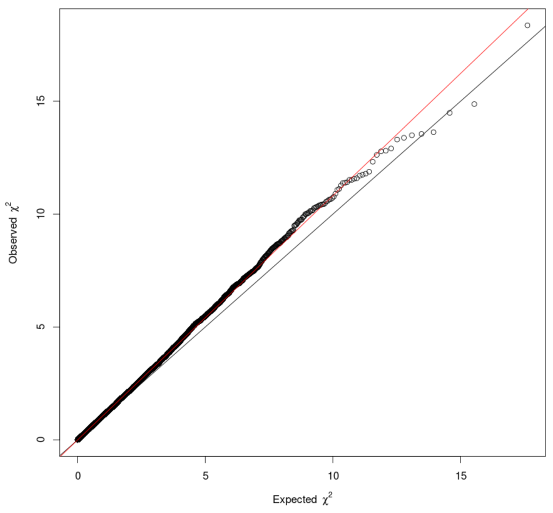


d)

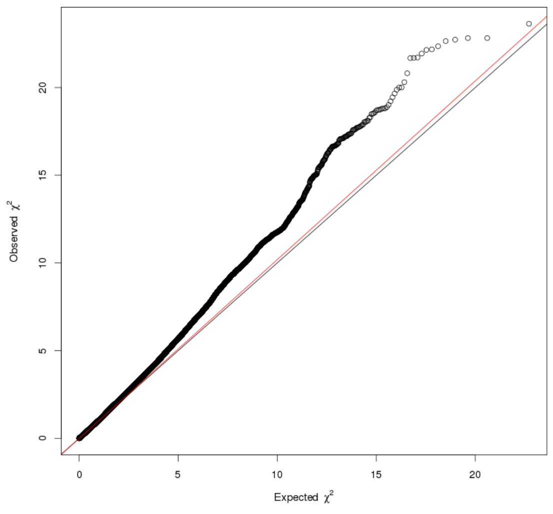


e)

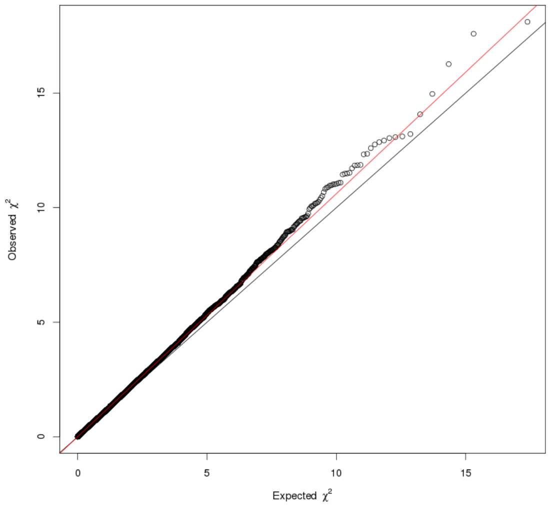


f)

Figure S1: QQ-plots of the r-XCI approach meta-analyses. The black line represents the affine function (y=x) and the red line a regression of observed values against expected values. The left column (a), c) and e)) shows QQ-plots with only common variants (MAF > 0.01), and the right column (b), d) and f)) the QQ-plots with only independent common variants (MAF > 0.01 and variants selected with the PLINK pruning procedure applied on EADB-core variants, which keeps only one variant from each pair of variants with $r^2$>0.2 and within 500 kb from each other, considering only female samples). Figures a) and b) are QQ-plots of the r-XCI meta-analysis including AD-proxy cases, with lambda = 1.116 and 1.074, respectively. Figures c) and d) are QQ-plots of the r-XCI meta-analysis including only diagnosed AD-cases, with lambda = 1.118 and 1.082, respectively. Figures e) and f) are QQ-plots of the r-XCI meta-analysis excluding biobanks, with lambda = 1.019 and 1.061, respectively.

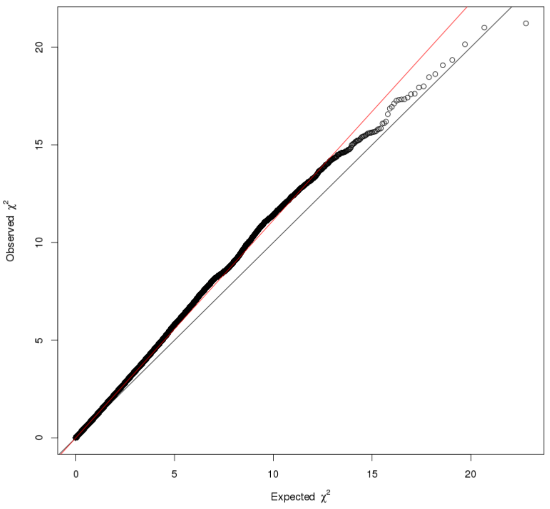


a)

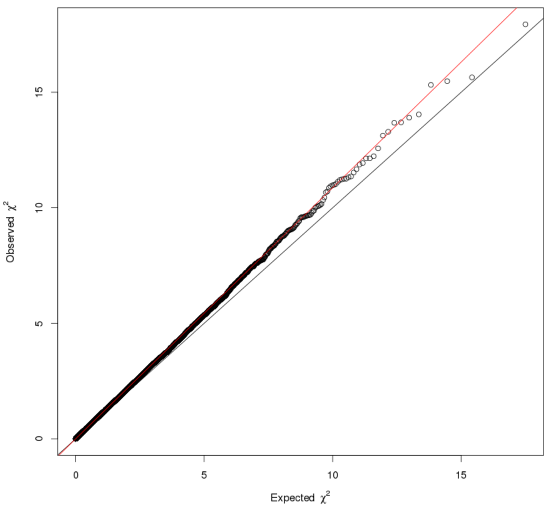


b)

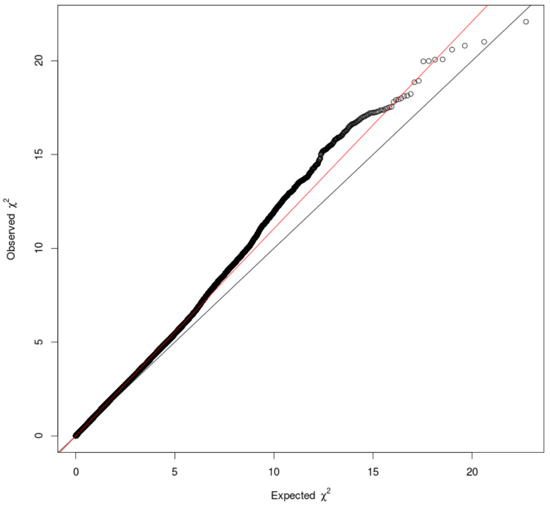


c)

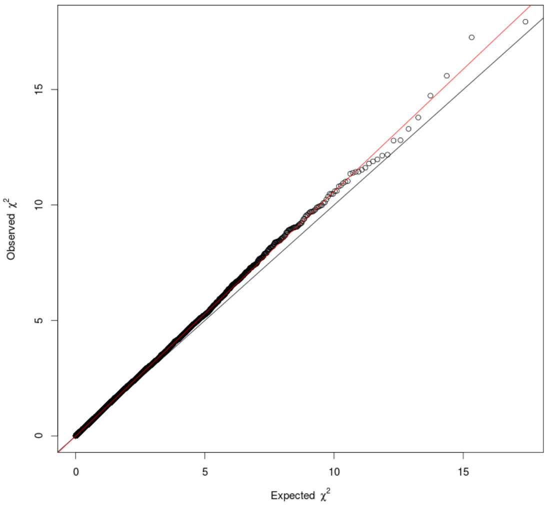


d)

Figure S2: QQ-plots of the e-XCI approach meta-analyses. The black line represents the affine function (y=x) and the red line a regression of observed values against expected values. The left column (a) and c)) shows QQ-plots with only common variants (MAF > 0.01), and the right column (b) and d)) the QQ-plots with only independent common variants (MAF > 0.01 and variants selected with the PLINK pruning procedure applied on EADB-core variants, which keeps only one variant from each pair of variants with $r^2$>0.2 and within 500 kb from each other, considering only female samples). Figures a) and b) are QQ-plots of the e-XCI meta-analysis including only diagnosed AD-cases, with lambda = 1.114 and 1.087, respectively. Figures c) and d) are QQ-plots of the e-XCI meta-analysis excluding biobanks, with lambda = 1.105 and 1.059, respectively.

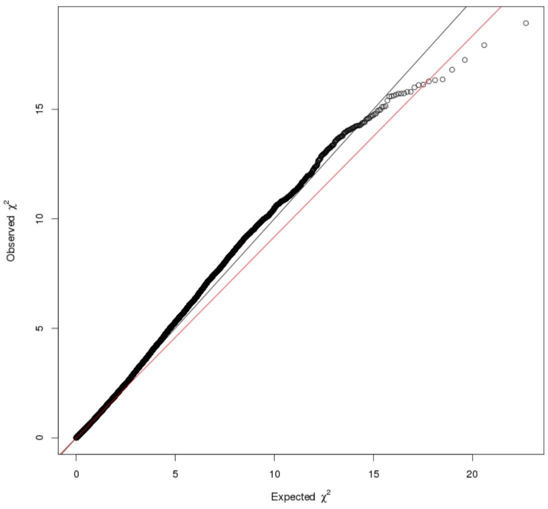


a)

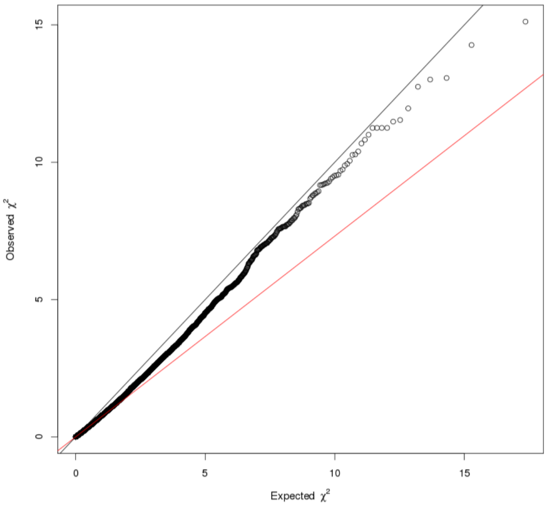


b)

Figure S3: QQ-plots of the s-XCI approach meta-analysis, including only case-control studies, with lambda = 0.914 and 0.735, respectively. The black line represents the affine function (y=x) and the red line a regression of observed values against expected values. The left column (a) shows QQ-plots with only common variants (MAF > 0.01), and the right column (b) the QQ-plots with only independent common variants (MAF > 0.01 and variants selected with the PLINK pruning procedure applied on EADB-core variants, which keeps only one variant from each pair of variants with $r^2$>0.2 and within 500 kb from each other, considering only female samples).

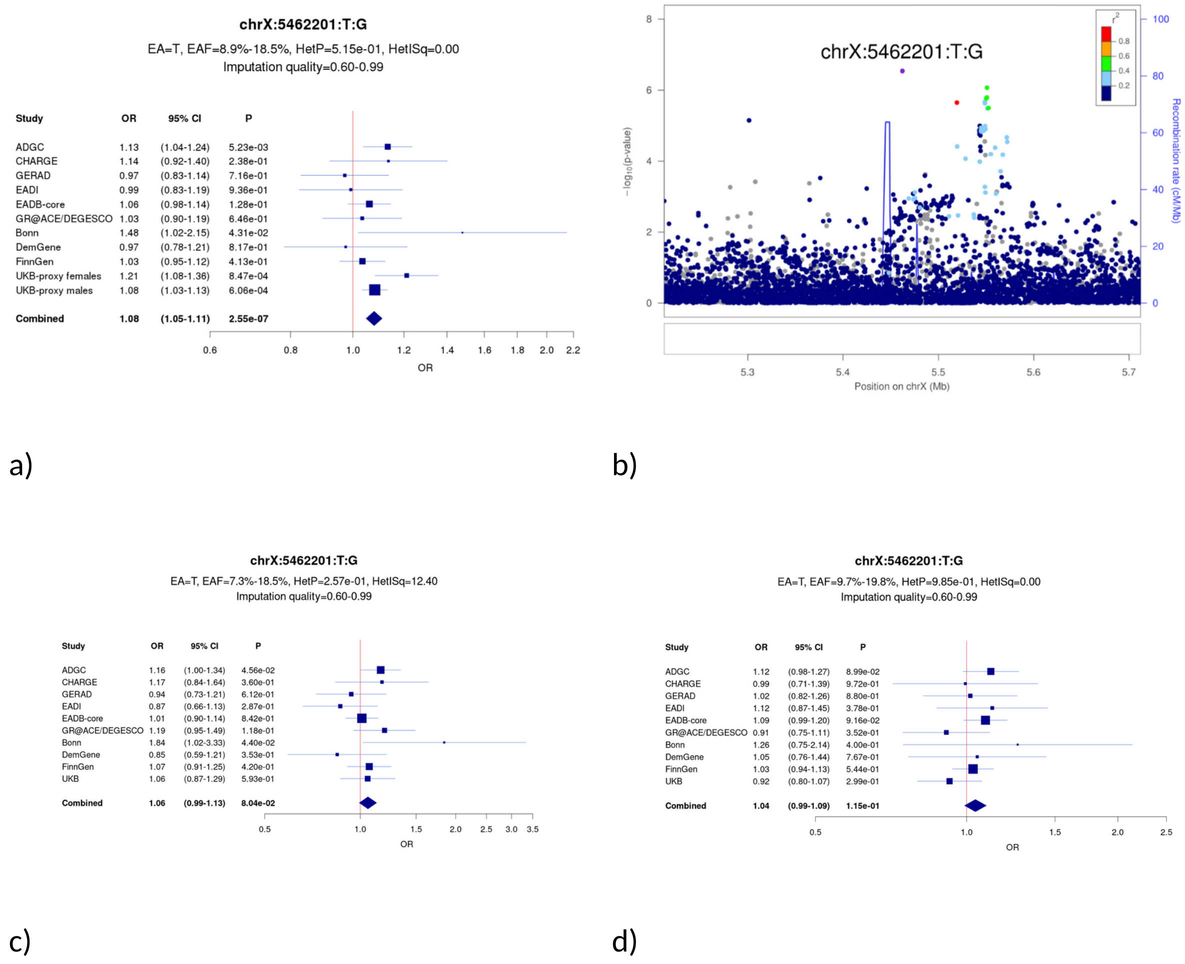


Figure S4: Forest plots and locus zoom of rs4364769 (chrX:5462201:T:G): a) forest plot and b) locus zoom in the r-XCI meta-analysis including AD-proxy, c) forest plot of female-only models excluding AD-proxy, where genotypes were coded G = {0, 0.5, 1} and d) forest plot of male-only models excluding AD-proxy, where genotypes were coded G = {0, 1}. The variant in purple is rs4364769. The positions are in GRCh38 Assembly. OR: odds ratio, CI: confidence interval, EA: effect allele, EAF: effect allele frequency range across all studies, HetP: heterogeneity P value, HetISq: heterogeneity statistic.

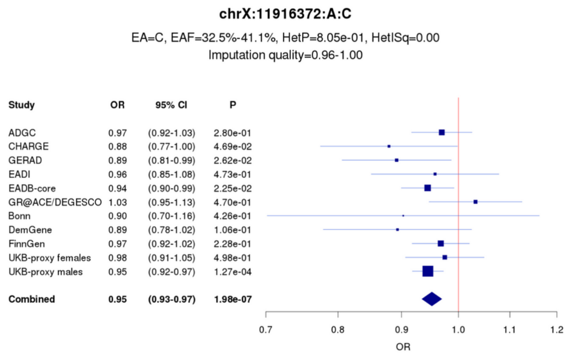


a)

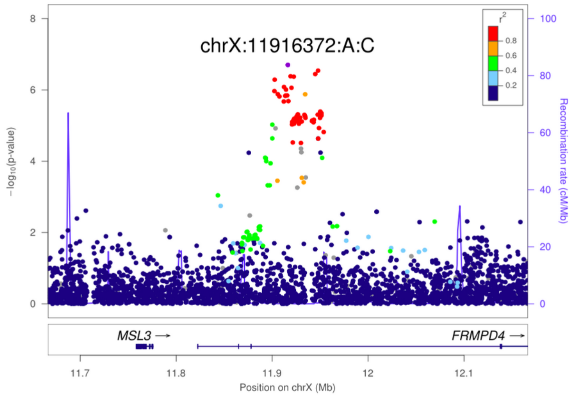


b)

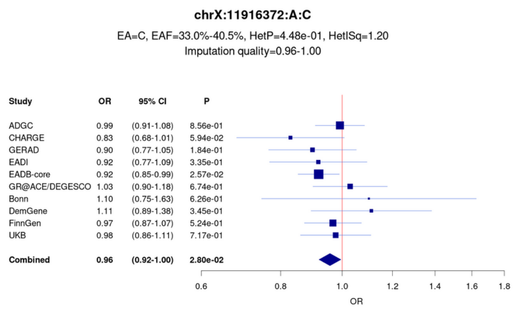


c)

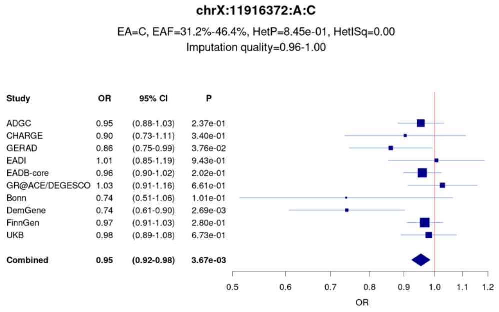


d)

Figure S5: Forest plot and locus zoom of rs5933929 (chrX:11916372:A:C): a) forest plot and b) locus zoom in the r-XCI meta-analysis including AD-proxy, c) forest plot of female-only models excluding AD-proxy, where genotypes were coded G = {0, 0.5, 1} and d) forest plot of male-only models excluding AD-proxy, where genotypes were coded G = {0, 1}. The variant in purple is rs5933929. The positions are in GRCh38 Assembly. OR: odds ratio, CI: confidence interval, EA: effect allele, EAF: effect allele frequency range across all studies, HetP: heterogeneity P value, HetISq: heterogeneity statistic.

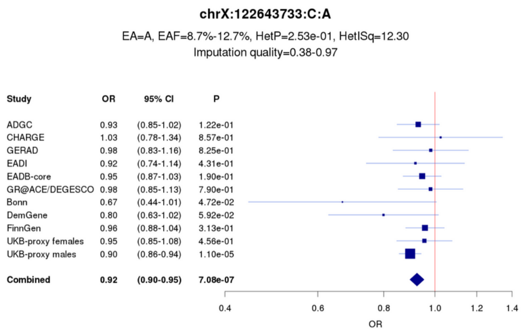


a)

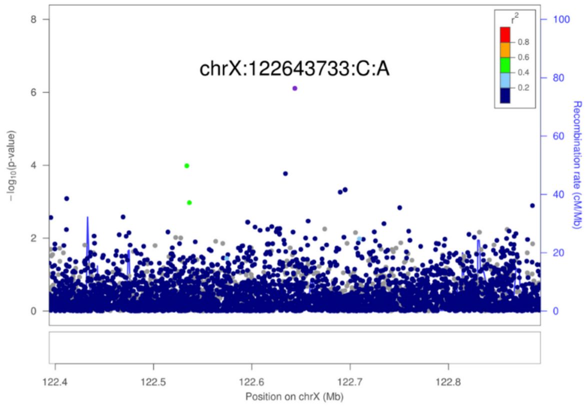


b)

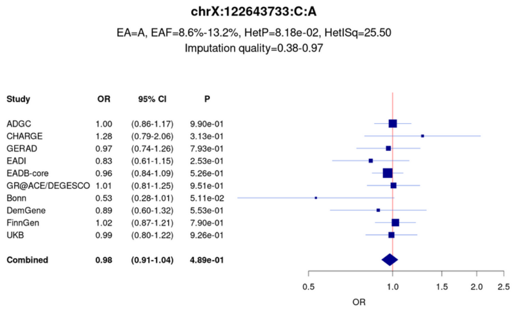


c)

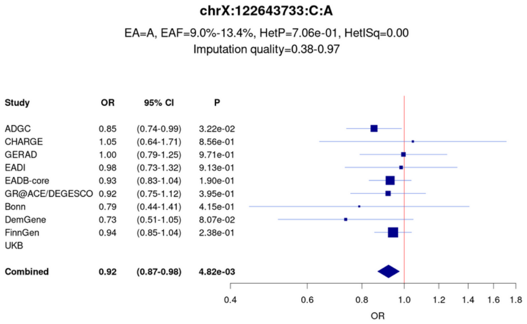


d)

Figure S6: Forest plot and locus zoom of rs191195705 (chrX:122643733:C:A): a) forest plot and b) locus zoom in the r-XCI meta-analysis including AD-proxy, c) forest plot of female-only models excluding AD-proxy, where genotypes were coded G = {0, 0.5, 1}, and d) forest plot of male-only models excluding AD-proxy, where genotypes were coded G = {0, 1}. The variant in purple is rs191195705. The positions are in GRCh38 Assembly. OR: odds ratio, CI: confidence interval, EA: effect allele, EAF: effect allele frequency range across all studies, HetP: heterogeneity P value, HetISq: heterogeneity statistic.

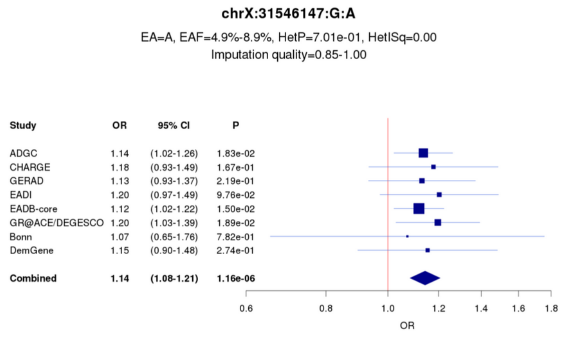


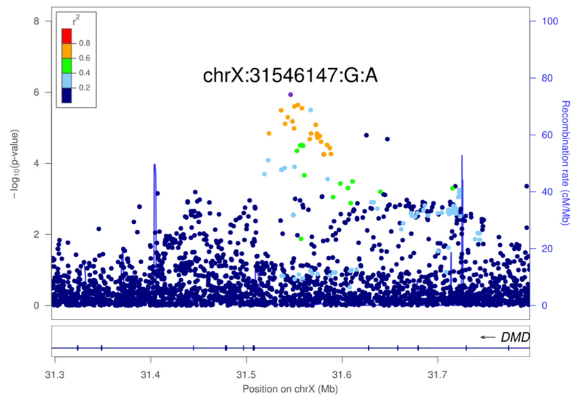


a)

b)

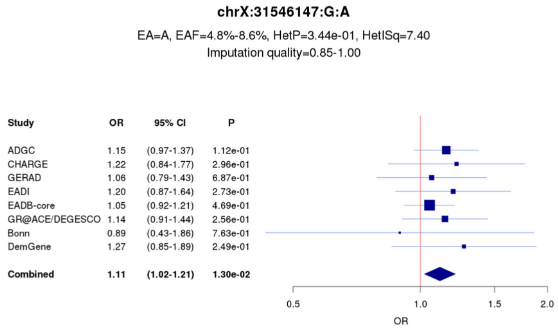


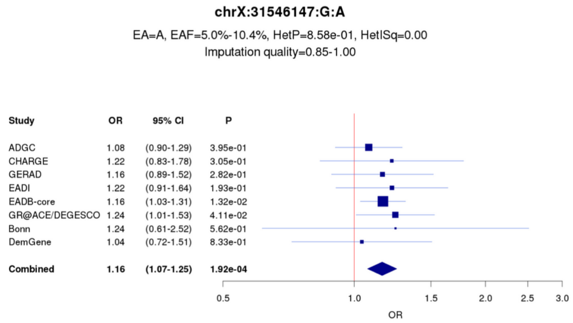


c)

d)

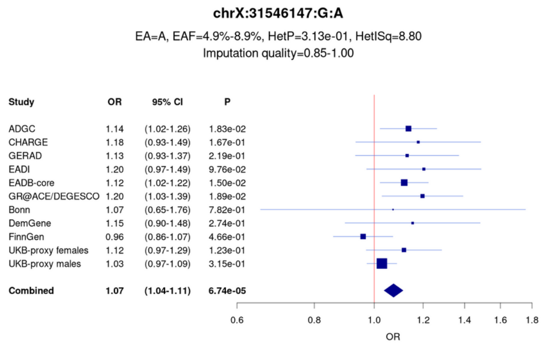


e)

Figure S7: Forest plot and locus zoom of rs5972406 (chrX:31546147:G:A): a) forest plot of the r-XCI meta-analysis excluding biobanks, b) locus zoom in the r-XCI meta-analysis excluding biobanks, c) forest plot of the female-only meta-analysis excluding biobanks and d) forest plot of the male-only meta-analysis excluding biobanks, e) forest plot of the r-XCI meta-analysis including AD-proxy cases. The variant in purple is rs5972406. The female-only and the male-only models were coded G = {0, 0.5, 1} and G = {0, 1}, respectively. The positions are in GRCh38 Assembly. OR: odds ratio, CI: confidence interval, EA: effect allele, EAF: effect allele frequency range across all studies, HetP: heterogeneity P value, HetISq:

heterogeneity statistic.

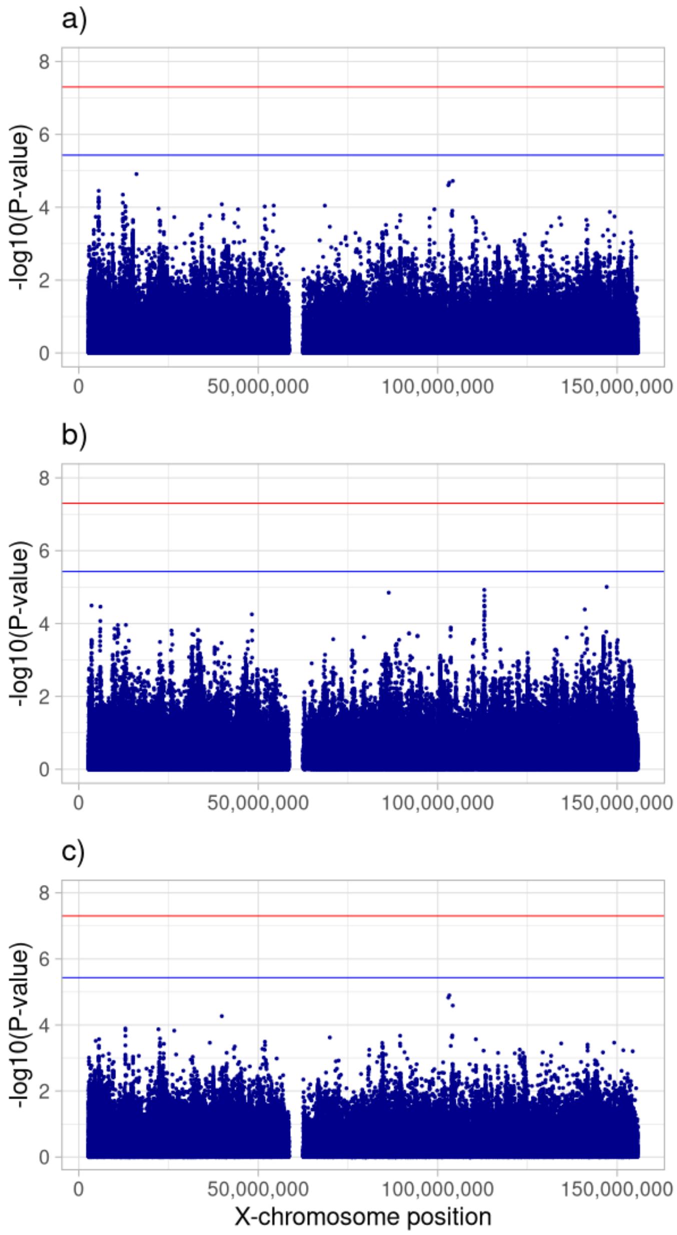


Figure S8: Manhattan plot of common variants (MAF > 0.01) for the a) female-only, b) male-only and c) interaction between genotype and sex in the meta-analysis excluding AD-proxy cases. The red and blue lines represent the genome-wide significant threshold ($5 \times 10^{-8}$) and the X-chromosome-wide significant threshold ($3.7 \times 10^{-6}$), respectively.

### J. Supplementary references


Aalten, P., Ramakers, I. H. G. B., Biessels, G. J., de Deyn, P. P., Koek, H. L., OldeRikkert, M. G. M., Oleksik, A. M., Richard, E., Smits, L. L., van Swieten, J. C., Teune, L. K., van der Lugt, A., Barkhof, F., Teunissen, C. E., Rozendaal, N., Verhey, F. R. J., & van der Flier, W. M. (2014). The Dutch Parelsnoer Institute - Neurodegenerative diseases; methods, design and baseline results. *BMC Neurology*, *14*(1), 1–8. https://doi.org/10.1186/s12883-014-0254-4

Abraham, G., Qiu, Y., & Inouye, M. (2017). FlashPCA2: principal component analysis of Biobank-scale genotype datasets. *Bioinformatics (Oxford, England)*, *33*(17), 2776–2778. https://doi.org/10.1093/bioinformatics/btx299

Beecham, G. W., Martin, E. R., Li, Y. J., Slifer, M. A., Gilbert, J. R., Haines, J. L., & Pericak-Vance, M. A. (2009). Genome-wide Association Study Implicates a Chromosome 12 Risk Locus for Late-Onset Alzheimer Disease. *American Journal of Human Genetics*, *84*(1), 35–43. https://doi.org/10.1016/j.ajhg.2008.12.008

Beekly, D. L., Ramos, E. M., Lee, W. W., Deitrich, W. D., Jacka, M. E., Wu, J., Hubbard, J. L., Koepsell, T. D., Morris, J. C., Kukull, W. A., Reiman, E. M., Kowall, N., Landreth, G., Shelanski, M., Welsh-Bohmer, K., Levey, A. I., Potter, H., Ghetti, B., Price, D., … Raskind, M. (2007). *The National Alzheimer's Coordinating Center (NACC) database: The uniform data set* (Vol. 21, Issue 3). Alzheimer Dis Assoc Disord. https://doi.org/10.1097/WAD.0b013e318142774e

Bellenguez, C., Küçükali, F., Jansen, I. E., Kleineidam, L., Moreno-Grau, S., Amin, N., Naj, A. C., Campos-Martin, R., Grenier-Boley, B., Andrade, V., Holmans, P. A., Boland, A., Damotte, V., van der Lee, S. J., Costa, M. R., Kuulasmaa, T., Yang, Q., de Rojas, I., Bis, J. C., … Lambert, J. C. (2022). New insights into the genetic etiology of Alzheimer's disease and related dementias. *Nature Genetics*, *54*(4), 412–436. https://doi.org/10.1038/s41588-022-01024-z

Braak, H., & Braak, E. (1991). *Neuropathological stageing of Alzheimer-related changes* (Vol. 82, Issue 4). Springer-Verlag. https://doi.org/10.1007/BF00308809

Bycroft, C., Freeman, C., Petkova, D., Band, G., Elliott, L. T., Sharp, K., Motyer, A., Vukcevic, D., Delaneau, O., O'Connell, J., Cortes, A., Welsh, S., Young, A., Effingham, M., McVean, G., Leslie, S., Allen, N., Donnelly, P., & Marchini, J. (2018). The UK Biobank resource with deep phenotyping and genomic data. *Nature*, *562*(7726), 203–209. https://doi.org/10.1038/s41586-018-0579-z

Caselli, R. J., Reiman, E. M., Locke, D. E. C., Hutton, M. L., Hentz, J. G., Hoffman-Snyder, C., Woodruff, B. K., Alexander, G. E., & Osborne, D. (2007). Cognitive domain decline in healthy apolipoprotein E ϵ4 homozygotes before the diagnosis of mild cognitive impairment. *Archives of Neurology*, *64*(9), 1306–1311. https://doi.org/10.1001/archneur.64.9.1306

Danecek, P., Bonfield, J. K., Liddle, J., Marshall, J., Ohan, V., Pollard, M. O., Whitwham, A., Keane, T., McCarthy, S. A., & Davies, R. M. (2021). Twelve years of SAMtools and BCFtools. *GigaScience*, *10*(2), 1–4. https://doi.org/10.1093/gigascience/giab008

De Bruijn, R. F. A. G., Akoudada, S., Lotte, L. G., Hofman, A., Niessen, W. J., Van Der Lugt, A., Koudstaal, P. J., Vernooij, M. W., & ArfanIkrama, M. M. (2014). Determinants, MRI correlates, and prognosis of mild cognitive impairment: The rotterdam study. *Journal of Alzheimer's Disease*, *42*, S239–S249. https://doi.org/10.3233/JAD-132558

De Roeck, A., Duchateau, L., Van Dongen, J., Cacace, R., Bjerke, M., Van den Bossche, T., Cras, P., Vandenberghe, R., De Deyn, P. P., Engelborghs, S., Van Broeckhoven, C., Sleegers, K., Goeman, J., Crols, R., Nuytten, D., Mercelis, R., Vandenbulcke, M., Sieben, A., De Bleecker, J. L., … Salmon, E. (2018). An intronic VNTR affects splicing of ABCA7 and increases risk of Alzheimer's disease. *Acta Neuropathologica*, *135*(6), 827–837. https://doi.org/10.1007/s00401-018-1841-z

de Rojas, I., Moreno-Grau, S., Tesi, N., Grenier-Boley, B., Andrade, V., Jansen, I. E., Pedersen, N. L., Stringa, N., Zettergren, A., Hernández, I., Montrreal, L., Antúnez, C., Antonell, A., Tankard, R. M., Bis, J. C., Sims, R., Bellenguez, C., Quintela, I., González-Perez, A., … Ruiz, A. (2021). Common variants in Alzheimer's disease and risk stratification by polygenic risk scores. *Nature Communications*, *12*(1). https://doi.org/10.1038/s41467-021-22491-8

Dubois, B., Feldman, H. H., Jacova, C., DeKosky, S. T., Barberger-Gateau, P., Cummings, J., Delacourte, A., Galasko, D., Gauthier, S., Jicha, G., Meguro, K., O'Brien, J., Pasquier, F., Robert, P., Rossor, M., Salloway, S., Stern, Y., Visser, P. J., & Scheltens, P. (2007). Research criteria for the diagnosis of Alzheimer's disease: revising the NINCDS-ADRDA criteria. *Lancet Neurology*, *6*(8), 734–746. https://doi.org/10.1016/S1474-4422(07)70178-3

Dufouil, C., Dubois, B., Vellas, B., Pasquier, F., Blanc, F., Hugon, J., Hanon, O., Dartigues, J. F., Harston, S., Gabelle, A., Ceccaldi, M., Beauchet, O., Krolak-Salmon, P., David, R., Rouaud, O., Godefroy, O., Belin, C., Rouch, I., Auguste, N., … Chêne, G. (2017). Cognitive and imaging markers in non-demented subjects attending a memory clinic: Study design and baseline findings of the MEMENTO cohort. *Alzheimer's Research and Therapy*, *9*(1), 1–13. https://doi.org/10.1186/s13195-017-0288-0

Edwards, T. L., Scott, W. K., Almonte, C., Burt, A., Powell, E. H., Beecham, G. W., Wang, L., Züchner, S., Konidari, I., Wang, G., Singer, C., Nahab, F., Scott, B., Stajich, J. M., Pericak-Vance, M., Haines, J., Vance, J. M., & Martin, E. R. (2010). Genome-Wide association study confirms SNPs in SNCA and the MAPT region as common risk factors for parkinson disease. *Annals of Human Genetics*, *74*(2), 97–109. https://doi.org/10.1111/j.1469-1809.2009.00560.x

Fried, L. P., Borhani, N. O., Enright, P., Furberg, C. D., Gardin, J. M., Kronmal, R. A., Kuller, L. H., Manolio, T. A., Mittelmark, M. B., Newman, A., O'Leary, D. H., Psaty, B., Rautaharju, P., Tracy, R. P., & Weiler, P. G. (1991). The cardiovascular health study: Design and rationale. *Annals of Epidemiology*, *1*(3), 263–276. https://doi.org/10.1016/1047-2797(91)90005-W

Green, R. C., Cupples, L. A., Go, R., Benke, K. S., Edeki, T., Griffith, P. A., Williams, M., Hipps, Y., Graff-Radford, N., Bachman, D., & Farrer, L. A. (2002). Risk of dementia among white and African American relatives of patients with Alzheimer disease. *Journal of the American Medical Association*, *287*(3), 329–336. https://doi.org/10.1001/jama.287.3.329

Hall, J. R., Wiechmann, A. R., Johnson, L. A., Edwards, M., Barber, R. C., Winter, A. S., Singh, M., & O'Bryant, S. E. (2013). Biomarkers of vascular risk, systemic inflammation, and microvascular pathology and neuropsychiatric symptoms in Alzheimer's disease. *Journal of Alzheimer's Disease*, *35*(2), 363–371. https://doi.org/10.3233/JAD-122359

Hanon, O., Vidal, J. S., Lehmann, S., Bombois, S., Allinquant, B., Tréluyer, J. M., Gelé, P., Delmaire, C., Blanc, F., Mangin, J. F., Buée, L., Touchon, J., Hugon, J., Vellas, B., Galbrun, E., Benetos, A., Berrut, G., Paillaud, E., Wallon, D., … Schraen-Maschke, S. (2018). Plasma amyloid levels within the Alzheimer's process and correlations with central biomarkers. *Alzheimer's and Dementia*, *14*(7), 858–868. https://doi.org/10.1016/j.jalz.2018.01.004

Harold, D., Abraham, R., Hollingworth, P., Sims, R., Gerrish, A., Hamshere, M. L., Pahwa, J. S., Moskvina, V., Dowzell, K., Williams, A., Jones, N., Thomas, C., Stretton, A., Morgan, A. R., Lovestone, S., Powell, J., Proitsi, P., Lupton, M. K., Brayne, C., … Williams, J. (2009). Genome-wide association study identifies variants at CLU and PICALM associated with Alzheimer's disease. *Nature Genetics*, *41*(10), 1088–1093. https://doi.org/10.1038/ng.440

Haroutunian, V., Perl, D. P., Purohit, D. P., Marin, D., Khan, K., Lantz, M., Davis, K. L., & Mohs, R. C. (1998). Regional distribution of neuritic plaques in the nondemented elderly and subjects with very mild Alzheimer Disease. *Archives of Neurology*, *55*(9), 1185–1191. https://doi.org/10.1001/archneur.55.9.1185

Holstege, H., Beker, N., Dijkstra, T., Pieterse, K., Wemmenhove, E., Schouten, K., Thiessens, L., Horsten, D., Rechtuijt, S., Sikkes, S., Van Poppel, F. W. A., Meijers-Heijboer, H., Hulsman, M., & Scheltens, P. (2018). The 100-plus study of cognitively healthy centenarians: Rationale, design and cohort description. *European Journal of Epidemiology*, *33*(12), 1229–1249. https://doi.org/10.1007/s10654-018-0451-3

Hughes, C. P., Berg, L., Danziger, W. L., Coben, L. A., & Martin, R. L. (1982). A new clinical scale for the staging of dementia. *British Journal of Psychiatry*, *140*(6), 566–572. https://doi.org/10.1192/bjp.140.6.566

Ikram, M. A., Brusselle, G. G. O., Murad, S. D., van Duijn, C. M., Franco, O. H., Goedegebure, A., Klaver, C. C. W., Nijsten, T. E. C., Peeters, R. P., Stricker, B. H., Tiemeier, H., Uitterlinden, A. G., Vernooij, M. W., & Hofman, A. (2017). The Rotterdam Study: 2018 update on objectives, design and main results. *European Journal of Epidemiology*, *32*(9), 807–850. https://doi.org/10.1007/s10654-017-0321-4

Jansen, I. E., Savage, J. E., Watanabe, K., Bryois, J., Williams, D. M., Steinberg, S., Sealock, J., Karlsson, I. K., Hägg, S., Athanasiu, L., Voyle, N., Proitsi, P., Witoelar, A., Stringer, S., Aarsland, D., Almdahl, I. S., Andersen, F., Bergh, S., Bettella, F., … Posthuma, D. (2019). Genome-wide meta-analysis identifies new loci and functional pathways influencing

Alzheimer's disease risk. *Nature Genetics*, *51*(3), 404–413. https://doi.org/10.1038/s41588-018-0311-9

Jessen, F., Wolfsgruber, S., Wiese, B., Bickel, H., Mösch, E., Kaduszkiewicz, H., Pentzek, M., Riedel-Heller, S. G., Luck, T., Fuchs, A., Weyerer, S., Werle, J., Van Den Bussche, H., Scherer, M., Maier, W., & Wagner, M. (2014). AD dementia risk in late MCI, in early MCI, and in subjective memory impairment. *Alzheimer's and Dementia*, *10*(1), 76–83. https://doi.org/10.1016/j.jalz.2012.09.017

Jun, G., Ibrahim-Verbaas, C. A., Vronskaya, M., Lambert, J. C., Chung, J., Naj, A. C., Kunkle, B. W., Wang, L. S., Bis, J. C., Bellenguez, C., Harold, D., Lunetta, K. L., Destefano, A. L., Grenier-Boley, B., Sims, R., Beecham, G. W., Smith, A. V., Chouraki, V., Hamilton-Nelson, K. L., ... Ortega-Cubero, S. (2016). A novel Alzheimer disease locus located near the gene encoding tau protein. *Molecular Psychiatry*, *21*(1), 108–117. https://doi.org/10.1038/mp.2015.23

Jun, Gyungah, Naj, A. C., Beecham, G. W., Wang, L. S., Buros, J., Gallins, P. J., Buxbaum, J. D., Ertekin-Taner, N., Fallin, M. D., Friedland, R., Inzelberg, R., Kramer, P., Rogaeva, E., St George-Hyslop, P., Cantwell, L. B., Dombroski, B. A., Saykin, A. J., Reiman, E. M., Bennett, D. A., ... Schellenberg, G. D. (2010). Meta-analysis confirms CR1, CLU, and PICALM as Alzheimer disease risk loci and reveals interactions with APOE genotypes. *Archives of Neurology*, *67*(12), 1473–1484. https://doi.org/10.1001/archneurol.2010.201

Kamboh, M. I., Minster, R. L., Demirci, F. Y., Ganguli, M., DeKosky, S. T., Lopez, O. L., & Barmada, M. M. (2012). Association of CLU and PICALM variants with Alzheimer's disease. *Neurobiology of Aging*, *33*(3), 518–521. https://doi.org/10.1016/j.neurobiolaging.2010.04.015

Kornhuber, J., Schmidtke, K., Frölich, L., Perneczky, R., Wolf, S., Hampel, H., Jessen, F., Heuser, I., Peters, O., Weih, M., Jahn, H., Luckhaus, C., Hüll, M., Gertz, H. J., Schröder, J., Pantel, J., Rienhoff, O., Seuchter, S. A., Rüther, E., ... Wiltfang, J. (2009). Early and differential diagnosis of dementia and mild cognitive impairment: *Dementia and Geriatric Cognitive Disorders*, *27*(5), 404–417. https://doi.org/10.1159/000210388

Kramer, P. L., Xu, H., Woltjer, R. L., Westaway, S. K., Clark, D., Erten-Lyons, D., Kaye, J. A., Welsh-Bohmer, K. A., Troncoso, J. C., Markesbery, W. R., Petersen, R. C., Turner, R. S., Kukull, W. A., Bennett, D. A., Galasko, D., Morris, J. C., & Ott, J. (2011). Alzheimer disease pathology in cognitively healthy elderly: A genome-wide study. *Neurobiology of Aging*, *32*(12), 2113–2122. https://doi.org/10.1016/j.neurobiolaging.2010.01.010

Kukull, W. A., Higdon, R., Bowen, J. D., McCormick, W. C., Teri, L., Schellenberg, G. D., Van Belle, G., Jolley, L., & Larson, E. B. (2002). Dementia and Alzheimer disease incidence: A prospective cohort study. *Archives of Neurology*, *59*(11), 1737–1746. https://doi.org/10.1001/archneur.59.11.1737

Kunkle, B. W., Grenier-Boley, B., Sims, R., Bis, J. C., Damotte, V., Naj, A. C., Boland, A., Vronskaya, M., van der Lee, S. J., Amlie-Wolf, A., Bellenguez, C., Frizatti, A., Chouraki, V.,

Martin, E. R., Sleegers, K., Badarinarayan, N., Jakobsdottir, J., Hamilton-Nelson, K. L., Moreno-Grau, S., … Pericak-Vance, M. A. (2019). Genetic meta-analysis of diagnosed Alzheimer's disease identifies new risk loci and implicates Aβ, tau, immunity and lipid processing. *Nature Genetics*, *51*(3), 414–430. https://doi.org/10.1038/s41588-019-0358-2

Kurki, M. I., Karjalainen, J., Palta, P., Sipilä, T. P., & Kristiansson, K. (2022). *FinnGen : Unique genetic insights from combining isolated population and national health register data*. 1–56.

Lambert, J. C., Heath, S., Even, G., Campion, D., Sleegers, K., Hiltunen, M., Combarros, O., Zelenika, D., Bullido, M. J., Tavernier, B., Letenneur, L., Bettens, K., Berr, C., Pasquier, F., Fiévet, N., Barberger-Gateau, P., Engelborghs, S., De Deyn, P., Mateo, I., … Amouyel, P. (2009). Genome-wide association study identifies variants at CLU and CR1 associated with Alzheimer's disease. *Nature Genetics*, *41*(10), 1094–1099. https://doi.org/10.1038/ng.439

Lambert, J. C., Ibrahim-Verbaas, C. A., Harold, D., Naj, A. C., Sims, R., Bellenguez, C., Jun, G., DeStefano, A. L., Bis, J. C., Beecham, G. W., Grenier-Boley, B., Russo, G., Thornton-Wells, T. A., Jones, N., Smith, A. V., Chouraki, V., Thomas, C., Ikram, M. A., Zelenika, D., … Seshadri, S. (2013). Meta-analysis of 74,046 individuals identifies 11 new susceptibility loci for Alzheimer's disease. *Nature Genetics*, *45*(12), 1452–1458. https://doi.org/10.1038/ng.2802

Larson, E. B., Wang, L., Bowen, J. D., McCormick, W. C., Teri, L., Crane, P., & Kukull, W. (2006). Exercise is associated with reduced risk for incident dementia among persons 65 years of age and older. *Annals of Internal Medicine*, *144*(2), 73–81. https://doi.org/10.7326/0003-4819-144-2-200601170-00004

Lee, J. H., Cheng, R., Graff-Radford, N., Foroud, T., & Mayeux, R. (2008). Analyses of the national institute on aging late-onset Alzheimer's disease family study: Implication of additional loci. *Archives of Neurology*, *65*(11), 1518–1526. https://doi.org/10.1001/archneur.65.11.1518

Li, H., Wetten, S., Li, L., St. Jean, P. L., Upmanyu, R., Surh, L., Hosford, D., Barnes, M. R., Briley, J. D., Borrie, M., Coletta, N., Delisle, R., Dhalla, D., Ehm, M. G., Feldman, H. H., Fornazzari, L., Gauthier, S., Goodgame, N., Guzman, D., … Roses, A. D. (2008). Candidate single-nucleotide polymorphisms from a genomewide association study of Alzheimer disease. *Archives of Neurology*, *65*(1), 45–53. https://doi.org/10.1001/archneurol.2007.3

Loh, P. R., Danecek, P., Palamara, P. F., Fuchsberger, C., Reshef, Y. A., Finucane, H. K., Schoenherr, S., Forer, L., McCarthy, S., Abecasis, G. R., Durbin, R., & Price, A. L. (2016). Reference-based phasing using the Haplotype Reference Consortium panel. *Nature Genetics*, *48*(11), 1443–1448. https://doi.org/10.1038/ng.3679

Luck, T., Riedel-Heller, S. G., Kaduszkiewicz, H., Bickel, H., Jessen, F., Pentzek, M., Wiese, B., Koelsch, H., Van Den Bussche, H., Abholz, H. H., Moesch, E., Gorfer, S., Angermeyer, M. C., Maier, W., & Weyerer, S. (2007). Mild cognitive impairment in general practice: Age-specific prevalence and correlate results from the German study on ageing, cognition and dementia in primary care patients (AgeCoDe). *Dementia and Geriatric Cognitive Disorders*, *24*(4), 307–316. https://doi.org/10.1159/000108099

Marchini, J., Howie, B., Myers, S., McVean, G., & Donnelly, P. (2007). A new multipoint method for genome-wide association studies by imputation of genotypes. *Nature Genetics*, *39*(7), 906–913. https://doi.org/10.1038/ng2088

Mbatchou, J., Barnard, L., Backman, J., Marcketta, A., Kosmicki, J. A., Ziyatdinov, A., Benner, C., O'Dushlaine, C., Barber, M., Boutkov, B., Habegger, L., Ferreira, M., Baras, A., Reid, J., Abecasis, G., Maxwell, E., & Marchini, J. (2021). Computationally efficient whole-genome regression for quantitative and binary traits. *Nature Genetics*, *53*(7), 1097–1103. https://doi.org/10.1038/s41588-021-00870-7

McKhann, G., Drachman, D., Folstein, M., Katzman, R., Price, D., & Stadlan, E. M. (1984). Clinical diagnosis of alzheimer's disease: Report of the NINCDS-ADRDA work group⋆ under the auspices of department of health and human services task force on alzheimer's disease. *Neurology*, *34*(7), 939–944. https://doi.org/10.1212/wnl.34.7.939

McKhann, G. M., Knopman, D., Chertkow, H., Hyman, B. T., Jack, C. R., Kawas, C. H., Klunk, W. E., Manly, J. J., Mayeux, R., Mohs, R. C., Morris, J. C., Rossor, M. N., Scheltens, P., Carrillo, M. C., Thies, B., Weintraub, S., & Phelps, C. H. (2011). The diagnosis of dementia due to Alzheimer's disease: Recommendations from the National Institute on Aging- Alzheimer's Association workgroups on diagnostic guidelines for Alzheimer's disease. *Alzheimers Dement*, *7*(3), 263–269. https://doi.org/10.1016/j.jalz.2011.03.005.The

Mirra, S. S., Hart, M. N., & Terry, R. D. (1993). Making the diagnosis of Alzheimer's disease: A primer for practicing pathologists. *Archives of Pathology and Laboratory Medicine*, *117*(2), 132–144. https://europepmc.org/article/med/8427562

Moreno-Grau, S., de Rojas, I., Hernández, I., Quintela, I., Montrreal, L., Alegret, M., Hernández-Olasagarre, B., Madrid, L., González-Perez, A., Maroñas, O., Rosende-Roca, M., Mauleón, A., Vargas, L., Lafuente, A., Abdelnour, C., Rodríguez-Gómez, O., Gil, S., Santos-Santos, M. Á., Espinosa, A., … Ávila, J. (2019). Genome-wide association analysis of dementia and its clinical endophenotypes reveal novel loci associated with Alzheimer's disease and three causality networks: The GR@ACE project. *Alzheimer's and Dementia*, *15*(10), 1333–1347. https://doi.org/10.1016/j.jalz.2019.06.4950

Morris, J. C., Weintraub, S., Chui, H. C., Cummings, J., DeCarli, C., Ferris, S., Foster, N. L., Galasko, D., Graff-Radford, N., Peskind, E. R., Beekly, D., Ramos, E. M., & Kukull, W. A. (2006). The Uniform Data Set (UDS): Clinical and cognitive variables and descriptive data from Alzheimer disease centers. *Alzheimer Disease and Associated Disorders*, *20*(4), 210–216. https://doi.org/10.1097/01.wad.0000213865.09806.92

Nagy, Z., Yilmazer-Hanke, D. M., Braak, H., Braak, E., Schultz, C., & Hanke, J. (1998). Assessment of the pathological stages of Alzheimer's disease in thin paraffin sections: A comparative study. *Dementia and Geriatric Cognitive Disorders*, *9*(3), 140–144. https://doi.org/10.1159/000017038

Naj, A. C., Beecham, G. W., Martin, E. R., Gallins, P. J., Powell, E. H., Konidari, I., Whitehead, P. L., Cai, G., Haroutunian, V., Scott, W. K., Vance, J. M., Slifer, M. A., Gwirtsman, H. E., Gilbert, J. R., Haines, J. L., Buxbaum, J. D., & Pericak-Vance, M. A. (2010). Dementia revealed: Novel chromosome 6 locus for Late-onset alzheimer disease provides genetic evidence for Folate-pathway abnormalities. *PLoS Genetics*, *6*(9). https://doi.org/10.1371/journal.pgen.1001130

Naj, A. C., Jun, G., Beecham, G. W., Wang, L. S., Vardarajan, B. N., Buros, J., Gallins, P. J., Buxbaum, J. D., Jarvik, G. P., Crane, P. K., Larson, E. B., Bird, T. D., Boeve, B. F., Graff-Radford, N. R., De Jager, P. L., Evans, D., Schneider, J. A., Carrasquillo, M. M., Ertekin-Taner, N., … Schellenberg, G. D. (2011). Common variants at MS4A4/MS4A6E, CD2AP, CD33 and EPHA1 are associated with late-onset Alzheimer's disease. *Nature Genetics*, *43*(5), 436–443. https://doi.org/10.1038/ng.801

Ngandu, T., Lehtisalo, J., Solomon, A., Levälahti, E., Ahtiluoto, S., Antikainen, R., Bäckman, L., Hänninen, T., Jula, A., Laatikainen, T., Lindström, J., Mangialasche, F., Paajanen, T., Pajala, S., Peltonen, M., Rauramaa, R., Stigsdotter-Neely, A., Strandberg, T., Tuomilehto, J., … Kivipelto, M. (2015). A 2 year multidomain intervention of diet, exercise, cognitive training, and vascular risk monitoring versus control to prevent cognitive decline in at-risk elderly people (FINGER): A randomised controlled trial. *The Lancet*, *385*(9984), 2255–2263. https://doi.org/10.1016/S0140-6736(15)60461-5

Nicolas, G., Wallon, D., Charbonnier, C., Quenez, O., Rousseau, S., Richard, A. C., Rovelet-Lecrux, A., Coutant, S., Le Guennec, K., Bacq, D., Garnier, J. G., Olaso, R., Boland, A., Meyer, V., Deleuze, J. F., Munter, H. M., Bourque, G., Auld, D., Montpetit, A., … Hannequin, D. (2016). Screening of dementia genes by whole-exome sequencing in early-onset Alzheimer disease: Input and lessons. *European Journal of Human Genetics*, *24*(5), 710–716. https://doi.org/10.1038/ejhg.2015.173

Niemeijer, M. N., Leening, M. J. G., Van Den Berg, M. E., Hofman, A., Franco, O. H., Deckers, J. W., Rijnbeek, P. R., Stricker, B. H., & Eijgelsheim, M. (2016). Subclinical Abnormalities in Echocardiographic Parameters and Risk of Sudden Cardiac Death in a General Population: The Rotterdam Study. *Journal of Cardiac Failure*, *22*(1), 17–23. https://doi.org/10.1016/j.cardfail.2015.06.007

Petersen, R. C., Aisen, P. S., Beckett, L. A., Donohue, M. C., Gamst, A. C., Harvey, D. J., Jack, C. R., Jagust, W. J., Shaw, L. M., Toga, A. W., Trojanowski, J. Q., & Weiner, M. W. (2010). Alzheimer's Disease Neuroimaging Initiative (ADNI): Clinical characterization. *Neurology*, *74*(3), 201–209. https://doi.org/10.1212/WNL.0b013e3181cb3e25

Petyuk, V. A., Chang, R., Ramirez-Restrepo, M., Beckmann, N. D., Henrion, M. Y. R., Piehowski, P. D., Zhu, K., Wang, S., Clarke, J., Huentelman, M. J., Xie, F., Andreev, V., Engel, A., Guettoche, T., Navarro, L., De Jager, P., Schneider, J. A., Morris, C. M., McKeith, I. G., … Myers, A. J. (2018). The human brainome: network analysis identifies HSPA2 as a novel Alzheimer's disease target. *Brain: A Journal of Neurology*, *141*(9), 2721–2739. https://doi.org/10.1093/brain/awy215

Reiman, E. M., Webster, J. A., Myers, A. J., Hardy, J., Dunckley, T., Zismann, V. L., Joshipura, K. D., Pearson, J. V, Hu-Lince, D., Huentelman, M. J. J., Craig, D. W., Coon, K. D., Liang, W. S., Herbert, R. L. H., Beach, T., Rohrer, K. C., Zhao, A. S., Leung, D., Bryden, L., … Stephan, D. A. (2007). GAB2 Alleles Modify Alzheimer's Risk in APOE ϵ4 Carriers. *Neuron*, *54*(5), 713–720. https://doi.org/10.1016/j.neuron.2007.05.022

Roccaforte, W. H., Burke, W. J., Bayer, B. L., & Wengel, S. P. (1992). Validation of a Telephone Version of the Mini-Mental State Examination. *Journal of the American Geriatrics Society*, *40*(7), 697–702. https://doi.org/10.1111/j.1532-5415.1992.tb01962.x

Ruiz, A., Dols-Icardo, O., Bullido, M. J., Pastor, P., Rodríguez-Rodríguez, E., Munain, A. L. de, de Pancorbo, M. M., Pérez-Tur, J., Álvarez, V., Antonell, A., López-Arrieta, J., Hernández, I., Tárraga, L., Boada, M., Lleó, A., Rafael Blesab, c, A., Frank, Sastre, I., Razquin, C., … Clarimón, J. (2014). Assessing the role of the TREM2 p.R47H variant as a risk factor for Alzheimer's disease and frontotemporal dementia. *Neurobiology of Aging*, *35*(2), 444.e1-444.e4. https://doi.org/https://doi.org/10.1016/j.neurobiolaging.2013.08.011

Schmermund, A., Möhlenkamp, S., Stang, A., Grönemeyer, D., Seibel, R., Hirche, H., Mann, K., Siffert, W., Lauterbach, K., Siegrist, J., Jöckel, K. H., & Erbel, R. (2002). Assessment of clinically silent atherosclerotic disease and established and novel risk factors for predicting myocardial infarction and cardiac death in healthy middle-aged subjects: Rationale and design of the Heinz Nixdorf RECALL study. *American Heart Journal*, *144*(2), 212–218. https://doi.org/10.1067/mhj.2002.123579

Scott, W. K., Nance, M. A., Watts, R. L., Hubble, J. P., Koller, W. C., Lyons, K., Pahwa, R., Stern, M. B., Colcher, A., Hiner, B. C., Jankovic, J., Ondo, W. G., Allen, F. H., Goetz, C. G., Small, G. W., Masterman, D., Mastaglia, F., Laing, N. G., Stajich, J. M., … Pericak-Vance, M. A. (2001). Complete genomic screen in parkinson disease evidence for multiple genes. *Journal of the American Medical Association*, *286*(18), 2239–2244. https://doi.org/10.1001/jama.286.18.2239

Sheardova, K., Vyhnalek, M., Nedelska, Z., Laczo, J., Andel, R., Marciniak, R., Cerman, J., Lerch, O., & Hort, J. (2019). Czech Brain Aging Study (CBAS): Prospective multicentre cohort study on risk and protective factors for dementia in the Czech Republic. *BMJ Open*, *9*(12), 1–8. https://doi.org/10.1136/bmjopen-2019-030379

Sims, R., Van Der Lee, S. J., Naj, A. C., Bellenguez, C., Badarinarayan, N., Jakobsdottir, J., Kunkle, B. W., Boland, A., Raybould, R., Bis, J. C., Martin, E. R., Grenier-Boley, B., Heilmann-Heimbach, S., Chouraki, V., Kuzma, A. B., Sleegers, K., Vronskaya, M., Ruiz, A., Graham, R. R., … Schellenberg, G. D. (2017). Rare coding variants in PLCG2, ABI3, and TREM2 implicate microglial-mediated innate immunity in Alzheimer's disease. *Nature Genetics*, *49*(9), 1373–1384. https://doi.org/10.1038/ng.3916

Stang, A., Moebus, S., Dragano, N., Beck, E. M., Möhlenkamp, S., Schmermund, A., Siegrist, J., Erbel, R., & Jöckel, K. H. (2005). Baseline recruitment and analyses of nonresponse of the Heinz Nixdorf Recall Study: Identifiability of phone numbers as the major determinant of response. *European Journal of Epidemiology*, *20*(6), 489–496. https://doi.org/10.1007/s10654-005-5529-z

Steinberg, S., Stefansson, H., Jonsson, T., Johannsdottir, H., Ingason, A., Helgason, H., Sulem, P., Magnusson, O. T., Gudjonsson, S. A., Unnsteinsdottir, U., Kong, A., Helisalmi, S., Soininen, H., Lah, J. J., Aarsland, D., Fladby, T., Ulstein, I. D., Djurovic, S., Sando, S. B., … Stefansson, K. (2015). Loss-of-function variants in ABCA7 confer risk of Alzheimer's disease. *Nature Genetics*, *47*(5), 445–447. https://doi.org/10.1038/ng.3246

Van Der Flier, W. M., & Scheltens, P. (2018). Amsterdam dementia cohort: Performing research to optimize care. *Journal of Alzheimer's Disease*, *62*(3), 1091–1111. https://doi.org/10.3233/JAD-170850

Webster, J. A., Gibbs, J. R., Clarke, J., Ray, M., Zhang, W., Holmans, P., Rohrer, K., Zhao, A., Marlowe, L., Kaleem, M., McCorquodale, D. S., Cuello, C., Leung, D., Bryden, L., Nath, P., Zismann, V. L., Joshipura, K., Huentelman, M. J., Hu-Lince, D., … Myers, A. J. (2009). Genetic Control of Human Brain Transcript Expression in Alzheimer Disease. *American Journal of Human Genetics*, *84*(4), 445–458. https://doi.org/10.1016/j.ajhg.2009.03.011
